# Multi-omics reveals selective metabolic remodeling with trace element supplementation in chronic kidney disease

**DOI:** 10.64898/2026.10.19.26365915

**Authors:** Jianyu Piao, Mingji Jin, Fengxian Yin, Zhenyu Quan, Zhonggao Gao, Zhenhua Cui

**Author notes:** **Corresponding authors:** Zhonggao Gao: Institute of Materia Medica, Chinese Academy of Medical Sciences and Peking Union Medical College, Beijing 100050, China., Zhenhua Cui: Department of Nephrology, Yanbian University Hospital, 1327 Juzi Street, Yanji 133000, Jilin, China.

## Abstract

**Introduction:** Chronic kidney disease (CKD) generates metabolically heterogeneous stress across circulating, microbial, and renal compartments. Trace element supplementation may engage redox and metabolic pathways and selectively modulate responsive metabolic programs.

**Objectives:** To define the metabolic branch selectivity, renal regional distribution, and tissue correlates of responses to a trace element cocktail (TEX) through an integrated clinical-to-mouse multi-omics framework, and to organize these findings into an experimentally testable model.

**Methods:** In a double-blind, placebo-controlled trial, 80 adults receiving maintenance hemodialysis received standard care plus TEX (20 mL/day; n = 40) or standard care plus matched placebo (n = 40). Longitudinal clinical outcomes were analyzed using participant random-intercept models. Endpoint plasma metabolomics and fecal metagenomics included healthy controls (n = 10) and voluntary trial subsets (placebo, n = 12; TEX, n = 20). Mice with 5/6 nephrectomy underwent targeted tryptophan metabolite profiling, renal spatial metabolomics, histopathology, and molecular analyses.

**Results:** Longitudinal monitoring showed no overt treatment-related laboratory safety signal. Plasma metabolomics identified treatment-associated differences (false discovery rate <0.05), including directional shifts in phospholipid and amino acid modules toward healthy reference profiles. Integrated microbial and targeted metabolic analyses resolved branch-selective responses, with TEX reducing 3-hydroxykynurenine, kynurenic acid, and xanthurenic acid and shifting the host kynurenine-branch score toward the sham profile in CKD mice. Spatial metabolomics mapped region-specific renal responses, and tissue analyses identified reduced collagen I accumulation together with increased AQP1, SIRT3, SOD2, and NRF2 signals.

**Conclusion:** This study establishes branch selectivity and renal regional specificity as defining features of TEX-associated metabolic remodeling in CKD. The integrated evidence defines a spatially organized, kynurenine-centered metabolic–renal response architecture comprising host kynurenine-branch remodeling, anatomically patterned renal metabolic responses, and selective structural, tubular, and mitochondrial/redox phenotypes, with kynurenine–mitochondrial/redox coupling as the central mechanistic proposition.

**Trial registration:** International Traditional Medicine Clinical Trial Registry, ITMCTR2026000263; retrospectively registered on 3 February 2026, after completion of recruitment and follow-up.

## Introduction

Chronic kidney disease (CKD) is a progressive systemic disorder and a major global health burden, with substantial morbidity and mortality despite advances in disease management and kidney replacement therapy.[1] Beyond declining renal filtration, CKD is characterized by persistent metabolic stress involving altered nutrient handling, redox imbalance, mitochondrial dysfunction,[2] inflammation, and accumulation of retained metabolites.[3,4] The kidney is particularly vulnerable to metabolic stress because tubular epithelial cells have high energetic demands and rely heavily on mitochondrial metabolism to sustain solute transport and specialized cellular functions.[2,4] Metabolomics is therefore valuable not only for biomarker discovery but also for defining systems-level pathway organization and treatment responsiveness.[5,6] This systems perspective motivates investigation of whether metabolic organization can be modified under persistent renal stress.

Trace elements provide a potential entry point to this question. Essential minerals serve as cofactors or structural components of enzymes involved in redox defense, mitochondrial function, intermediary metabolism, and cellular adaptation, while their systemic homeostasis is frequently altered in CKD and further influenced by dialysis.[7-9] Clinical studies of individual trace elements have produced heterogeneous biochemical and clinical responses.[10-13] Mechanistic studies of selenium biology further show that selenoprotein H can regulate MFN2-dependent mitochondrial quality control in experimental kidney injury.[14] A defined trace element cocktail therefore provides an opportunity to determine how combined elemental exposure engages selective metabolic programs across interconnected compartments, without assigning the response to any single element in advance.

The response to metabolic stress may extend across interacting systemic and tissue compartments. The gut– kidney axis links renal dysfunction, intestinal microbial ecology, and circulating metabolism, with bidirectional interactions between kidney disease, the microbiota, and microbe-derived metabolites shaping host homeostasis.[15-17] Tryptophan metabolism provides a useful framework for resolving these interactions because circulating tryptophan metabolites arise from both host and microbial pathways, including the host kynurenine pathway and microbial indole metabolism.[18-20] Kynurenine metabolites add further biological relevance through their participation in immune, inflammatory, and metabolic regulation.[21] At the same time, metabolic vulnerability within the kidney is inherently spatial: distinct anatomical regions operate within different oxygen, substrate, and energetic environments and maintain specialized metabolic programs.[22,23] Spatially resolved metabolomics can determine how these responses are distributed across anatomically defined compartments.[24]

Using a problem-oriented multi-omics strategy[25], we integrated longitudinal clinical assessment in maintenance hemodialysis patients with endpoint plasma metabolomics and fecal metagenomics, followed by targeted tryptophan-pathway profiling, renal spatial metabolomics, histopathology, and molecular analyses in an independent CKD mouse model. Rather than treating each platform as an independent endpoint, we used each layer to resolve a question raised by the preceding analysis: circulating metabolomics identifies responsive modules, metagenomics clarifies the microbial context, targeted profiling resolves branch selectivity, spatial metabolomics localizes renal responses, and tissue analyses define accompanying phenotypes. We hypothesized that TEX elicits selective metabolic responses that can be resolved progressively from circulating modules to tryptophan branches, renal regions, and tissue phenotypes. This design was intended to establish the direction, anatomical distribution, and tissue correlates of TEX-associated remodeling while generating mechanistically testable priorities.

## Material and methods

### Ethics statement

The clinical trial was approved by the Institutional Review Board and Ethics Committee of Yanbian Hospital, Yanbian University (Approval No. 2023266) on 16 February 2023, before enrollment. Written informed consent was obtained from all participants before enrollment, and the study followed the principles of the Declaration of Helsinki. Recruitment occurred from 1 March to 1 June 2023, and follow-up ended on 1 September 2024. The trial was registered with the International Traditional Medicine Clinical Trial Registry (ITMCTR2026000263; https://trialsearch.who.int/Trial2.aspx?TrialID=ITMCTR2026000263) on 3 February 2026, after recruitment and follow-up had been completed. Registration was therefore retrospective and did not meet the requirement for prospective registration. Ethics approval preceded enrollment but did not constitute prospective trial registration. Animal experiments were approved by the Laboratory Animal Ethics Committee of the Institute of Materia Medica, Peking Union Medical College (Approval No. IMM-N-26-0139) and were conducted in accordance with institutional animal care and use guidelines.

### Randomization and masking

In this randomized, double-blind, placebo-controlled, parallel-group trial, 80 adults (18–75 years) receiving maintenance hemodialysis were randomly assigned (1:1) to standard care plus oral trace element supplementation (experimental group [EG], n = 40) or standard care plus matched placebo (control group [CG], n = 40). An investigator who was not involved in participant recruitment, intervention assignment, or outcome assessment generated a computer-based random allocation sequence using permuted blocks of four without stratification. Allocation was concealed using sequentially numbered, opaque, sealed envelopes prepared in advance and opened only after eligibility was confirmed, written informed consent obtained, and baseline assessments completed. Enrolling clinicians had no access to the allocation sequence. After enrollment, a research coordinator opened the next envelope in numerical order and assigned the corresponding intervention; the coordinator could not access the sequence before opening the envelope. Participants and investigators were blinded to treatment allocation, and the placebo was matched to the active formulation. The trace element cocktail (TEX) was obtained from Beijing Jinshan Ecology Co., Ltd. (Beijing, China; food production license No. SC23221078200017) and administered orally at 20 mL/day for up to 12 months after a 1-month run-in period. Clinical laboratory parameters were collected during routine dialysis visits. A voluntary subset additionally provided endpoint blood and fecal specimens at Yanbian University Hospital for multi-omics analyses (healthy controls [HC], n = 10; CG, n = 12; EG, n = 20). Because multi-omics sampling was performed only at endpoint in a voluntary subset, these molecular analyses were interpreted as exploratory.

### Participants and follow-up

This single-center study was conducted at Yanbian University Hospital. Eligible participants were adults (18– 75 years) with CKD diagnosed and staged according to KDIGO criteria who were receiving regular maintenance hemodialysis for end-stage renal disease. Body weight, dialysis prescription, and major medications were required to be stable during the month before screening, and all participants provided written informed consent. Key exclusions were acute dialysis or intermittent kidney replacement therapy, planned kidney transplantation, recent severe cardiovascular events, uncontrolled hypertension, clinically important bleeding, malignancy, severe infection, HIV infection, active autoimmune disease, severe gastrointestinal disease or malnutrition, allergy to study materials, participation in another trial, and pregnancy or breastfeeding.

Follow-up visits were scheduled every 2–3 months with a ±14-day window. The protocol specified recording of adverse events and concomitant medications and assessment of adherence using returned packaging.

### Clinical outcomes, sample size, and protocol amendments

The primary clinical objective was to evaluate safety and tolerability. Longitudinal treatment-by-time slopes were analyzed for creatinine, BUN, cystatin C, eGFR, and parathyroid hormone; other clinical laboratory measurements were secondary indicators. The planned sample size was 40 participants per group, with no documented formal calculation based on an assumed effect size, significance level, or statistical power.

Endpoint omics sampling, originally planned in a random subset of approximately 20 EG and 10 CG participants, was ultimately conducted in voluntary subsets of 20 EG and 12 CG; together with absent baseline omics measurements, this warranted exploratory interpretation. The study design was finalized before recruitment began on 1 March 2023, and subsequent amendments did not alter the implemented trial design. Post-initiation changes comprised removal of platelet count, uric acid, carbon dioxide, total protein, prealbumin, and high-sensitivity C-reactive protein (hs-CRP) from the final reporting panel; addition of calcium–phosphorus product; reclassification of eGFR as a primary indicator; and specification of 12-month follow-up. The first five laboratory variables were absent from the final panel, whereas hs-CRP was measured only in a subset and had incomplete data. The registry listed creatinine, BUN, cystatin C, parathyroid hormone, and hs-CRP, but not eGFR, as primary outcomes. These amendments and discrepancies preclude claims of complete prospective alignment among the original study plan, registry record, and final manuscript; eGFR analyses were therefore treated as exploratory. Calcium–phosphorus product was derived and retained in Table S3 but was not analyzed as a treatment-effect outcome.

### Animal study

Male C57BL/6N mice (SPF, 8 weeks old) were obtained from Charles River Laboratories (Cat #213). The animals were group-housed under standard laboratory conditions, with temperature and humidity maintained within the animal facility’s routine ranges, adequate ventilation, and ad libitum access to standard chow and water. After 1 week of acclimatization, the mice were randomly allocated to Sham, CKD, and CKD-TEX groups using a random number generator. Chronic kidney disease was induced in the CKD and CKD-TEX groups by 5/6 nephrectomy, whereas mice in the Sham group underwent sham surgery. CKD-TEX mice received TEX diluted 300-fold in drinking water ad libitum for 8 weeks. Investigators performing histological scoring and image quantification were blinded to group allocation. At endpoint, blood and kidney tissues were collected for renal function assessment, histopathology, immunostaining, inflammatory profiling by RT-qPCR and ELISA, targeted plasma metabolite quantification, and spatial metabolomics.

### Elemental analysis by ICP-OES

The elemental composition of TEX was determined by inductively coupled plasma optical emission spectrometry (Agilent 5800). Three independent digestions of the same composite solution were analyzed (2 mL digested and diluted to a final volume of 10 mL); Fe, Mg, Al, Zn, K, and Li received an additional 20-fold dilution, whereas the other elements received no additional dilution. Full-element screening, operating conditions, replicate measurements, means, standard deviations, and coefficients of variation are provided in Table S1. Elements below the limit of detection in all replicates were excluded. Ag detected in only one replicate was excluded as an analytical artifact, as documented in Table S1. These measurements characterize the administered formulation and do not represent circulating or tissue elemental exposure in participants.

### Untargeted plasma metabolomics

Plasma samples were extracted with acetonitrile:methanol (1:4, v/v) containing internal standards, and pooled quality-control samples were analyzed alongside study samples. Profiling was performed on a Q Exactive mass spectrometer (Thermo Fisher Scientific) coupled to an ACQUITY Premier HSS T3 column (Waters), acquiring full-scan MS and data-dependent MS/MS over m/z 75–1,000 in both ionization modes. Data were processed in R and MetaboAnalystR using PCA, hierarchical clustering, and PLS-DA (999 label permutations); group comparisons used Kruskal–Wallis and Mann–Whitney U tests with Benjamini–Hochberg correction, metabolites were annotated against the KEGG Compound database, and effect sizes were summarized as Cohen’s d.

### Fecal metagenomic sequencing and analysis

Fecal genomic DNA was quality-controlled, fragmented, and subjected to shotgun library preparation and sequencing on an MGI DNBSEQ-T7 platform (BGI, Shenzhen, China). Reads were quality-filtered with fastp, host-depleted and mapped with Bowtie2, assembled with MEGAHIT, and gene-predicted with MetaGeneMark; a nonredundant catalog was built with CD-HIT, with taxonomic annotation by DIAMOND/MEGAN against the NCBI nonredundant database and functional annotation against KEGG. Abundance profiles were CLR-transformed, and microbial tryptophan biosynthetic capacity was summarized as a composite module score across seven KEGG orthologs (trpE–trpB).

### Targeted tryptophan metabolite analysis

Mouse plasma tryptophan and its metabolites were quantified by targeted LC–MS/MS on a QTRAP 6500+ system (SCIEX) using scheduled multiple reaction monitoring, with data acquired in Analyst 1.6.3 and quantified in MultiQuant 3.0.3; metabolite-by-metabolite MRM transitions were individually optimized.

### Spatial metabolomics

Frozen kidney sections (10 μm) were mounted on ITO-coated slides, coated with 2,5-dihydroxybenzoic acid matrix, and imaged on a timsTOF fleX mass spectrometer (Bruker Daltonics) in positive ion mode over m/z 50–1,300 at 50 μm spatial resolution, with MS/MS for structural confirmation. Data were RMS-normalized in SCiLS Lab and analyzed by PCA and OPLS-DA (200 permutations; differential features: VIP > 1 and P < 0.05), with regional comparisons across the cortex, outer stripe of the outer medulla (OSOM), inner stripe of the outer medulla (ISOM), and inner medulla (IM).

### Renal function, histology, immunostaining, ELISA, and RT-qPCR

Mouse Scr and BUN were measured with commercial assay kits (Cat #ADS-W-FM034 and Cat #ADS-W-N013-96; Jiangsu Aidisheng Biological Technology Co., Ltd.). Formalin-fixed, paraffin-embedded kidney sections (3 μm) were processed for H&E, PAS, IHC, and immunofluorescence using primary antibodies against AQP1, SIRT1, SIRT3, SOD2, CD38 (CST), IDH2 (Abcam), collagen type I, α-smooth muscle actin, NRF2, and F4/80 (Proteintech), with Alexa Fluor– or CoraLite-conjugated secondary antibodies (Abcam, Proteintech). Images (Olympus DP26) were quantified with Fiji/ImageJ (version 2.14.0) and QuPath under consistent settings. Renal IL-1β, IL-6, and IL-10 were measured by ELISA (Elabscience Cat #E-EL-M0044; R&D Systems Cat #VAL601 and #VAL605) and normalized to total protein (BCA assay). RT-qPCR was performed with SYBR Green chemistry on an ABI 7500 system using β-actin as the reference gene and the 2−ΔΔCt method; primer sequences are provided in Table S2.

### Statistical analysis

Analyses used R (version 4.4.1), GraphPad Prism (version 10.4.1), Python (version 3.13.13), and MetaboAnalystR. Data are presented as mean ± SD, mean ± SEM, or median with interquartile range, as specified in the figure legends; tests were two-sided with P < 0.05 considered significant, and the Benjamini– Hochberg procedure controlled the false discovery rate where applicable. Clinical analyses followed randomized allocation and used all available non-missing observations without imputing missing values. Linear mixed-effects models were fitted by restricted maximum likelihood, with participant-specific random intercepts and fixed effects for treatment, elapsed time, and treatment-by-time interaction. The interaction coefficient represents the between-group slope difference (EG minus CG); Wald 95% confidence intervals were calculated from model-based standard errors. Outcome-specific models used 236 observations from 80 participants for Scr, eGFR, and BUN, 200 observations from 79 participants for cystatin C, and 218 observations from 79 participants for PTH. The longitudinal clinical dataset and outcome-specific data availability are provided in Tables S3 and S4. Targeted metabolite, histological, cytokine, and biochemical data were analyzed by one-way ANOVA with Tukey’s test. Spatial metabolomics used PCA/OPLS-DA; zone-matched comparisons used Friedman tests followed by paired Wilcoxon signed-rank tests, while between-group comparisons used Kruskal–Wallis and Mann–Whitney U tests, with Holm correction for multiple comparisons. Individual participants or animals were the biological replicates; repeated assay measurements were not treated as independent biological replicates. Experiment-specific tests, sample sizes, and comparisons are provided in the figure legends. Effect-size distributions and recovery indices were treated as descriptive summaries and not as substitutes for direct between-group significance testing.

## Results

### A defined trace element cocktail establishes the clinical exposure framework

We first characterized exposure to the trace element cocktail in 80 maintenance hemodialysis patients randomized to standard care plus matched placebo (CG, n = 40) or standard care plus TEX (EG, n = 40) and followed longitudinally (Fig. 1A). The baseline characteristics presented were age, sex, and body mass index (Fig. 1B). The formulation comprised 20 mineral elements, with iron, magnesium, aluminum, zinc, potassium, and lithium representing the predominant constituents (Fig. S1 and Table S1). At 20 mL/day, it provided approximately 46.1 mg iron, 39.5 mg magnesium, 29.3 mg aluminum, 14.5 mg zinc, 11.2 mg potassium, 5.14 mg lithium, 2.66 mg manganese, and 463 μg selenium per day; the nominal aluminum exposure corresponded to 205.1 mg/week. Zinc, manganese, copper, and molybdenum exposures remained below general adult upper intake levels, whereas nominal iron and selenium exposures exceeded commonly cited general-population values.[26,27] Aluminum exposure was considered in relation to the general-population tolerable weekly intake framework.[28] These reference values provide general-population context rather than dialysis-specific thresholds. Together with the formulation analysis, they define the administered exposure framework for interpreting the subsequent longitudinal and molecular findings.

**Figure 1.**
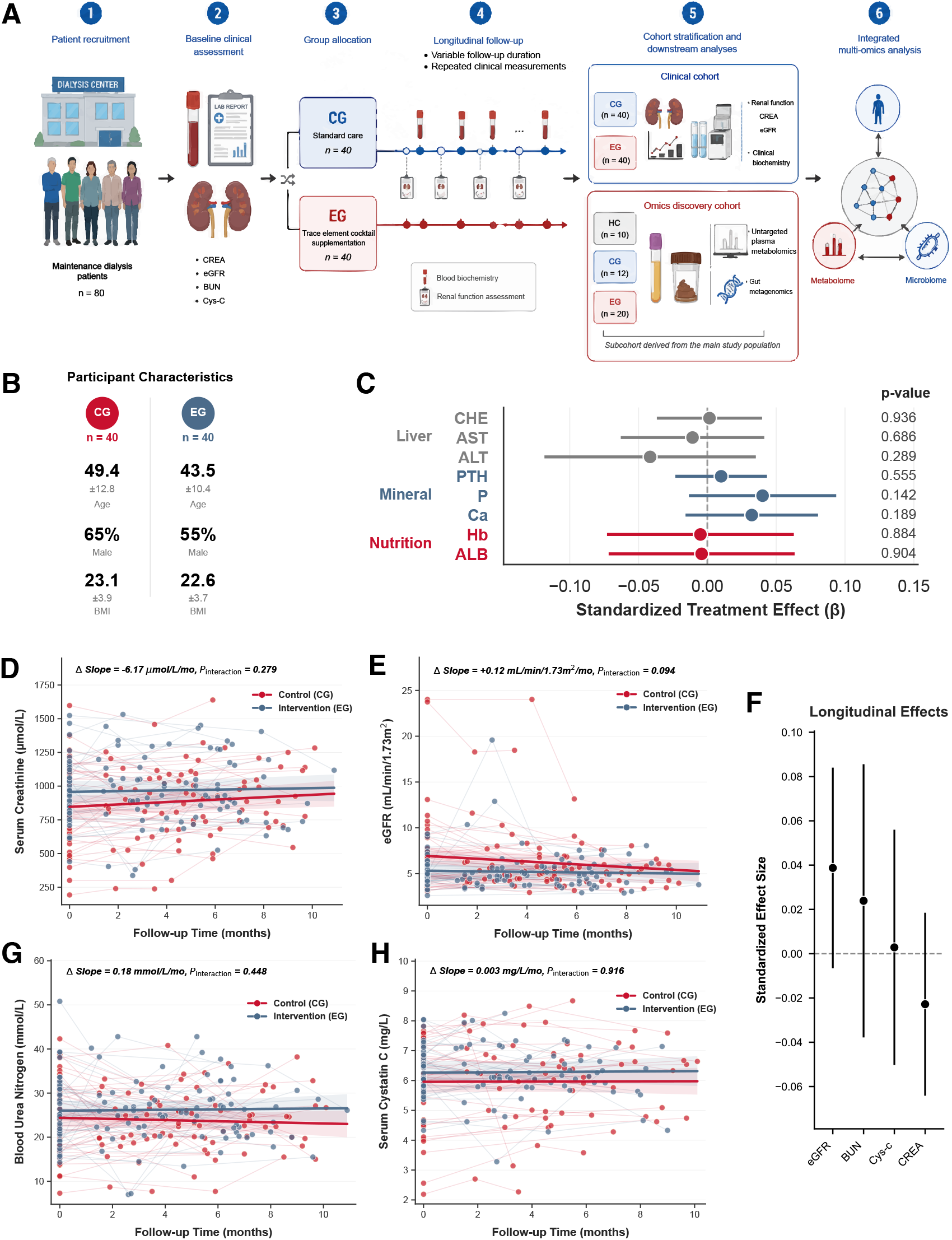
Longitudinal clinical and laboratory monitoring during trace element supplementation. **(A)** Schematic overview of the clinical study design. A total of 80 maintenance hemodialysis patients were randomized to standard care plus matched placebo (CG, n = 40) or standard care plus TEX (EG, n = 40). Endpoint blood and fecal specimens were collected from a voluntary subset with suitable specimens, together with healthy controls (HC). The resulting exploratory multi-omics cohort comprised HC (n = 10), CG (n = 12), and EG (n = 20). **(B)** Baseline demographic characteristics of the clinical cohort, including age, sex, and body mass index (BMI). Age and BMI are presented as mean ± SD; sex is summarized by group. **(C)** Monitored laboratory parameters during follow-up. Forest plots show standardized treatment effects (β coefficients) with 95% confidence intervals for selected nutritional, mineral-metabolic, and hepatic biomarkers. These parameters do not constitute a comprehensive assessment of elemental safety. **(D—E)** Longitudinal trajectories of serum creatinine (CREA) and estimated glomerular filtration rate (eGFR), respectively, during follow-up. **(F)** Summary of longitudinal treatment effects on major renal function indicators, including CREA, eGFR, blood urea nitrogen (BUN), and cystatin C (Cys-C), estimated using linear mixed-effects models. **(G—H)** Longitudinal trajectories of BUN and Cys-C, respectively, during follow-up. Individual points represent observed measurements, solid lines indicate group-level regression trends, and shaded areas represent 95% confidence intervals. For longitudinal analyses, Δ slope represents the difference in the rate of change between EG and CG, and Pinteraction denotes the significance of the time-by-treatment interaction in linear mixed-effects models. Models used 236 observations from 80 participants for creatinine, eGFR, and BUN and 200 observations from 79 participants for cystatin C; outcome-specific availability for all clinical fields is provided in Table S4.

### Longitudinal monitoring establishes the clinical and laboratory context

The clinical dataset comprised 236 observations (CG, 119; EG, 117): 77 participants contributed three records, 2 contributed two records, and 1 contributed baseline data only. The median participant-level maximum recorded follow-up was 6.55 months (IQR 5.48–8.10; range 0–10.9). Longitudinal monitoring showed no overt treatment-related laboratory safety signal. Hepatic biomarkers (cholinesterase, alanine aminotransferase, and aspartate aminotransferase), parathyroid hormone, phosphate, calcium, hemoglobin, and albumin showed no significant treatment-by-time interactions (all Pinteraction ≥ 0.142; Fig. 1C and Fig. S2). Standardized estimates were generally small, with 95% confidence intervals spanning the null (Fig. 1C). These findings support a reassuring laboratory safety profile within the monitored assessments during the observed follow-up.

The serum creatinine slope difference between EG and CG was −6.17 μmol/L/month (95% CI −17.32 to 4.99; Pinteraction = 0.28; Fig. 1D). The corresponding eGFR slope difference was +0.12 mL/min/1.73 m²/month (95% CI −0.02 to 0.27; Pinteraction = 0.094; Fig. 1E). Confidence intervals for the integrated renal-parameter estimates remained compatible with the null (Fig. 1F). Slope differences for BUN and cystatin C were +0.18 mmol/L/month (95% CI −0.28 to 0.63; Pinteraction = 0.45; Fig. 1G) and +0.003 mg/L/month (95% CI −0.060 to 0.066; Pinteraction = 0.92; Fig. 1H), respectively. The PTH slope difference was +3.45 pg/mL/month (95% CI −8.01 to 14.91; Pinteraction = 0.55; Fig. S2D). In maintenance dialysis, these circulating trajectories provide a longitudinal clinical context rather than a direct measurement of residual kidney function, thereby motivating molecular phenotyping of treatment-associated responses.

### Plasma metabolomics identifies treatment-associated metabolic modules

Untargeted plasma metabolomics showed pronounced separation between healthy controls (HC) and CG, consistent with broad CKD-associated metabolic differences,[29-31] whereas EG remained within the CKD-associated metabolic space with a descriptive displacement from CG (999 permutations, P = 0.0010; R²Y = 0.88, Q² = 0.43; Fig. 2A). Median absolute Cohen’s d was 2.09 for CG versus HC and 0.25 for EG versus CG (Fig. 2B). The direct EG-versus-CG comparison identified metabolites in both directions exceeding the FDR = 0.05 threshold (Fig. 2C), and the effect-size distribution indicated selective remodeling across metabolic features. Enrichment analysis of metabolites differing between HC and CG identified multiple interconnected pathways (Fig. 2D), and representative metabolites illustrated heterogeneous endpoint patterns across groups (Fig. 2E). Module-level recovery indices relative to the HC reference were highest for phosphatidylethanolamine and phosphatidylcholine, followed by amino acids (Fig. 2F); selected metabolites likewise showed directional shifts toward the HC reference (Fig. 2G). These human molecular findings identify phospholipid and amino acid modules as priority candidates for longitudinal metabolic studies.

**Figure 2.**
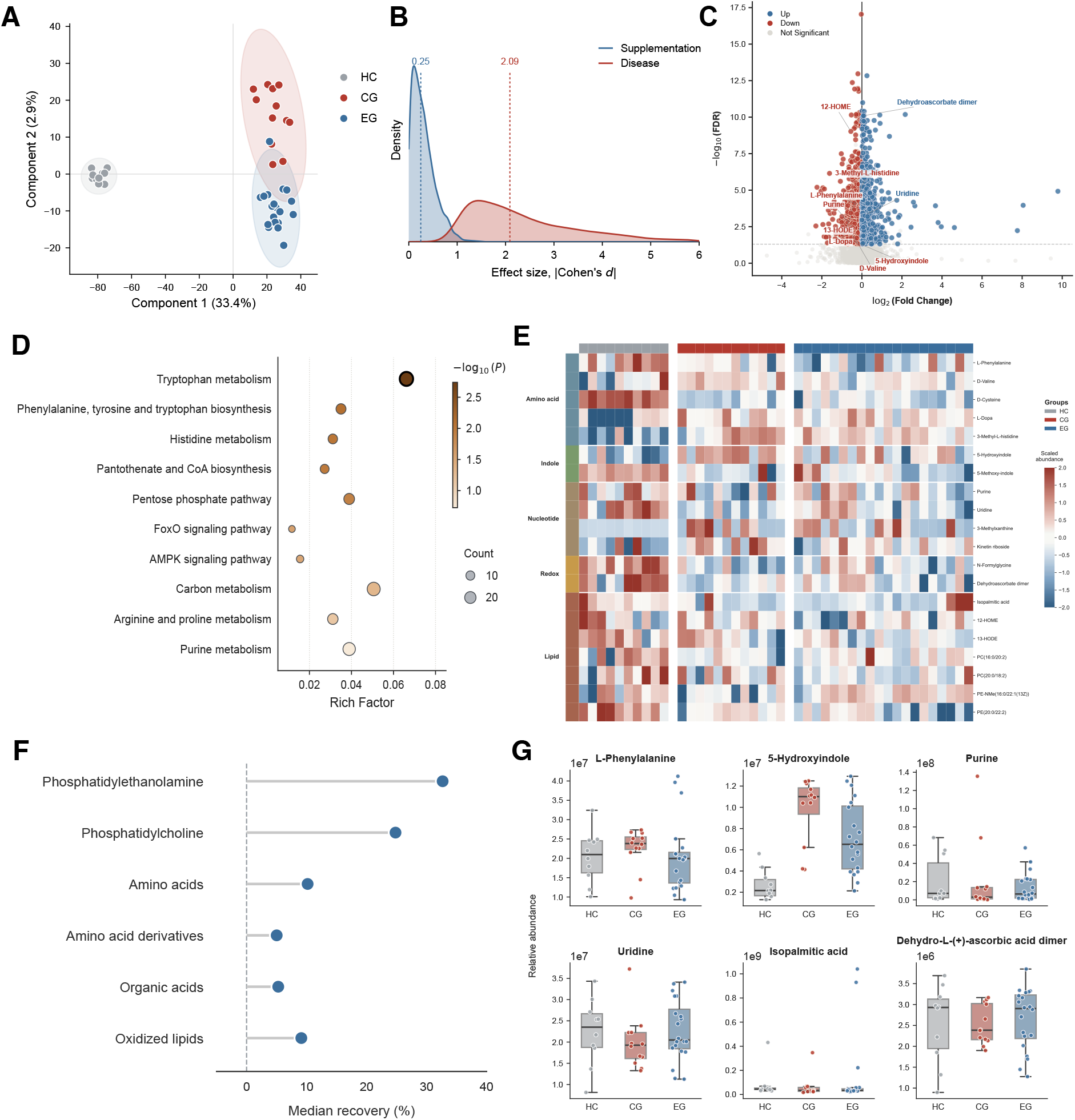
Plasma metabolomic responses to trace element supplementation in CKD. **(A)** Partial least squares discriminant analysis (PLS-DA) of plasma metabolomic profiles from healthy controls (HC, n = 10), CKD controls (CG, n = 12), and trace element-supplemented patients (EG, n = 20). Ellipses indicate 95% Hotelling’s T² confidence regions. Model significance was assessed by 999 label permutations (R²Y = 0.88, Q² = 0.43, P = 0.0010). **(B)** Distribution of metabolite-level effect sizes associated with CKD and trace element supplementation, comparing disease-associated changes (CG vs HC) with supplementation-associated changes (EG vs CG). Dashed lines indicate the median absolute Cohen’s d values. **(C)** Volcano plot of plasma metabolomic changes associated with trace element supplementation (EG vs CG). Blue points denote upregulated metabolites, red points downregulated metabolites, and grey points non-significant metabolites. The dashed line indicates the FDR = 0.05 threshold; points above this line represent metabolites significant after FDR correction. **(D)** KEGG pathway enrichment analysis of CKD-associated metabolites identified by comparison of HC and CG. Dot size represents the number of enriched metabolites and color indicates −log10(p value). **(E)** Heatmap of representative metabolites spanning amino acid, indole, nucleotide, redox, and lipid metabolic modules. Values represent row-wise z scores, and samples are ordered according to the first principal component within each group. **(F)** Descriptive module-level recovery indices following TEX supplementation. For each CKD-associated metabolite, the index was calculated as (EG − CG)/(HC − CG) × 100% and summarized within each metabolic module as the median. Positive values indicate a direction toward the HC reference; these values are descriptive and do not constitute independent confirmatory tests. **(G)** Representative metabolites illustrating CKD-associated metabolic disruption and TEX-associated remodeling toward the HC reference state across HC, CG, and EG groups. Overall and pairwise differences were assessed using two-sided Kruskal–Wallis and Mann–Whitney U tests, respectively. Data are presented as median with interquartile range (IQR); whiskers extend to 1.5× the IQR, and individual samples are shown as points.

### Metagenomics refines the microbial context of metabolic remodeling

Principal coordinate analysis based on Aitchison distance showed separation between HC and CG (PERMANOVA, P < 0.001), whereas EG did not differ significantly from CG (P = 0.14) and remained distinct from HC (P < 0.001; Fig. 3A). Shannon diversity differed across the three groups (P = 0.0072; Fig. 3B). At the taxonomic and functional levels, TEX-associated effect sizes were descriptively smaller than disease-associated differences (Fig. 3C–F). All seven KEGG orthologs in the microbial tryptophan biosynthesis pathway (trpE, trpG, trpD, trpC, trpF, trpA, and trpB) were depleted in CG, whereas their TEX-associated effect sizes were close to the null (Fig. 3G). The composite module score was lower in CG than HC (P < 0.001) and remained lower in EG than HC (P = 0.0050), with no significant CG-versus-EG difference (P = 0.18; Fig. 3H). These findings distinguish community-wide microbial composition from the systemic metabolic response and motivate branch-resolved assessment of host and microbial tryptophan metabolism.

**Figure 3.**
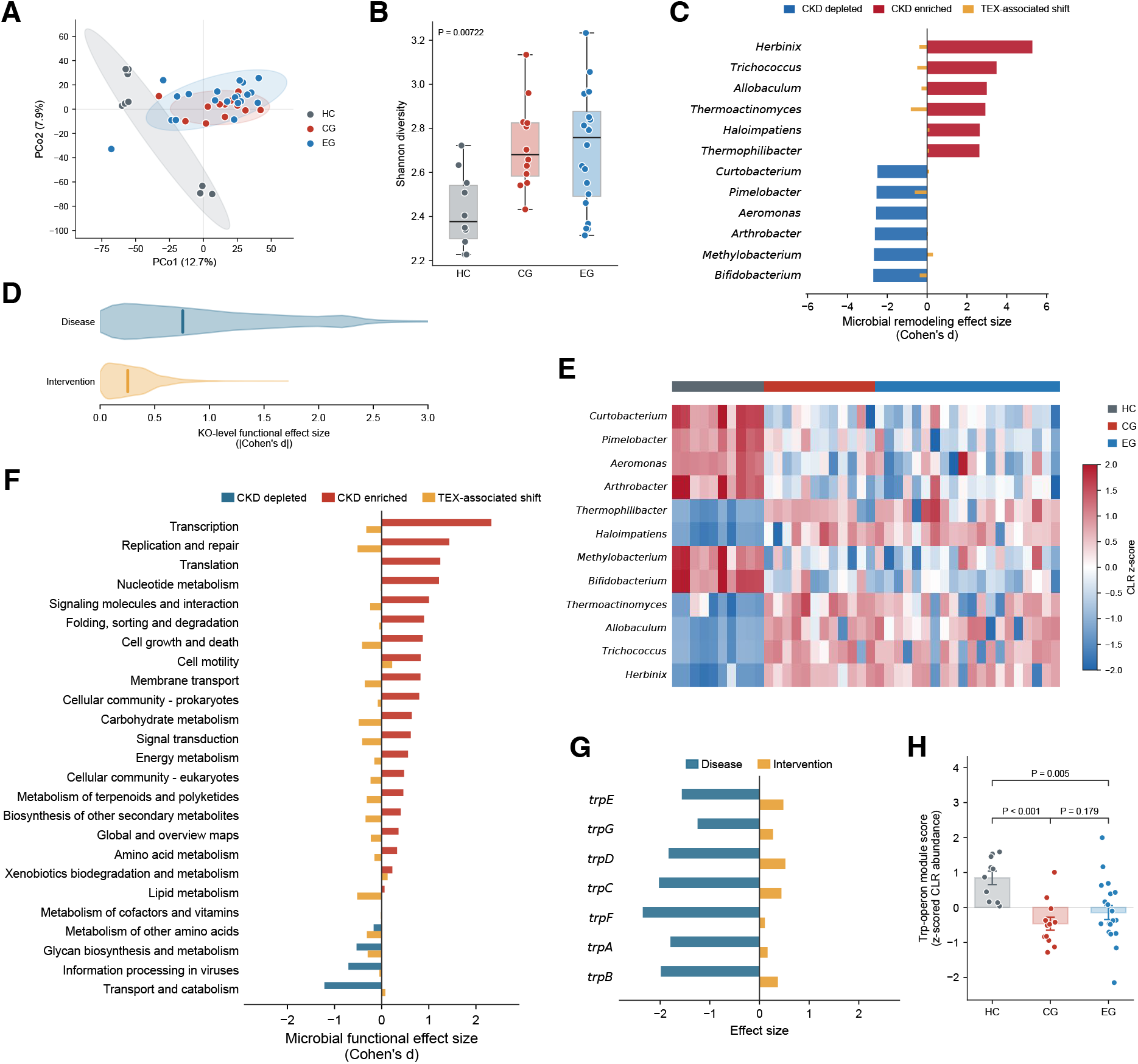
Metagenomic resolution of gut microbial responses to TEX in CKD. **(A)** Principal coordinate analysis (PCoA) of gut microbial community profiles based on Aitchison distance across healthy controls (HC, n = 10), CKD controls (CG, n = 12), and TEX-treated CKD participants (EG, n = 20). **(B)** Shannon diversity of the gut microbiome across the three groups. Each point represents an individual participant. **(C)** Genus-level differential abundance analysis highlighting microbial taxa associated with CKD and TEX supplementation. Bars represent signed Cohen’s d effect sizes, with negative and positive values indicating relative depletion and enrichment, respectively. **(D)** Distribution of KO-level functional effect sizes for CKD-associated and TEX-associated comparisons, illustrating broad functional remodeling in CKD and comparatively limited shifts following TEX supplementation. **(E)** Heatmap of selected differentially abundant microbial genera across the three groups. Values represent standardized CLR-transformed abundances. **(F)** KEGG level-2 functional categories showing CKD-associated depletion and enrichment and TEX-associated functional shifts. **(G)** Effect sizes of the seven KEGG orthologs comprising the microbial tryptophan biosynthesis pathway (trpE, trpG, trpD, trpC, trpF, trpA, and trpB), showing depletion in CKD and limited changes following TEX supplementation. **(H)** Microbial tryptophan biosynthesis module score across the three groups. The module score was calculated as the mean z-scored CLR abundance of the seven tryptophan biosynthesis-related KOs. Statistical analyses were performed using CLR-transformed abundance data with appropriate differential testing and Benjamini–Hochberg false discovery rate correction where applicable. P values are indicated in the corresponding panels.

### TEX selectively modulates host kynurenine metabolism in CKD mice

To resolve the host and microbial components of tryptophan metabolism, we examined the two major branches of circulating tryptophan metabolism, which generate bioactive metabolites implicated in kidney disease, inflammation, and systemic uremic toxicity,[19,32,33] using an independent 5/6 nephrectomy-induced CKD mouse model (Fig. 4A). Targeted profiling identified coordinated CKD-associated changes across both branches (Fig. S3A–C). The host kynurenine branch showed increased KYN/TRP and circulating KYN, 3-hydroxykynurenine (3-HK), kynurenic acid (KYNA), and xanthurenic acid (XA) (Fig. 4B–F). Supplementation shifted this profile toward the sham state, with significant reductions in 3-HK, KYNA, and XA relative to untreated CKD, whereas KYN and KYN/TRP showed similar directional changes without reaching significance. Recovery analysis showed attenuation of CKD-associated elevations in 3-HK, KYNA, and XA (Fig. S3D), and the kynurenine branch metabolite score likewise shifted toward the sham profile (Fig. 4G). The coordinated downstream response was therefore more pronounced than the change in the upstream KYN/TRP ratio, focusing interpretation on metabolite handling within the branch.

**Figure 4.**
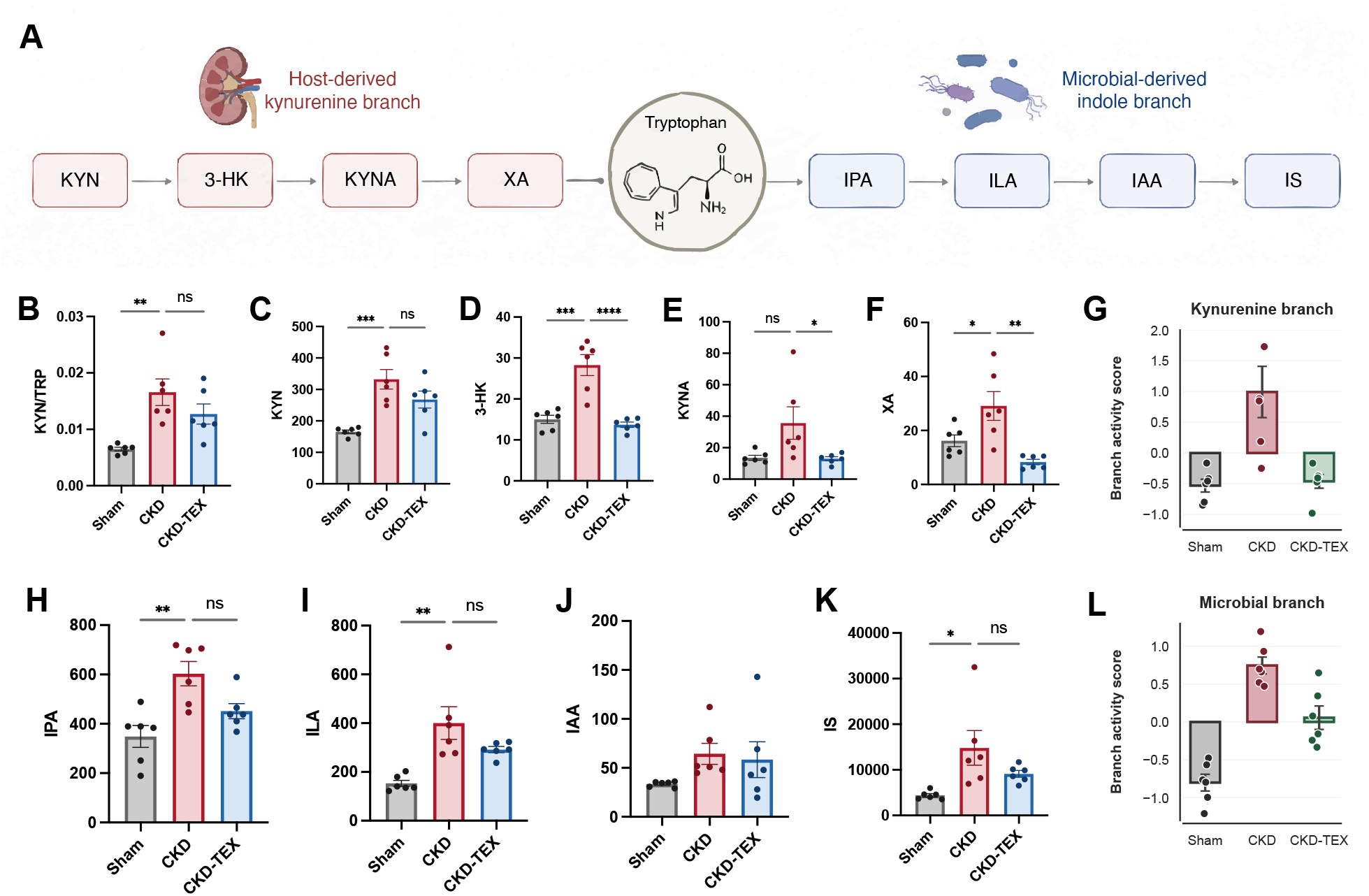
Branch-selective tryptophan metabolic responses to TEX in CKD mice. **(A)** Schematic overview of the two major tryptophan catabolic branches quantified in mouse plasma, comprising the host-derived kynurenine branch and the microbial-derived indole branch. **(B–F)** Plasma levels of kynurenine branch metabolites, including the KYN/TRP ratio, L-kynurenine (KYN), 3-hydroxykynurenine (3-HK), kynurenic acid (KYNA), and xanthurenic acid (XA). **(G)** Kynurenine branch metabolite score calculated as the arithmetic mean of z-score-transformed KYN, 3-HK, KYNA, and XA levels. KYN/TRP was excluded from the score because it represents a ratio derived from two measured metabolites rather than an independent metabolite abundance. **(H–K)** Plasma levels of microbial indole branch metabolites, including indole-3-propionic acid (IPA), indole-3-lactic acid (ILA), indole-3-acetic acid (IAA), and indoxyl sulfate (IS). **(L)** Microbial indole branch metabolite score calculated as the arithmetic mean of z-score-transformed IPA, ILA, and IAA levels. IS was excluded from the score because circulating IS levels are strongly influenced by renal clearance and therefore may not directly reflect microbial pathway activity. n = 6 mice per group. Data are presented as mean ± SEM with individual animals shown. Metabolite data are expressed as normalized relative abundance (arbitrary units, a.u.), reflecting normalized LC-MS/MS signal responses rather than absolute metabolite concentrations. The KYN/TRP ratio was calculated from the corresponding metabolite signals. Statistical significance was determined by one-way ANOVA followed by Tukey’s multiple-comparisons test. Comparisons shown include Sham versus CKD and CKD versus CKD-TEX. *P < 0.05, **P < 0.01, ***P < 0.001, ****P < 0.0001; ns, not significant.

In contrast, metabolites within the microbial indole branch showed less consistent responses. CKD increased circulating indole-3-propionic acid (IPA) and indole-3-lactic acid (ILA), but neither was significantly reduced following supplementation, while indole-3-acetic acid (IAA) showed no clear treatment-associated change (Fig. 4H–J). Indoxyl sulfate was elevated in CKD, but the CKD-versus-CKD-TEX comparison was not significant (Fig. 4K). Because circulating indoxyl sulfate is strongly influenced by renal clearance, it was not interpreted as a direct indicator of microbial pathway activity.[19,34] Consistently, the composite indole branch metabolite score showed no clear restoration with TEX (Fig. 4L). The response in mice was therefore branch selective, with movement toward the sham phenotype in the host kynurenine arm. Together with the human metagenomic findings, these observations prioritize host-associated kynurenine metabolism for mechanistic investigation of TEX-associated renal responses.

### Spatial metabolomics resolves region-specific renal responses to TEX

MALDI imaging mass spectrometry-based spatial metabolomics was performed on kidney sections from sham and CKD mice, with four anatomically defined regions—the cortex, outer stripe of the outer medulla (OSOM), inner stripe of the outer medulla (ISOM), and inner medulla (IM)—analyzed separately (Fig. 5A). Principal component analysis revealed pronounced spatial structuring, consistent with the established regional organization of renal metabolism,[22,35,36] and CKD produced substantial displacement of these regional profiles (Fig. 5B). Regional deviation was heterogeneous across zones (Friedman test, P = 0.00090), with the inner medulla showing the greatest deviation (Fig. 5C). The global measure of renal metabolic zonation showed an overall group difference (Kruskal–Wallis test, P = 0.042), but no pairwise comparison remained significant after Holm correction, including CKD versus CKD-TEX (Fig. 5D). Representative metabolites displayed distinct zone-enriched distributions under physiological conditions—triglyceride species in the cortex, kynurenine in the OSOM, acetyl-L-carnitine in the ISOM, and glycerophosphocholine in the IM—with corresponding ion images supporting their regional distributions and CKD-associated remodeling (Fig. 5E– G).

**Figure 5.**
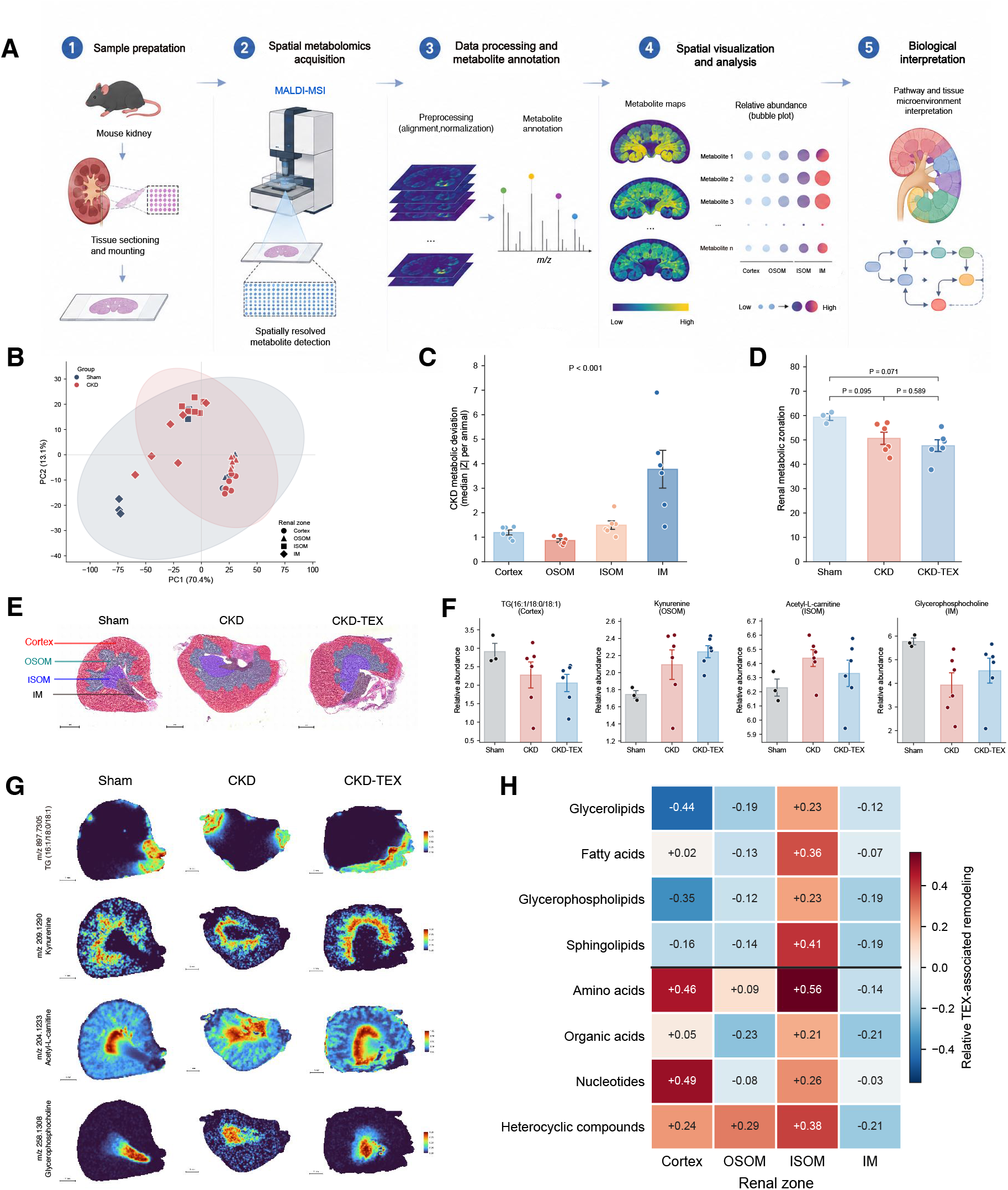
Region-specific renal metabolic responses to TEX in CKD mice. **(A)** Schematic overview of the MALDI imaging mass spectrometry (MALDI-MSI) spatial metabolomics workflow and analysis of four renal anatomical zones: cortex, outer stripe of the outer medulla (OSOM), inner stripe of the outer medulla (ISOM), and inner medulla (IM). **(B)** Principal component analysis (PCA) of spatial metabolic profiles from Sham and CKD kidneys. Each point represents a renal zone from an individual animal; ellipses indicate 95% confidence regions. **(C)** Regional metabolic deviation in CKD, quantified as the median absolute Z score relative to the corresponding Sham zone. IM showed the greatest metabolic deviation across renal zones. **(D)** Quantification of renal metabolic zonation based on the mean pairwise distance among the four regional metabolic profiles within each animal. **(E)** Representative H&E-stained kidney sections illustrating the four anatomical zones analyzed by spatial metabolomics. Scale bars, 1 mm. **(F)** Relative abundance of representative zone-enriched metabolites, including TG(16:1/18:0/18:1), kynurenine, acetyl-L-carnitine, and glycerophosphocholine. **(G)** Representative ion distribution maps of the metabolites shown in (F). Scale bars, 1 mm. **(H)** Relative TEX-associated metabolic remodeling across renal zones and metabolic classes. Values represent median metabolite-level Cohen’s d for CKD-TEX versus CKD, centered to the global median. Sham, n = 3; CKD, n = 6; CKD-TEX, n = 6 animals. Data are presented as mean ± SEM unless otherwise indicated. For (C), regional metabolic deviation across renal zones was evaluated using the Friedman test, followed by paired Wilcoxon signed-rank tests with Holm correction. For (D) and (F), group differences were assessed using Kruskal–Wallis tests followed by two-sided Mann–Whitney U tests with Holm correction. For (H), values represent median metabolite-level Cohen’s d for CKD-TEX versus CKD, centered to the global median; no significance testing was performed.

Descriptive TEX-associated effect-size patterns varied by anatomical location and metabolic class rather than showing a uniform shift (Fig. 5H). After centering to the global median, all displayed metabolic classes had positive values in ISOM and negative values in IM, whereas the cortex and OSOM showed class-dependent mixtures of positive and negative values. No significance testing was performed for these centered effect-size patterns. Together, the regional deviation and centered effect-size maps distinguish the anatomy of CKD-associated disruption from the anatomy of TEX-associated responsiveness. These findings define regional heterogeneity as a central feature of renal metabolic remodeling and identify anatomical compartments for focused tissue analyses.

### TEX engages selected renal structural and tubular markers

To place the regional metabolic patterns in a tissue context, we examined selected renal structural and tubular markers (Fig. 6A). PAS staining showed prominent tubular injury in CKD kidneys (Fig. 6B). CKD increased collagen I deposition and α-SMA-positive area; TEX significantly reduced collagen I accumulation but not α-SMA area (Fig. 6E and F).[30] AQP1 fluorescence intensity was reduced in CKD kidneys and was significantly higher following TEX supplementation (Fig. 6E and F). Endpoint serum creatinine and BUN did not differ significantly between CKD and CKD-TEX, and the CKD-versus-CKD-TEX difference in tubular injury score was not significant (Fig. 6C and D). The principal tissue signal was therefore concentrated in selected structural and tubular markers, providing phenotypic context for the metabolic findings.

**Figure 6.**
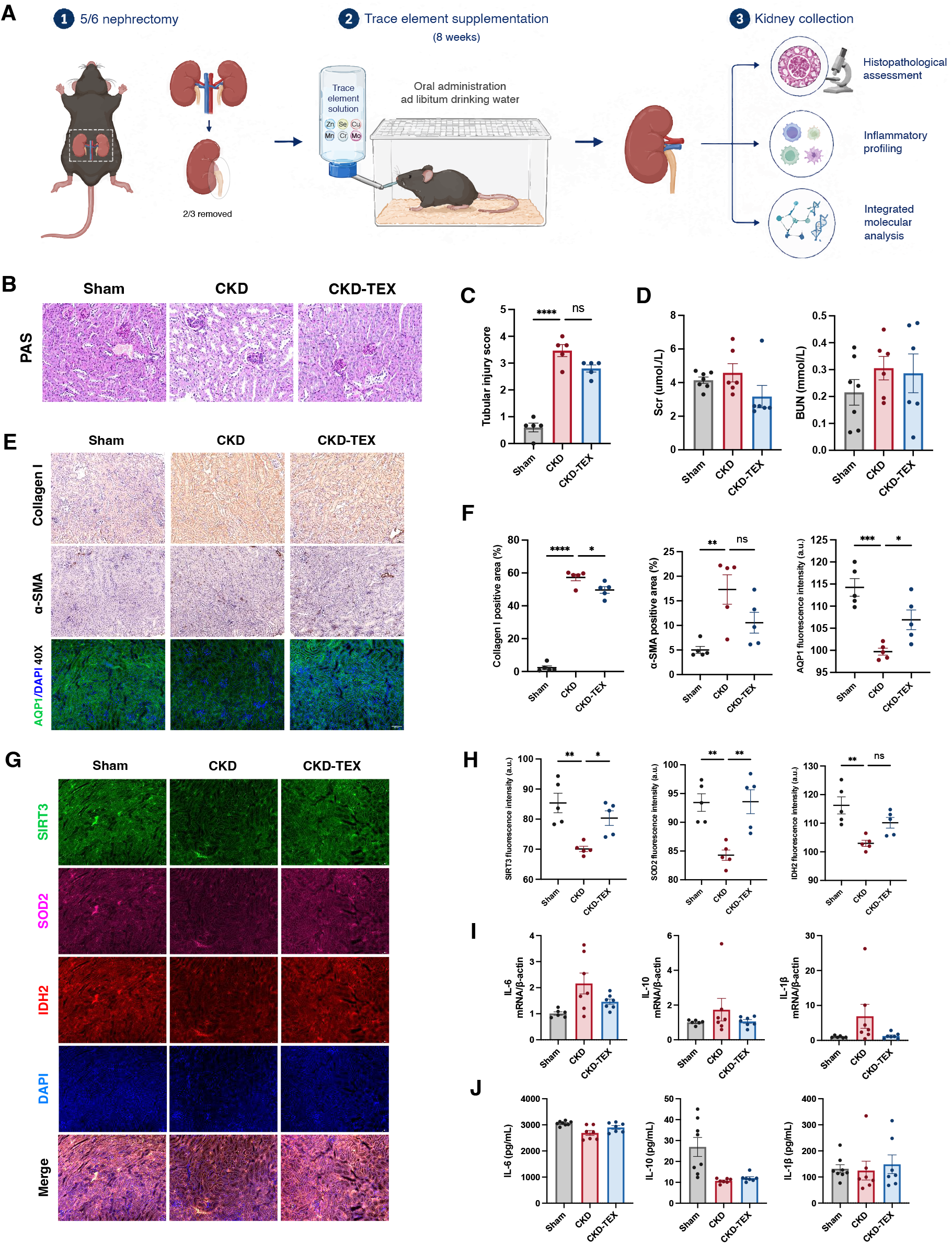
Selective renal structural and antioxidant-related responses to TEX in CKD mice. **(A)** Experimental workflow of the 5/6 nephrectomy-induced CKD mouse model. Following nephrectomy, mice received oral trace element supplementation through ad libitum drinking water for 8 weeks before kidney collection for histopathological, inflammatory, and molecular analyses. **(B)** Representative PAS-stained kidney sections from Sham, CKD, and CKD-TEX mice. **(C)** Quantification of tubular injury score in kidney sections (n = 5 per group). **(D)** Serum creatinine (Scr) and blood urea nitrogen (BUN; n = 6–7 per group). **(E)** Representative kidney sections showing Collagen I and α-SMA immunohistochemical staining and AQP1 immunofluorescence staining. **(F)** Quantification of collagen I-positive area, α-SMA-positive area, and AQP1 fluorescence intensity (n = 5 per group). **(G)** Representative renal immunofluorescence images of SIRT3 (green), SOD2 (magenta), IDH2 (red), and DAPI (blue), with corresponding merged images. **(H)** Quantification of SIRT3, SOD2, and IDH2 fluorescence intensity (n = 5 per group). **(I)** Renal mRNA expression of IL-6, IL-10, and IL-1β measured by RT-qPCR (n = 6–7 per group). **(J)** Renal protein levels of IL-6, IL-10, and IL-1β measured by ELISA (n = 7–8 per group). Data are presented as mean ± SEM with individual biological replicates shown. Statistical comparisons were performed using ordinary one-way ANOVA followed by Tukey’s multiple-comparisons test. *P < 0.05, **P < 0.01, ***P < 0.001, ****P < 0.0001; ns, not significant. Scale bars, 50 μm.

Exploratory interaction analyses further examined whether circulating kynurenine-branch metabolites showed group-dependent associations with BUN. Significant metabolite-by-group interactions were observed for 3-HK and KYNA (Pinteraction = 0.026 and 0.0050, respectively; Fig. S3E). These group-dependent associations provide a quantitative link between branch metabolites and renal phenotypic variation, nominating kynurenine-branch metabolites for controlled studies of metabolic–renal phenotype coupling.

### TEX enhances selected antioxidant and stress-response markers

CKD was associated with marked reductions in renal SIRT3 and SOD2 fluorescence intensity, together with a more modest reduction in IDH2, consistent with alteration of mitochondrial antioxidant-related markers.[37,38] Representative images are shown in Fig. 6G and quantification in Fig. 6H. TEX significantly increased SIRT3 and SOD2 signals relative to untreated CKD, whereas IDH2 showed a directional but non-significant difference. NRF2 fluorescence intensity was significantly higher following supplementation than in untreated CKD, while CKD alone did not differ significantly from sham (Fig. S4A and B).[39] Together, these changes identify a selective antioxidant-related molecular response centered on SIRT3, SOD2, and NRF2, providing a tissue-level complement to the metabolic remodeling observed in CKD mice.

SIRT1, CD38, and F4/80 showed no treatment-associated differences: although SIRT1 and CD38 were altered in CKD relative to sham, neither differed significantly between CKD and CKD-TEX (Fig. S4C–E), and F4/80-positive area did not differ across groups (Fig. S4F and G). Renal IL-6, IL-10, and IL-1β transcript levels and corresponding cytokine concentrations likewise showed no consistent treatment-associated changes (Fig. 6I and J). This marker specificity reinforces the selective nature of the tissue response and narrows the molecular model to a defined mitochondrial/redox module rather than a uniform change across all measured pathways.

## Discussion

This study identifies branch-selective and region-specific metabolic remodeling as the central feature of TEX-associated responses in CKD. The clinical cohort establishes the human context, with longitudinal laboratory monitoring showing no overt treatment-related safety signal and plasma metabolomics resolving treatment-associated molecular differences. The mouse experiments then progressively refine this signal: targeted profiling identifies the host kynurenine branch as a responsive pathway, spatial metabolomics maps renal responses to anatomical compartments, and tissue analyses reveal accompanying structural and antioxidant-related phenotypes. The resulting evidence chain moves from systemic metabolic differences to pathway selectivity, renal localization, and tissue context, providing a coherent model of how a multi-element intervention engages metabolic organization in CKD. Because creatinine and eGFR trajectories do not directly quantify residual kidney function in maintenance dialysis,[40,41] molecular phenotyping adds a complementary dimension to conventional clinical assessment.

The formulation provides a biologically plausible basis for this selective response. Essential trace elements participate in redox regulation, mitochondrial and intermediary metabolism, and cellular homeostasis, while their availability in CKD is influenced by absorption, tissue distribution, renal handling, and dialysis-associated removal.[7,9,42] The formulation also contains elements not classified as essential trace elements. Previous clinical studies have generally evaluated individual elements and reported heterogeneous biochemical and clinical responses.[10-13] The present findings extend this work by evaluating a defined multi-element formulation as a coordinated exposure whose responses can be resolved across metabolic branches and tissue compartments. Experimental evidence linking selenoprotein H to mitochondrial quality control illustrates how elemental biology can be translated into specific renal mechanisms,[14] while the present systems-level phenotype provides a basis for future component-resolved analysis. This shifts the emphasis from single-element endpoints toward the organization and selectivity of the metabolic response.

Within the clinical cohort, plasma metabolomics provides the strongest human molecular evidence of treatment-associated differences. CKD is characterized by extensive disruption of circulating metabolic networks,[5,43-45] and the healthy-control and CKD comparisons reproduced that broad metabolic background. Against this background, the EG-versus-CG comparison identified FDR-significant metabolites in both directions, with selected phospholipid and amino acid modules and individual metabolites shifting toward the healthy reference. The smaller treatment-associated effect-size distribution relative to the disease-associated comparison further supports selectivity: the response is concentrated in defined modules rather than dispersed uniformly across the metabolome. Phosphatidylethanolamine and phosphatidylcholine recovery also connects the intervention-associated signature to membrane and lipoprotein-related lipid organization. Detailed HDL lipidomics in CKD has shown that molecular lipid species provide pathophysiological information beyond conventional lipid measurements,[46] supporting the value of resolving lipid responses at the species and module levels. Recovery indices quantify the direction and magnitude of these shifts and provide a practical framework for evaluating their temporal durability in baseline-controlled studies of metabolic resilience.[47] Accordingly, the clinical metabolomic signature serves as a pathway-selection layer for the mechanistic analyses that follow.

The fecal metagenomic findings refine this model by distinguishing microbial community structure from metabolic pathway responses. CKD markedly altered microbial composition and tryptophan biosynthetic potential, whereas TEX-associated community-level effects were more circumscribed. This pattern directs attention to selective microbial functions and host metabolic handling, rather than requiring broad community restructuring as an explanation for systemic metabolic differences. Fecal metagenomics captures genetic and functional potential, and its integration with microbial activity, substrate availability, metabolite flux, and localized intestinal interactions will further resolve the gut–kidney metabolic interface.[48-51] Integrated human metagenomic and metabolomic studies have linked gut microbial and circulating metabolic variation to CKD severity,[52] while defined microbial pathways can generate circulating indole metabolites with host biological activity.[17,20] In hemodialysis, gut microbial features have been linked to indole metabolism and circulating uremic solutes, while kidney function itself can influence microbial metabolic output.[53,54] Accordingly, the metagenomic results serve as a mechanistic filter that motivates branch-resolved analysis of tryptophan metabolism. A useful mechanistic comparison is provided by a JAR study in which microbial IPA alleviated diabetic kidney disease through mitochondrial protection involving SIRT1 stabilization.[55] The present data show a different response architecture: the indole-branch score and SIRT1 were not treatment-responsive, whereas host kynurenine metabolites and SIRT3, SOD2, and NRF2 markers changed in a coordinated manner. This contrast narrows the model from a generalized microbial metabolite–sirtuin explanation to a branch- and node-specific framework.

The host kynurenine branch is closely linked to inflammatory and metabolic features of CKD.[33,56] Kynurenines are bioactive regulators of immune and metabolic processes,[21] and kynurenine-3-monooxygenase represents a regulated step connecting upstream kynurenine handling to 3-HK and downstream metabolism.[57] Within this pathway, targeted mouse analysis identifies the clearest pathway-level response to TEX. Reductions in 3-HK, KYNA, and XA, together with the shift in the kynurenine-branch metabolite score toward the sham profile, form a coherent response across related metabolites. The different behavior of microbial indole metabolites reinforces this branch selectivity. Host kynurenine metabolism and microbial indole production are interconnected but distinct arms of tryptophan metabolism in CKD,[18,19,58] and kynurenine-pathway metabolites have been associated with inflammation, renal dysfunction, and subsequent kidney failure.[58-60] Recent mechanistic studies of quinolinic acid further demonstrate the value of connecting kynurenine-pathway metabolites with renal spatial metabolic phenotypes and systemic outcomes.[61,62] Against this broader pathway context, the coordinated 3-HK, KYNA, and XA responses narrow the TEX-associated signature to specific downstream metabolites. Their stronger response relative to KYN and KYN/TRP raises the possibility that downstream production, utilization, or transport may be particularly relevant, while the group-dependent associations of 3-HK and KYNA with BUN add phenotypic relevance. These alternatives can now be distinguished directly using isotope-resolved flux analysis together with enzyme- and transport-focused measurements.[63]

Renal spatial metabolomics localizes this pathway-selective response within the kidney. CKD-associated disruption was anatomically heterogeneous, with the inner medulla showing the greatest deviation from sham. Kynurenine itself displayed a zone-enriched distribution in the OSOM, while TEX-associated effect sizes varied across regions and metabolic classes. Spatial metabolomics is specifically suited to resolving such regional alterations, which can be diluted when kidney tissue is analyzed as a single homogenate.[24,64-67] Human studies combining spatial metabolomics with clinical and molecular phenotyping have likewise demonstrated regional differences in renal oxidative metabolism,[36] reinforcing the importance of anatomical context. The observation that the IM showed the greatest CKD-associated deviation while centered TEX-associated class effects differed across the ISOM and IM further distinguishes the spatial distribution of disease disruption from that of treatment responsiveness. The maps therefore convert whole-organ metabolic averages into anatomically resolved phenotypes and provide spatial coordinates for examining how circulating pathway responses relate to renal tissue metabolism. Global zonation and regional analyses offer complementary views of renal metabolic organization,[68,69] with the regional profiles identifying priority compartments for mechanistic investigation in the context of their distinctive oxygen and substrate environments.[23]

The tissue analyses add structural and molecular significance to this spatially resolved model. Reduced collagen I accumulation and increased AQP1 signals identify extracellular-matrix and tubular marker responses, while increased SIRT3, SOD2, and NRF2 signals support a mitochondrial and antioxidant-related component. AQP1 has an established role in tubular water handling,[70] making its increased signal a useful tubular phenotypic correlate for future transport-focused validation. Tubular cells have substantial energetic requirements and depend heavily on mitochondrial metabolism,[37,38,71] and SIRT3 integrates metabolic and oxidative signals relevant to tubular adaptation.[72] Mitochondrial ROS can function as both signals and amplifiers of cellular injury,[73] providing a biologically coherent point of convergence for the observed metabolic and redox-related responses. Recent JAR studies have shown how selenoprotein H–MTCH2–MFN2 signaling and ANXA1–UCP1 signaling can be experimentally resolved into defined mitochondrial quality-control and homeostatic mechanisms in kidney models.[14,74] The present findings nominate a complementary SIRT3–SOD2–NRF2-associated axis for this level of mechanistic analysis. The convergence of kynurenine-branch remodeling, region-specific metabolic responses, and antioxidant-related marker changes therefore defines a kynurenine–mitochondrial/redox framework as a testable mechanism linking systemic and renal responses, with collagen I and AQP1 extending the framework to extracellular-matrix and tubular phenotypes.

Across these complementary experimental layers, each analysis resolves a question raised by the preceding one, consistent with a problem-oriented rather than platform-oriented use of multi-omics.[25] Human metabolomics establishes systemic treatment-associated differences; metagenomics clarifies the microbial context; targeted profiling identifies branch selectivity; spatial metabolomics provides renal localization; and tissue analyses add phenotypic context. Human and mouse findings are complementary rather than interchangeable: the former anchor clinical relevance, while the latter provide pathway, regional, and tissue resolution. This evidence chain creates a coherent and experimentally tractable model with three explicit predictions. First, if TEX preferentially alters host kynurenine handling, isotope-resolved studies should detect branch-specific changes in metabolite flux rather than relying on abundance alone.[63] Second, region- and cell-resolved analyses should determine whether kynurenine metabolism and mitochondrial/redox markers coincide within defined renal compartments; single-cell metabolomics and integrated kidney spatial studies provide directly relevant methodological precedents.[75,76] Third, component-resolved formulations coupled with blood and tissue element measurements should establish which exposure features account for the response. Together, these predictions convert the kynurenine–mitochondrial/redox model into a practical research program with defined readouts and experimental endpoints.

These findings also define a focused agenda for confirmation and translation. The human omics analyses were conducted at endpoint in voluntary, unequally sized subsets without baseline molecular measurements, and one participant contributed baseline clinical data only; baseline-controlled longitudinal multi-omics in adequately powered cohorts will therefore be important for establishing temporal persistence and clinical generalizability.[77] Urine-based measures of residual kidney function, direct blood and tissue element measurements, absorption and retention analyses, and component-resolved formulations will further refine clinical and exposure attribution. Participant-level adverse-event counts and returned-packaging adherence records were unavailable for quantitative analysis, and general-population intake reference values are not dialysis-specific safety thresholds; future studies should integrate systematic harms and adherence reporting with direct exposure assessment. In mice, larger groups, individual dose assessment, isotope tracing, and cell-resolved kidney analyses will strengthen causal and mechanistic resolution.[78]

The trial was registered retrospectively after completion of recruitment and follow-up, and post-initiation changes were made to the outcome panel and classification. These factors limit assessment of complete prospective alignment and are therefore disclosed explicitly. Ethics approval preceded enrollment but did not constitute prospective trial registration.

## Conclusion

In conclusion, TEX-associated metabolic remodeling in CKD is defined by branch selectivity, renal regional specificity, and selective tissue involvement. Human circulating phospholipid and amino acid modules provide the clinical anchor, while the independent CKD mouse experiments resolve host kynurenine-branch remodeling, anatomically patterned renal metabolic responses, and coordinated collagen I/AQP1 and SIRT3/SOD2/NRF2 phenotypes. These findings establish a specific response architecture—kynurenine-branch remodeling accompanied by spatially organized renal metabolic, mitochondrial/redox, and structural changes—rather than a generalized metabolic or microbial signature. The kynurenine–mitochondrial/redox connection is thus the central mechanistic proposition emerging from this study.

## Data Availability

De-identified individual participant data underlying the longitudinal clinical analyses, together with a data
dictionary and outcome-specific data availability, are provided in Tables S3 and S4 and are available with
the article from the date of publication. These data include randomized group, demographic variables, visit
time, and reported laboratory measurements; direct identifiers are not included. They may be used to verify
the reported analyses and for secondary research subject to applicable ethical and institutional requirements.
Metagenomic sequencing data are available in the NCBI Sequence Read Archive (PRJNA1517233;
https://www.ncbi.nlm.nih.gov/bioproject/PRJNA1517233), untargeted and targeted metabolomics data in
Page 12
MetaboLights (MTBLS15494 and MTBLS15794; https://www.ebi.ac.uk/metabolights/MTBLS15494 and
https://www.ebi.ac.uk/metabolights/MTBLS15794), and spatial metabolomics data in METASPACE
(project 0f0d8880-9fbb-11f1-a04c-93788e0a92d9; https://metaspace2020.eu/project/0f0d8880-9fbb-11f1-a04c-93788e0a92d9). Other study materials are available from the corresponding author upon reasonable
request for verification or research use, subject to institutional and ethics approval and a data-use agreement
where required.

## Data sharing statement

De-identified individual participant data underlying the longitudinal clinical analyses, together with a data dictionary and outcome-specific data availability, are provided in Tables S3 and S4 and are available with the article from the date of publication. These data include randomized group, demographic variables, visit time, and reported laboratory measurements; direct identifiers are not included. They may be used to verify the reported analyses and for secondary research subject to applicable ethical and institutional requirements. Metagenomic sequencing data are available in the NCBI Sequence Read Archive (PRJNA1517233; https://www.ncbi.nlm.nih.gov/bioproject/PRJNA1517233), untargeted and targeted metabolomics data in MetaboLights (MTBLS15494 and MTBLS15794; https://www.ebi.ac.uk/metabolights/MTBLS15494 and https://www.ebi.ac.uk/metabolights/MTBLS15794), and spatial metabolomics data in METASPACE (project 0f0d8880-9fbb-11f1-a04c-93788e0a92d9; https://metaspace2020.eu/project/0f0d8880-9fbb-11f1-a04c-93788e0a92d9). Other study materials are available from the corresponding author upon reasonable request for verification or research use, subject to institutional and ethics approval and a data-use agreement where required.

## Declaration of generative AI and AI-assisted technologies in the manuscript preparation process

During the preparation of this work, the authors used ChatGPT (OpenAI) and Kimi K3.1 (Moonshot AI) to polish the language and improve readability, and to assist with manuscript formatting and layout adjustments. After using these tools, the authors reviewed and edited the content as needed and take full responsibility for the content and integrity of the publication.

## Acknowledgments

The authors thank all individuals involved in this study and acknowledge Wuhan Maiwei Metabolism Biotechnology Co., Ltd. for providing metabolomics, microbiome sequencing, and related analytical services.

## CREDIT Author Statement

Jianyu Piao: Writing – original draft, Visualization, Investigation, Formal analysis. Mingji Jin: Supervision, Funding acquisition. Fengxian Yin: Supervision, Investigation, Data curation. Zhenyu Quan: Formal analysis. Zhonggao Gao: Writing – review & editing, Supervision, Validation, Methodology, Funding acquisition, Conceptualization. Zhenhua Cui: Writing – review & editing, Supervision, Validation, Methodology, Funding acquisition, Conceptualization.

