## Supplementary information for "Multi-omics reveals selective metabolic remodeling with trace element supplementation in chronic kidney disease"

#### Trace element supplementation and selective metabolic remodelling in chronic kidney disease: a randomised controlled trial with exploratory multi-omics and mouse experiments

Jianyu Piao, Mingji Jin, Fengxian Yin, Zhenyu Quan, Zhonggao Gao, Zhenhua Cui

##### Contents

Supplementary Figure 1. Composition and elemental concentrations 2

Supplementary Figure 2. Longitudinal laboratory monitoring 3

Supplementary Figure 3. Targeted tryptophan metabolite profiles 4

Supplementary Figure 4. Renal stress-response and inflammatory markers 5

Table S1. Elemental composition, ICP-OES conditions and replicate measurements 6

Table S2. Primer sequences 9

Table S3A–F. De-identified longitudinal clinical dataset 10

Table S4. Clinical data dictionary and availability 40

All supplementary text is provided in English. Blank cells in Table S3 denote missing values in the source workbook; no imputation was applied.

**Supplementary Figure 1. Composition and elemental concentrations of the trace element cocktail**

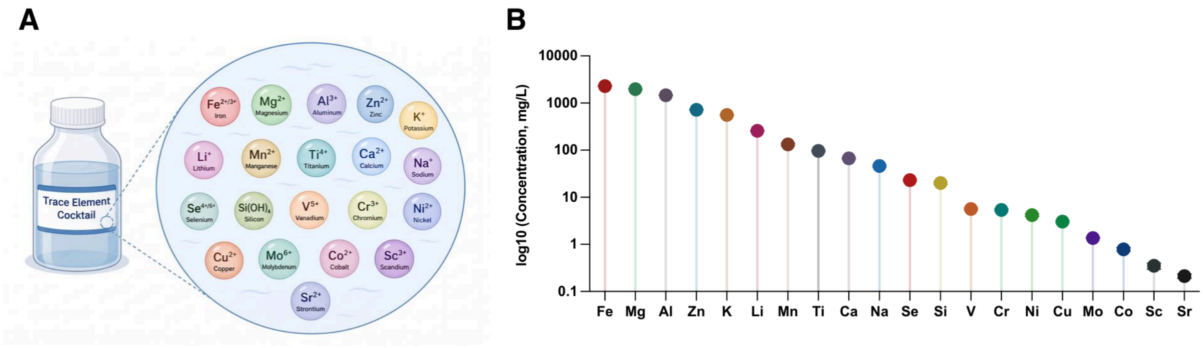


**(A)** Schematic illustration of the trace element cocktail (TEX) showing its constituent mineral elements.

**(B)** Absolute concentrations of individual elements determined by inductively coupled plasma optical emission spectrometry (ICP-OES; Agilent 5800). Concentrations are presented on a $\log_{10}$ scale and expressed as mg/L.

Quantitative measurements were obtained from three independent digestions of the same composite solution (2 mL digested and diluted to a final volume of 10 mL; n = 3). Fe, Mg, Al, Zn, K, and Li received an additional 20-fold dilution; the other elements received no additional dilution. Raw data, operating conditions, means, standard deviations, and coefficients of variation are provided in Table S1.

### Supplementary Figure 2. Longitudinal monitoring of selected laboratory parameters during trace element supplementation

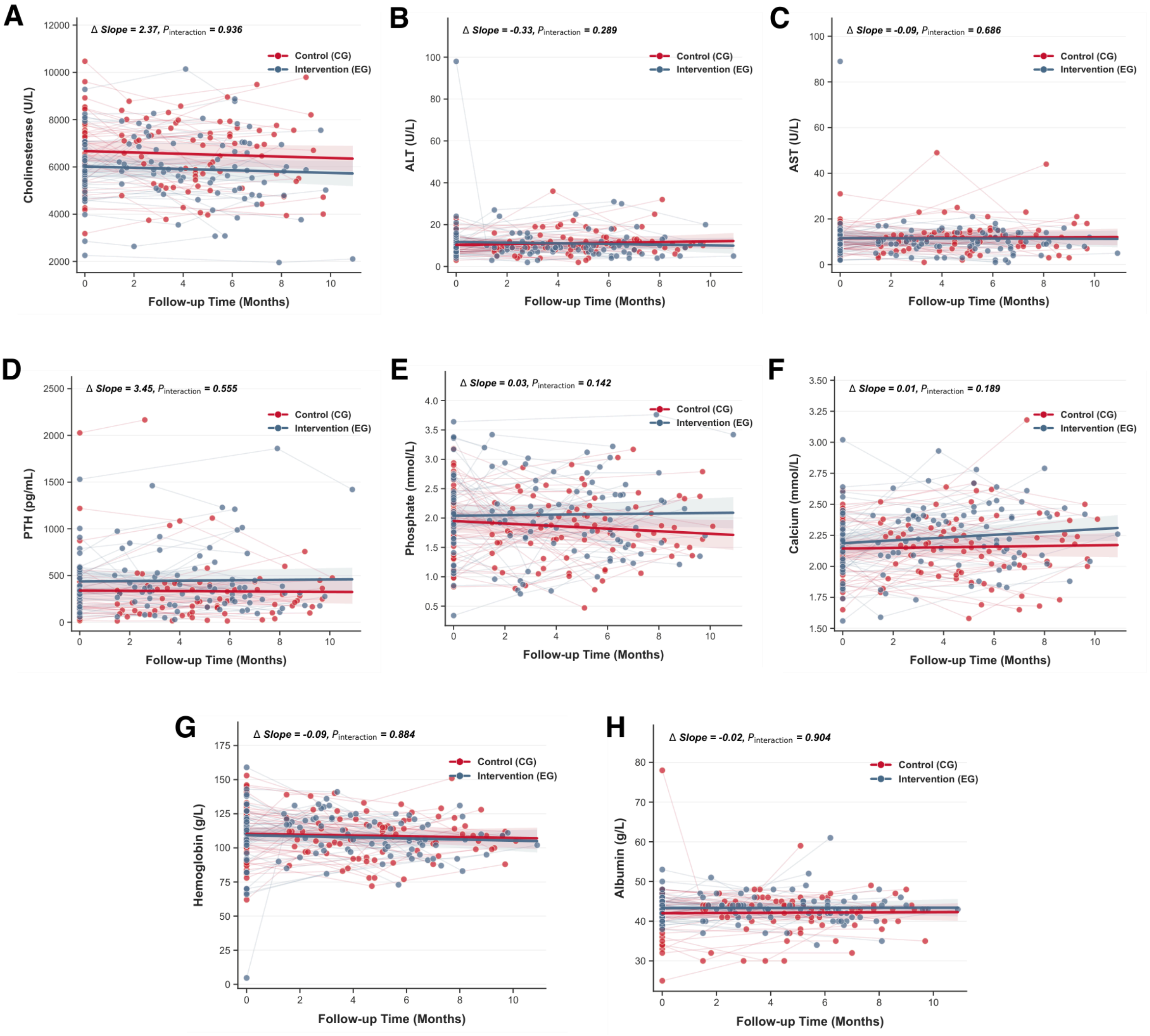


**(A–H)** Longitudinal trajectories of cholinesterase (CHE), alanine aminotransferase (ALT), aspartate aminotransferase (AST), parathyroid hormone (PTH), phosphate, calcium, haemoglobin (Hb), and albumin (ALB), respectively, in the control group (CG) and experimental group (EG). Individual points represent observed measurements, solid lines indicate group-level regression trends, and shaded areas represent 95% confidence intervals. Δ slope represents the difference in the longitudinal rate of change between EG and CG, and $P_{interaction}$ denotes the significance of the time-by-treatment interaction in linear mixed-effects models. Outcome-specific record and participant counts are provided in Table S4. These selected parameters do not constitute a comprehensive assessment of elemental safety.

#### Supplementary Figure 3. Targeted tryptophan metabolite profiles and exploratory associations with BUN in CKD mice

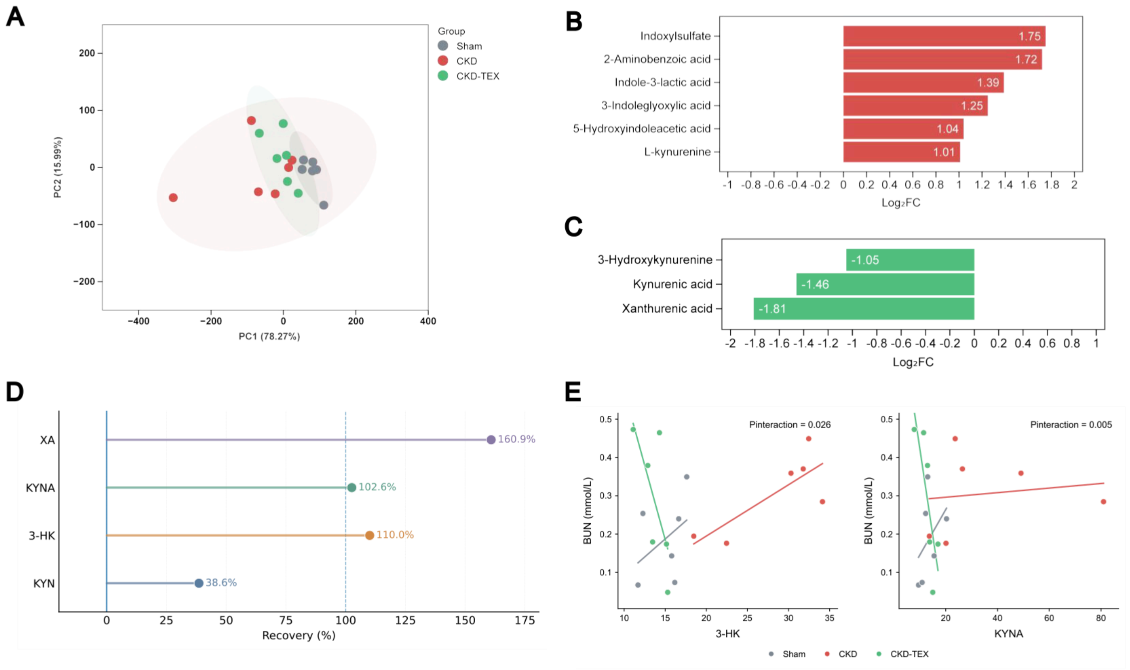


**(A)** Principal component analysis (PCA) of targeted tryptophan-related metabolites across the Sham, CKD, and CKD-TEX groups. Each point represents an individual mouse.

**(B)** Tryptophan-related metabolites increased in CKD relative to Sham. Metabolites shown met the predefined criteria of $|\log_2 FC| \geq 1$ and Tukey-adjusted $P < 0.05$.

**(C)** Tryptophan-related metabolites altered in CKD-TEX relative to CKD using the same criteria. Negative $\log_2$ fold changes indicate lower abundance in CKD-TEX than in CKD.

**(D)** Descriptive recovery indices for CKD-associated alterations in selected kynurenine pathway metabolites following TEX supplementation. Recovery = (CKD − CKD-TEX)/(CKD − Sham) × 100%, with positive values indicating attenuation towards the Sham level. These indices summarise direction and magnitude and are not independent confirmatory tests.

**(E)** Group-dependent associations between selected kynurenine pathway metabolites and blood urea nitrogen (BUN). Regression models included metabolite abundance, experimental group, and the metabolite × group interaction term. P values indicate the significance of the interaction term.

n = 6 mice per group. Differential metabolite analyses were performed by one-way ANOVA followed by Tukey's multiple-comparisons test. Interaction analyses used rank-transformed regression models.

**Supplementary Figure 4. Selected renal stress-response and inflammatory markers after TEX supplementation in CKD mice**

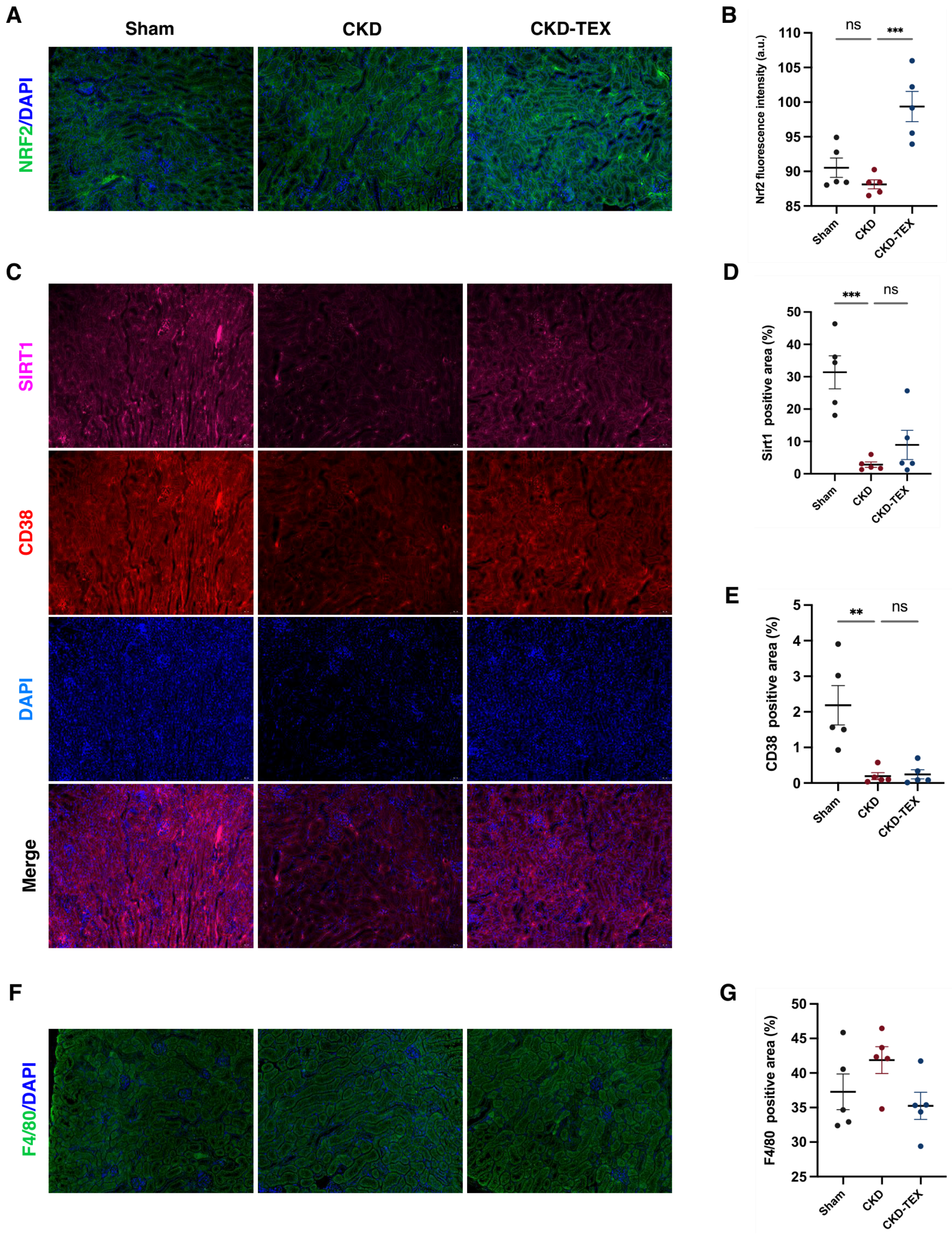


**(A)** Representative renal sections showing NRF2 immunofluorescence in Sham, CKD, and CKD-TEX mice. **(B)** Quantification of NRF2 fluorescence intensity. **(C)** Representative dual immunofluorescence staining for SIRT1 (magenta), CD38 (red), and DAPI (blue). **(D and E)** Quantification of SIRT1-positive area and CD38-positive area, respectively. **(F)** Representative F4/80/DAPI immunofluorescence images. **(G)** Quantification of F4/80-positive area. Data are presented as mean ± SEM; individual points represent biological replicates (n = 5 per group). Statistical analysis used ordinary one-way ANOVA followed by Tukey's multiple-comparisons test. ns, not significant; *P < 0.05, **P < 0.01, ***P < 0.001.

### Table S1. Elemental composition, ICP-OES operating conditions and replicate measurements

#### Table S1A. Elemental composition of TEX determined by ICP-OES (n = 3)

| Element | Content (mg/L), mean ± SD | CV (%) |
|---|---|---|
| Fe | 2304 ± 42 | 1.8 |
| Se | 23.1 ± 1.0 | 4.4 |
| Mg | 1974 ± 25 | 1.2 |
| Si | 20.2 ± 1.2 | 5.7 |
| Al | 1463 ± 10 | 0.7 |
| V | 5.6 ± 0.3 | 5.7 |
| Zn | 723 ± 16 | 2.2 |
| Cr | 5.4 ± 0.3 | 5.1 |
| K | 558 ± 5 | 0.9 |
| Ni | 4.2 ± 0.2 | 5.9 |
| Li | 257 ± 4 | 1.4 |
| Cu | 3.0 ± 0.2 | 5.6 |
| Mn | 133 ± 8 | 6 |
| Mo | 1.4 ± 0.0 | 2.6 |
| Ti | 96.4 ± 4.4 | 4.5 |
| Co | 0.79 ± 0.07 | 9 |
| Ca | 67.0 ± 2.6 | 3.8 |
| Sc | 0.35 ± 0.05 | 14.9 |
| Na | 46.1 ± 2.7 | 5.8 |
| Sr | 0.21 ± 0.02 | 10.7 |

Three independent digestions of the same composite solution were analysed; these were not three independent formulations. Ag was detected at 8.6 mg/L in only one replicate and was excluded as an analytical artefact. All other screened elements (As, Au, B, Ba, Be, Bi, Cd, Ce, Dy, Er, Eu, Ga, Gd, Ge, Hf, Hg, Ho, In, Ir, La, Lu, Nb, Nd, P, Pb, Pd, Pr, Pt, Rb, Re, Ru, Rh, Sb, Sn, Ta, Tb, Te, Th, Tl, Tm, U, W, Y, Yb, Zr) were below the limit of detection in all replicates.

#### Table S1B. ICP-OES operating conditions and sample preparation

| Parameter | Value |
|---|---|
| Instrument | Agilent 5800 ICP-OES |
| RF power | 1200 W |
| Plasma gas flow | 12 L/min |
| Auxiliary gas flow | 1 L/min |
| Nebulizer gas flow | 0.7 L/min |
| Sample uptake delay | 15 s |
| Instrument stabilization time | 15 s |
| Replicate read time | 5 s |
| Replicates per measurement | 3 |
| Sample digestion | 2 mL of solution digested and diluted to a final volume of 10 mL (V0/V = 5) |
| Dilution factor | f = 20 for Fe, Mg, Al, Zn, K, Li; f = 1 for all other elements |
| Quantification | Cx = C0 × f × V0 / V, where C0 is the measured concentration of the test solution and Cx the concentration in the original sample |
| Analysis date | 12 May 2026 |
| Test item | Full-element screening |

#### Table S1C. Replicate digestion and measurement values

| Element | Replicate 1 (mg/L) | Replicate 2 (mg/L) | Replicate 3 (mg/L) | Mean | SD | CV (%) |
|---|---|---|---|---|---|---|
| Al | 1472.1 | 1451.8 | 1465.1 | 1463 | 10.312 | 0.705 |
| Ca | 64.905 | 66.3 | 69.875 | 67.027 | 2.563 | 3.825 |
| Co | 0.705 | 0.82 | 0.835 | 0.787 | 0.071 | 9.041 |
| Cr | 5.135 | 5.365 | 5.68 | 5.393 | 0.274 | 5.073 |
| Cu | 2.865 | 3.085 | 3.2 | 3.05 | 0.17 | 5.581 |
| Fe | 2351 | 2289 | 2271.3 | 2303.767 | 41.852 | 1.817 |
| K | 561.1 | 552.1 | 560.2 | 557.8 | 4.957 | 0.889 |
| Li | 254.6 | 255.3 | 261.1 | 257 | 3.568 | 1.388 |
| Mg | 2001.8 | 1960.6 | 1958.2 | 1973.533 | 24.509 | 1.242 |
| Mn | 125.015 | 133.19 | 140.975 | 133.06 | 7.981 | 5.998 |
| Mo | 1.365 | 1.335 | 1.405 | 1.368 | 0.035 | 2.567 |
| Na | 43.66 | 45.8 | 48.935 | 46.132 | 2.653 | 5.751 |
| Ni | 3.93 | 4.175 | 4.425 | 4.177 | 0.248 | 5.926 |
| Sc | 0.315 | 0.325 | 0.41 | 0.35 | 0.052 | 14.915 |
| Se | 22.21 | 22.965 | 24.205 | 23.127 | 1.007 | 4.355 |
| Si | 18.875 | 20.54 | 21.085 | 20.167 | 1.151 | 5.709 |
| Sr | 0.19 | 0.21 | 0.235 | 0.212 | 0.023 | 10.652 |
| Ti | 93.2 | 94.54 | 101.365 | 96.368 | 4.379 | 4.544 |
| V | 5.325 | 5.64 | 5.965 | 5.643 | 0.32 | 5.671 |
| Zn | 740.8 | 718.6 | 709.6 | 723 | 16.059 | 2.221 |

Mean, SD and CV values are reproduced from the source workbook. CV, coefficient of variation; SD, standard deviation.

#### Table S2. Primer sequences used for quantitative real-time PCR analysis

Primer sequences for the target genes (IL-1β, IL-6, and IL-10) and the reference gene β-actin are shown in the 5′–3′ direction.

| Primer | Direction | Sequence (5′–3′) | Size (bp) |
|---|---|---|---|
| IL-1β | F | TTTCCTCCTTGCCTCTGATG | 134 |
| IL-1β | R | CCACACGTTGACAGCTAGGT | 134 |
| IL-6 | F | GAAATGATGGATGCTACCAAACTG | 139 |
| IL-6 | R | ACTCTGGCTTTGTCTTTCTTGTTATC | 139 |
| IL-10 | F | GTTGCCAAGCCTTATCGGAAATG | 199 |
| IL-10 | R | TCACTCTTCACCTGCTCCACTGC | 199 |
| β-actin | F | CGGTGCTGAGTATGTCGTGGAGTC | 100 |
| β-actin | R | GGCGGAGATGATGACCCTTTTG | 100 |

#### Table S3A. Participant characteristics and serum creatinine

De-identified individual participant data are shown in source-workbook row order. Patient_ID, Group and Time_Month are repeated across Table S3A–F for row-level cross-reference. Blank cells indicate missing values; no imputation was applied. Ca×P is the source-workbook product of Ca and P. Field definitions, units and availability are given in Table S4.

| Patient_ID | Group | Sex | Age | BMI | Time_Month | CREA |
|---|---|---|---|---|---|---|
| P001 | EG | M | 60–64 | 21.71 | 0 | 1102 |
| P001 | EG | M | 60–64 | 21.71 | 1.7 | 1045 |
| P001 | EG | M | 60–64 | 21.71 | 2.9 | 1051 |
| P002 | EG | M | 40–44 | 27.17 | 0 | 855 |
| P002 | EG | M | 40–44 | 27.17 | 3.6 | 867 |
| P002 | EG | M | 40–44 | 27.17 | 6.7 | 891 |
| P003 | EG | M | 40–44 | 21.13 | 0 | 628 |
| P003 | EG | M | 40–44 | 21.13 | 5.2 | 769 |
| P003 | EG | M | 40–44 | 21.13 | 8.8 | 692 |
| P004 | EG | F | 45–49 | 21.78 | 0 | 1156 |
| P004 | EG | F | 45–49 | 21.78 | 2.2 | 1056 |
| P004 | EG | F | 45–49 | 21.78 | 8.2 | 1196 |
| P005 | EG | M | 55–59 | 18.89 | 0 | 964 |
| P005 | EG | M | 55–59 | 18.89 | 3.8 | 932 |
| P005 | EG | M | 55–59 | 18.89 | 6.1 | 1012 |
| P006 | EG | F | 40–44 | 21.97 | 0 | 733 |
| P006 | EG | F | 40–44 | 21.97 | 3 | 746 |
| P006 | EG | F | 40–44 | 21.97 | 6.8 | 874 |
| P007 | EG | M | 40–44 | 21.88 | 0 | 1054 |
| P007 | EG | M | 40–44 | 21.88 | 2.6 | 337 |
| P007 | EG | M | 40–44 | 21.88 | 4.8 | 1024 |
| P008 | EG | F | 25–29 | 17.47 | 0 | 1524 |
| P008 | EG | F | 25–29 | 17.47 | 7.9 | 1405 |
| P008 | EG | F | 25–29 | 17.47 | 10.9 | 1118 |
| P009 | EG | M | 40–44 | 24.42 | 0 | 708 |
| P009 | EG | M | 40–44 | 24.42 | 3.8 | 677 |
| P009 | EG | M | 40–44 | 24.42 | 7.1 | 734 |
| P010 | EG | F | 30–34 | 18.67 | 0 | 986 |
| P010 | EG | F | 30–34 | 18.67 | 1.4 | 961 |
| P010 | EG | F | 30–34 | 18.67 | 6.8 | 1155 |
| P011 | EG | F | 25–29 | 17.96 | 0 | 945 |
| P011 | EG | F | 25–29 | 17.96 | 5.2 | 1002 |
| P011 | EG | F | 25–29 | 17.96 | 8 | 1069 |
| P012 | EG | M | 60–64 | 24.68 | 0 | 815 |
| P012 | EG | M | 60–64 | 24.68 | 6.5 | 847 |
| P012 | EG | M | 60–64 | 24.68 | 9.8 | 872 |
| P013 | EG | M | 45–49 | 21.45 | 0 | 1466 |
| P013 | EG | M | 45–49 | 21.45 | 3.4 | 1296 |
| P013 | EG | M | 45–49 | 21.45 | 5.8 | 1285 |
| P014 | EG | F | 35–39 | 21.09 | 0 | 805 |
| P014 | EG | F | 35–39 | 21.09 | 3.2 | 860 |
| P014 | EG | F | 35–39 | 21.09 | 5.2 | 1048 |
| P015 | EG | F | 50–54 | 20.21 | 0 | 974 |
| P015 | EG | F | 50–54 | 20.21 | 1.5 | 792 |
| P015 | EG | F | 50–54 | 20.21 | 3.9 | 996 |
| P016 | EG | M | 35–39 | 20.15 | 0 | 1164 |
| P016 | EG | M | 35–39 | 20.15 | 3.1 | 1213 |
| P016 | EG | M | 35–39 | 20.15 | 5.4 | 1451 |
| P017 | EG | M | 55–59 | 21.91 | 0 | 937 |
| P017 | EG | M | 55–59 | 21.91 | 2.8 | 894 |
| P017 | EG | M | 55–59 | 21.91 | 6.1 | 978 |
| P018 | EG | F | 60–64 | 27.44 | 0 | 440 |

| | | | | | | |
|---|---|---|---|---|---|---|
| P018 | EG | F | 60–64 | 27.44 | 1.6 | 404 |
| P018 | EG | F | 60–64 | 27.44 | 6.1 | 525 |
| P019 | EG | F | 55–59 | 24.69 | 0 | 621 |
| P019 | EG | F | 55–59 | 24.69 | 4.7 | 685 |
| P019 | EG | F | 55–59 | 24.69 | 9.6 | 738 |
| P020 | EG | M | 35–39 | 21.88 | 0 | 1263 |
| P020 | EG | M | 35–39 | 21.88 | 2.9 | 1139 |
| P020 | EG | M | 35–39 | 21.88 | 5.7 | 912 |
| P021 | EG | F | 35–39 | 21.09 | 0 | 859 |
| P021 | EG | F | 35–39 | 21.09 | 2.7 | 381 |
| P021 | EG | F | 35–39 | 21.09 | 6.6 | 1175 |
| P022 | EG | M | 45–49 | 25.39 | 0 | 1214 |
| P022 | EG | M | 45–49 | 25.39 | 2 | 1304 |
| P022 | EG | M | 45–49 | 25.39 | 6.7 | 1310 |
| P023 | EG | F | 35–39 | 21.34 | 0 | 883 |
| P023 | EG | F | 35–39 | 21.34 | 2.5 | 891 |
| P023 | EG | F | 35–39 | 21.34 | 6.4 | 931 |
| P024 | EG | M | 40–44 | 23.66 | 0 | 1033 |
| P024 | EG | M | 40–44 | 23.66 | 4.1 | 1313 |
| P024 | EG | M | 40–44 | 23.66 | 6.1 | 1276 |
| P025 | EG | F | 45–49 | 19.13 | 0 | 894 |
| P025 | EG | F | 45–49 | 19.13 | 2.8 | 828 |
| P025 | EG | F | 45–49 | 19.13 | 6.1 | 919 |
| P026 | EG | F | 35–39 | 19.04 | 0 | 773 |
| P026 | EG | F | 35–39 | 19.04 | 4.6 | 734 |
| P026 | EG | F | 35–39 | 19.04 | 8.1 | 730 |
| P027 | EG | F | 30–34 | 18.32 | 0 | 1006 |
| P027 | EG | F | 30–34 | 18.32 | 3.2 | 913 |
| P027 | EG | F | 30–34 | 18.32 | 6.3 | 886 |
| P028 | EG | M | 30–34 | 17.79 | 0 | 1141 |
| P028 | EG | M | 30–34 | 17.79 | 2 | 1339 |
| P028 | EG | M | 30–34 | 17.79 | 5.3 | 1430 |
| P029 | EG | F | 40–44 | 31.25 | 0 | 927 |
| P029 | EG | F | 40–44 | 31.25 | 1.5 | 1134 |
| P029 | EG | F | 40–44 | 31.25 | 6.2 | 1272 |
| P030 | EG | F | 30–34 | 25.96 | 0 | 615 |
| P030 | EG | F | 30–34 | 25.96 | 3 | 673 |
| P030 | EG | F | 30–34 | 25.96 | 8.1 | 632 |
| P031 | EG | M | 35–39 | 23.45 | 0 | 1036 |
| P032 | EG | M | 50–54 | 20.78 | 0 | 845 |
| P032 | EG | M | 50–54 | 20.78 | 2.6 | 888 |
| P032 | EG | M | 50–54 | 20.78 | 5.6 | 933 |
| P033 | EG | M | 65–69 | 24.34 | 0 | 967 |
| P033 | EG | M | 65–69 | 24.34 | 2.8 | 1046 |
| P033 | EG | M | 65–69 | 24.34 | 6.6 | 902 |
| P034 | EG | M | 40–44 | 21.22 | 0 | 1121 |
| P034 | EG | M | 40–44 | 21.22 | 1.2 | 1198 |
| P035 | EG | F | 60–64 | 19.19 | 0 | 751 |
| P035 | EG | F | 60–64 | 19.19 | 4 | 763 |
| P035 | EG | F | 60–64 | 19.19 | 7.3 | 748 |
| P036 | EG | F | 45–49 | 26.06 | 0 | 820 |
| P036 | EG | F | 45–49 | 26.06 | 5.1 | 716 |
| P036 | EG | F | 45–49 | 26.06 | 9 | 678 |
| P037 | EG | M | 30–34 | 34.66 | 0 | 1382 |
| P037 | EG | M | 30–34 | 34.66 | 2.2 | 1532 |
| P037 | EG | M | 30–34 | 34.66 | 6.5 | 1335 |
| P038 | EG | M | 35–39 | 19.9 | 0 | 800 |
| P038 | EG | M | 35–39 | 19.9 | 5.4 | 857 |

| | | | | | | |
|---|---|---|---|---|---|---|
| P038 | EG | M | 35–39 | 19.9 | 7 | 806 |
| P039 | EG | M | 35–39 | 26.57 | 0 | 1062 |
| P039 | EG | M | 35–39 | 26.57 | 1.5 | 1443 |
| P039 | EG | M | 35–39 | 26.57 | 6.2 | 828 |
| P040 | EG | M | 45–49 | 27.47 | 0 | 1184 |
| P040 | EG | M | 45–49 | 27.47 | 1.8 | 1004 |
| P040 | EG | M | 45–49 | 27.47 | 3.9 | 1019 |
| P041 | CG | M | 30–34 | 22.68 | 0 | 1168 |
| P041 | CG | M | 30–34 | 22.68 | 4.8 | 1221 |
| P041 | CG | M | 30–34 | 22.68 | 9.3 | 925 |
| P042 | CG | M | 25–29 | 21.3 | 0 | 301 |
| P042 | CG | M | 25–29 | 21.3 | 4.2 | 301 |
| P042 | CG | M | 25–29 | 21.3 | 5.9 | 497 |
| P043 | CG | M | 45–49 | 27.68 | 0 | 1268 |
| P043 | CG | M | 45–49 | 27.68 | 3.6 | 1116 |
| P043 | CG | M | 45–49 | 27.68 | 5.1 | 1005 |
| P044 | CG | F | 45–49 | 24.23 | 0 | 760 |
| P044 | CG | F | 45–49 | 24.23 | 5.1 | 745 |
| P044 | CG | F | 45–49 | 24.23 | 8.8 | 707 |
| P045 | CG | M | 40–44 | 22.11 | 0 | 1133 |
| P045 | CG | M | 40–44 | 22.11 | 2.1 | 982 |
| P045 | CG | M | 40–44 | 22.11 | 6.1 | 928 |
| P046 | CG | M | 35–39 | 29.13 | 0 | 474 |
| P046 | CG | M | 35–39 | 29.13 | 1.8 | 524 |
| P046 | CG | M | 35–39 | 29.13 | 7 | 745 |
| P047 | CG | M | 35–39 | 19.78 | 0 | 1162 |
| P047 | CG | M | 35–39 | 19.78 | 5.3 | 1192 |
| P047 | CG | M | 35–39 | 19.78 | 10.1 | 1283 |
| P048 | CG | M | 45–49 | 33.81 | 0 | 1054 |
| P048 | CG | M | 45–49 | 33.81 | 4.5 | 1038 |
| P048 | CG | M | 45–49 | 33.81 | 7.2 | 992 |
| P049 | CG | M | 45–49 | 24.26 | 0 | 1016 |
| P049 | CG | M | 45–49 | 24.26 | 5.5 | 907 |
| P049 | CG | M | 45–49 | 24.26 | 7.8 | 993 |
| P050 | CG | M | 30–34 | 21.52 | 0 | 667 |
| P050 | CG | M | 30–34 | 21.52 | 4.6 | 1084 |
| P050 | CG | M | 30–34 | 21.52 | 6.4 | 1285 |
| P051 | CG | F | 40–44 | 16.21 | 0 | 863 |
| P051 | CG | F | 40–44 | 16.21 | 3.3 | 949 |
| P051 | CG | F | 40–44 | 16.21 | 8.7 | 1143 |
| P052 | CG | M | 40–44 | 28.83 | 0 | 572 |
| P052 | CG | M | 40–44 | 28.83 | 3.4 | 624 |
| P052 | CG | M | 40–44 | 28.83 | 6.2 | 683 |
| P053 | CG | F | 35–39 | 25.47 | 0 | 1089 |
| P053 | CG | F | 35–39 | 25.47 | 2.2 | 1132 |
| P053 | CG | F | 35–39 | 25.47 | 7.8 | 1208 |
| P054 | CG | M | 55–59 | 22.02 | 0 | 1140 |
| P054 | CG | M | 55–59 | 22.02 | 1.5 | 1254 |
| P054 | CG | M | 55–59 | 22.02 | 3.7 | 1061 |
| P055 | CG | M | 50–54 | 23.18 | 0 | 1171 |
| P055 | CG | M | 50–54 | 23.18 | 5 | 1110 |
| P055 | CG | M | 50–54 | 23.18 | 9.7 | 1139 |
| P056 | CG | M | 55–59 | 21.05 | 0 | 794 |
| P056 | CG | M | 55–59 | 21.05 | 2.5 | 876 |
| P057 | CG | M | 35–39 | 28.22 | 0 | 1126 |
| P057 | CG | M | 35–39 | 28.22 | 3.9 | 1020 |
| P057 | CG | M | 35–39 | 28.22 | 9 | 1250 |
| P058 | CG | M | 45–49 | 24.61 | 0 | 632 |

| | | | | | | |
|---|---|---|---|---|---|---|
| P058 | CG | M | 45–49 | 24.61 | 1.5 | 855 |
| P058 | CG | M | 45–49 | 24.61 | 5.1 | 936 |
| P059 | CG | M | 45–49 | 19.88 | 0 | 1047 |
| P059 | CG | M | 45–49 | 19.88 | 1.5 | 1081 |
| P059 | CG | M | 45–49 | 19.88 | 3.3 | 1055 |
| P060 | CG | M | 45–49 | 22.72 | 0 | 1373 |
| P060 | CG | M | 45–49 | 22.72 | 1.7 | 1204 |
| P060 | CG | M | 45–49 | 22.72 | 5.5 | 1322 |
| P061 | CG | M | 65–69 | 21.86 | 0 | 634 |
| P061 | CG | M | 65–69 | 21.86 | 4.4 | 807 |
| P061 | CG | M | 65–69 | 21.86 | 7.7 | 1046 |
| P062 | CG | F | 40–44 | 21.55 | 0 | 1108 |
| P062 | CG | F | 40–44 | 21.55 | 5.2 | 1092 |
| P062 | CG | F | 40–44 | 21.55 | 9.6 | 1116 |
| P063 | CG | F | 70–74 | 22.27 | 0 | 193 |
| P063 | CG | F | 70–74 | 22.27 | 1.9 | 240 |
| P063 | CG | F | 70–74 | 22.27 | 3.5 | 238 |
| P064 | CG | M | 70–74 | 24.38 | 0 | 510 |
| P064 | CG | M | 70–74 | 24.38 | 4.7 | 869 |
| P064 | CG | M | 70–74 | 24.38 | 9.7 | 776 |
| P065 | CG | F | 35–39 | 16.85 | 0 | 1131 |
| P065 | CG | F | 35–39 | 16.85 | 2.7 | 974 |
| P065 | CG | F | 35–39 | 16.85 | 7 | 975 |
| P066 | CG | M | 25–29 | 30.37 | 0 | 1598 |
| P066 | CG | M | 25–29 | 30.37 | 3.5 | 1458 |
| P066 | CG | M | 25–29 | 30.37 | 5.9 | 1639 |
| P067 | CG | F | 55–59 | 21.27 | 0 | 398 |
| P067 | CG | F | 55–59 | 21.27 | 2.4 | 525 |
| P067 | CG | F | 55–59 | 21.27 | 4.5 | 520 |
| P068 | CG | M | 65–69 | 20.76 | 0 | 633 |
| P068 | CG | M | 65–69 | 20.76 | 2.8 | 614 |
| P068 | CG | M | 65–69 | 20.76 | 4.5 | 679 |
| P069 | CG | F | 75–79 | 17.44 | 0 | 775 |
| P069 | CG | F | 75–79 | 17.44 | 5.8 | 634 |
| P069 | CG | F | 75–79 | 17.44 | 9.2 | 545 |
| P070 | CG | M | 40–44 | 18.62 | 0 | 765 |
| P070 | CG | M | 40–44 | 18.62 | 4 | 674 |
| P070 | CG | M | 40–44 | 18.62 | 8.2 | 693 |
| P071 | CG | F | 70–74 | 19.04 | 0 | 857 |
| P071 | CG | F | 70–74 | 19.04 | 4.1 | 1088 |
| P071 | CG | F | 70–74 | 19.04 | 7.3 | 886 |
| P072 | CG | F | 75–79 | 18.08 | 0 | 650 |
| P072 | CG | F | 75–79 | 18.08 | 1.6 | 807 |
| P072 | CG | F | 75–79 | 18.08 | 5.1 | 718 |
| P073 | CG | F | 50–54 | 22.96 | 0 | 458 |
| P073 | CG | F | 50–54 | 22.96 | 3 | 421 |
| P073 | CG | F | 50–54 | 22.96 | 4.5 | 597 |
| P074 | CG | F | 55–59 | 24.69 | 0 | 603 |
| P074 | CG | F | 55–59 | 24.69 | 5.9 | 462 |
| P074 | CG | F | 55–59 | 24.69 | 8.5 | 738 |
| P075 | CG | M | 55–59 | 26.12 | 0 | 486 |
| P075 | CG | M | 55–59 | 26.12 | 3.8 | 798 |
| P075 | CG | M | 55–59 | 26.12 | 8.6 | 885 |
| P076 | CG | M | 45–49 | 30.83 | 0 | 805 |
| P076 | CG | M | 45–49 | 30.83 | 2.6 | 891 |
| P076 | CG | M | 45–49 | 30.83 | 7.2 | 1031 |
| P077 | CG | F | 45–49 | 21.63 | 0 | 832 |
| P077 | CG | F | 45–49 | 21.63 | 3.1 | 812 |

| | | | | | | |
|---|---|---|---|---|---|---|
| P077 | CG | F | 45–49 | 21.63 | 8.1 | 799 |
| P078 | CG | F | 40–44 | 22.14 | 0 | 1225 |
| P078 | CG | F | 40–44 | 22.14 | 2.3 | 902 |
| P078 | CG | F | 40–44 | 22.14 | 4.8 | 948 |
| P079 | CG | M | 55–59 | 24.31 | 0 | 685 |
| P079 | CG | M | 55–59 | 24.31 | 5.6 | 738 |
| P079 | CG | M | 55–59 | 24.31 | 7.1 | 823 |
| P080 | CG | M | 45–49 | 21.22 | 0 | 780 |
| P080 | CG | M | 45–49 | 21.22 | 1.6 | 835 |
| P080 | CG | M | 45–49 | 21.22 | 5.5 | 774 |

#### Table S3B. Urea, estimated glomerular filtration rate and cystatin C

De-identified individual participant data are shown in source-workbook row order. Patient_ID, Group and Time_Month are repeated across Table S3A–F for row-level cross-reference. Blank cells indicate missing values; no imputation was applied. Ca×P is the source-workbook product of Ca and P. Field definitions, units and availability are given in Table S4.

| Patient_ID | Group | Time_Month | BUN | eGFR | Cys_c |
|---|---|---|---|---|---|
| P001 | EG | 0 | 19.3 | 4.17295503318 | 7.23 |
| P001 | EG | 1.7 | 27.9 | 4.44756260217 | 7.21 |
| P001 | EG | 2.9 | 24.3 | 4.4171114683 | 7.13 |
| P002 | EG | 0 | 28.1 | 6.32878609406 | 5.37 |
| P002 | EG | 3.6 | 28.4 | 6.22381733567 | 5.39 |
| P002 | EG | 6.7 | 21.1 | 6.02318926475 | |
| P003 | EG | 0 | 24.7 | 9.05161695497 | 6.66 |
| P003 | EG | 5.2 | 26.7 | 7.09849256818 | 6.55 |
| P003 | EG | 8.8 | 20 | 8.05657448107 | 7.14 |
| P004 | EG | 0 | 26.4 | 3.19766133236 | 5.75 |
| P004 | EG | 2.2 | 24.9 | 3.56438957123 | 5.86 |
| P004 | EG | 8.2 | 26.9 | 3.06976019646 | 6.4 |
| P005 | EG | 0 | 28.4 | 4.96102245515 | 7.35 |
| P005 | EG | 3.8 | 25.1 | 5.1661205804 | 6.67 |
| P005 | EG | 6.1 | 32.7 | 4.68001255847 | 7.13 |
| P006 | EG | 0 | 15.3 | 5.7698450428 | 4.33 |
| P006 | EG | 3 | 14.5 | 5.6494000095 | 4.79 |
| P006 | EG | 6.8 | 20.2 | 4.67170273217 | |
| P007 | EG | 0 | 32.3 | 4.98509038631 | 6.76 |
| P007 | EG | 2.6 | 7 | 19.5851043156 | 4.99 |
| P007 | EG | 4.8 | 25.5 | 5.1608569594 | 6.82 |
| P008 | EG | 0 | 39.3 | 2.6152972983 | 6.52 |
| P008 | EG | 7.9 | 36.2 | 2.88331065241 | 5.64 |
| P008 | EG | 10.9 | 26.1 | 3.79291282643 | |
| P009 | EG | 0 | 16.9 | 7.88749029906 | 5.66 |
| P009 | EG | 3.8 | 20.2 | 8.32285531528 | 5.78 |
| P009 | EG | 7.1 | 19.9 | 7.55341724466 | 7.1 |
| P010 | EG | 0 | 20.4 | 4.24858862583 | 6.48 |
| P010 | EG | 1.4 | 22.3 | 4.38156152979 | 7.12 |
| P010 | EG | 6.8 | 22.3 | 3.51397444613 | |
| P011 | EG | 0 | 17.4 | 4.6696664772 | 6.11 |
| P011 | EG | 5.2 | 24.8 | 4.35274040971 | 5.82 |
| P011 | EG | 8 | 21.2 | 4.02745583736 | 5.52 |
| P012 | EG | 0 | 25.2 | 5.88260198343 | 6.25 |
| P012 | EG | 6.5 | 27.5 | 5.61692342382 | 6.64 |
| P012 | EG | 9.8 | 28.6 | 5.42423895838 | |
| P013 | EG | 0 | 31.4 | 3.25248998367 | 7.16 |
| P013 | EG | 3.4 | 27.5 | 3.77094962048 | 7.33 |
| P013 | EG | 5.8 | 28.7 | 3.80971930792 | 6.67 |
| P014 | EG | 0 | 31.8 | 5.28613065529 | 6.97 |
| P014 | EG | 3.2 | 20.4 | 4.8830908922 | 6.32 |
| P014 | EG | 5.2 | 42.8 | 3.85176170038 | 7.36 |
| P015 | EG | 0 | 32.7 | 3.80720340285 | 7.11 |
| P015 | EG | 1.5 | 25.9 | 4.87985154752 | 6.55 |
| P015 | EG | 3.9 | 36 | 3.7065138059 | 6.26 |
| P016 | EG | 0 | 38.1 | 4.50859491106 | 5.43 |
| P016 | EG | 3.1 | 30.4 | 4.29093397286 | 6.12 |
| P016 | EG | 5.4 | 39.2 | 3.46085893651 | 6.34 |
| P017 | EG | 0 | 30.8 | 5.22972854837 | 8.03 |
| P017 | EG | 2.8 | 26.9 | 5.53301227494 | 7.18 |
| P017 | EG | 6.1 | 32.8 | 4.9677534142 | |
| P018 | EG | 0 | 14.3 | 9.34167057222 | 4.6 |

| | | | | | |
|---|---|---|---|---|---|
| P018 | EG | 1.6 | 16.8 | 10.3492796381 | 4.62 |
| P018 | EG | 6.1 | 16.5 | 7.55747285262 | |
| P019 | EG | 0 | 16 | 6.41303906209 | 5.37 |
| P019 | EG | 4.7 | 13.4 | 5.70092227072 | 5.93 |
| P019 | EG | 9.6 | 15.5 | 5.21322142292 | |
| P020 | EG | 0 | 39.3 | 4.13907015379 | 7.82 |
| P020 | EG | 2.9 | 32.6 | 4.68552586018 | |
| P020 | EG | 5.7 | 28.5 | 6.11776797973 | |
| P021 | EG | 0 | 21.1 | 4.85959577126 | 6.02 |
| P021 | EG | 2.7 | 7.2 | 12.8908623582 | 3.28 |
| P021 | EG | 6.6 | 33.1 | 3.33692543401 | 6.33 |
| P022 | EG | 0 | 25.1 | 4.05334248225 | 5.54 |
| P022 | EG | 2 | 19.6 | 3.71999721623 | 5.79 |
| P022 | EG | 6.7 | 22.8 | 3.69956080595 | 5.92 |
| P023 | EG | 0 | 24.1 | 4.73086009995 | 6.58 |
| P023 | EG | 2.5 | 26.7 | 4.67993374626 | 6.1 |
| P023 | EG | 6.4 | 29.8 | 4.43969701068 | 6.26 |
| P024 | EG | 0 | 29.8 | 5.01254588869 | 6.43 |
| P024 | EG | 4.1 | 31.3 | 3.75890263053 | 6.05 |
| P024 | EG | 6.1 | 37.7 | 3.89007467343 | |
| P025 | EG | 0 | 27.6 | 4.32589362715 | 5.89 |
| P025 | EG | 2.8 | 26.4 | 4.7429053121 | 6.13 |
| P025 | EG | 6.1 | 27.2 | 4.18506535965 | 6.14 |
| P026 | EG | 0 | 20.7 | 5.44721521448 | 4.65 |
| P026 | EG | 4.6 | 25.6 | 5.79635071367 | 4.28 |
| P026 | EG | 8.1 | 23.4 | 5.83448455805 | 4.66 |
| P027 | EG | 0 | 21.4 | 4.19934367031 | 6.04 |
| P027 | EG | 3.2 | 22.5 | 4.71774064544 | 6.06 |
| P027 | EG | 6.3 | 19.8 | 4.89078452006 | 5.93 |
| P028 | EG | 0 | 30.9 | 4.79344849308 | 6.55 |
| P028 | EG | 2 | 30.1 | 3.9559805348 | 6.25 |
| P028 | EG | 5.3 | 30.5 | 3.65584347777 | 7.31 |
| P029 | EG | 0 | 15.7 | 4.27258012135 | 6.84 |
| P029 | EG | 1.5 | 29.5 | 3.35467329173 | 7.94 |
| P029 | EG | 6.2 | 26.8 | 2.92281523864 | |
| P030 | EG | 0 | 19.1 | 7.39341060159 | 7.29 |
| P030 | EG | 3 | 22.2 | 6.63554941588 | 8.25 |
| P030 | EG | 8.1 | 17.8 | 7.15540913213 | |
| P031 | EG | 0 | 33.6 | 5.15290432373 | 6.47 |
| P032 | EG | 0 | 22.6 | 5.99432962226 | 5.45 |
| P032 | EG | 2.6 | 23.2 | 5.64771941696 | 6.11 |
| P032 | EG | 5.6 | 34.2 | 5.32243907606 | 6.05 |
| P033 | EG | 0 | 23.1 | 4.70266241741 | 6.06 |
| P033 | EG | 2.8 | 26 | 4.27974163071 | 6.05 |
| P033 | EG | 6.6 | 22.1 | 5.11219872527 | |
| P034 | EG | 0 | 50.8 | 4.60101900748 | 8.04 |
| P034 | EG | 1.2 | 40.2 | 4.24847024421 | 7.08 |
| P035 | EG | 0 | 25.2 | 4.8876356956 | 5.84 |
| P035 | EG | 4 | 24.9 | 4.79553766939 | 5.76 |
| P035 | EG | 7.3 | 20.6 | 4.9111685004 | |
| P036 | EG | 0 | 24.7 | 4.8888561471 | 5.61 |
| P036 | EG | 5.1 | 23.6 | 5.75291893765 | 6.17 |
| P036 | EG | 9 | 30.6 | 6.14197788969 | 4.33 |
| P037 | EG | 0 | 29.5 | 3.76165681762 | 6.49 |
| P037 | EG | 2.2 | 42.8 | 3.32413233497 | 7.36 |
| P037 | EG | 6.5 | 36 | 3.92113057231 | |
| P038 | EG | 0 | 21.9 | 7.11504770154 | 5.82 |
| P038 | EG | 5.4 | 20.1 | 6.55101832328 | 5.67 |

| | | | | | |
|---|---|---|---|---|---|
| P038 | EG | 7 | 26.3 | 7.0515363712 | 5.62 |
| P039 | EG | 0 | 18.2 | 5.00189263241 | 6.04 |
| P039 | EG | 1.5 | 38.2 | 3.46229595193 | 5.26 |
| P039 | EG | 6.2 | 21.3 | 6.74291028121 | |
| P040 | EG | 0 | 33.6 | 4.17689618988 | 6.86 |
| P040 | EG | 1.8 | 21.9 | 5.09090823923 | 8.01 |
| P040 | EG | 3.9 | 24.5 | 5.00111341716 | 6.79 |
| P041 | CG | 0 | 38.5 | 4.60317421591 | 7.66 |
| P041 | CG | 4.8 | 28.8 | 4.36445506508 | 6.9 |
| P041 | CG | 9.3 | 19.7 | 6.09002035152 | |
| P042 | CG | 0 | 15.2 | 24.0166848306 | 2.56 |
| P042 | CG | 4.2 | 20.3 | 24.0166848306 | 3.82 |
| P042 | CG | 5.9 | 14.8 | 13.1572518285 | 3.58 |
| P043 | CG | 0 | 32.6 | 3.82324045293 | 5.61 |
| P043 | CG | 3.6 | 20.3 | 4.45633348604 | |
| P043 | CG | 5.1 | 20.1 | 5.05330420091 | |
| P044 | CG | 0 | 23.9 | 5.35559306616 | 4.68 |
| P044 | CG | 5.1 | 23.1 | 5.48524908995 | 4.91 |
| P044 | CG | 8.8 | 20 | 5.84091094618 | 5.09 |
| P045 | CG | 0 | 29.6 | 4.48645024228 | 6.62 |
| P045 | CG | 2.1 | 26.4 | 5.32653717 | 6.23 |
| P045 | CG | 6.1 | 26 | 5.70060797277 | 6.12 |
| P046 | CG | 0 | 18 | 13.0872611214 | 3.59 |
| P046 | CG | 1.8 | 15.6 | 11.6033991859 | |
| P046 | CG | 7 | 16.2 | 7.60668181779 | 4.28 |
| P047 | CG | 0 | 22.6 | 4.46206018715 | 7.17 |
| P047 | CG | 5.3 | 22.6 | 4.32764144415 | 7.23 |
| P047 | CG | 10.1 | 26.7 | 3.96196641754 | 6.64 |
| P048 | CG | 0 | 21.2 | 4.832457009 | 7.81 |
| P048 | CG | 4.5 | 26 | 4.92198071721 | 7.95 |
| P048 | CG | 7.2 | 23.9 | 5.19711977139 | 7.26 |
| P049 | CG | 0 | 21.8 | 4.98772232351 | 6.76 |
| P049 | CG | 5.5 | 19.8 | 5.71539134085 | 6.38 |
| P049 | CG | 7.8 | 22.3 | 5.12667301793 | 6.63 |
| P050 | CG | 0 | 31.2 | 9.0164965851 | 4.03 |
| P050 | CG | 4.6 | 33.1 | 5.0344691575 | 4.42 |
| P050 | CG | 6.4 | 36.7 | 4.10492351974 | |
| P051 | CG | 0 | 19.7 | 4.71384129704 | 6.53 |
| P051 | CG | 3.3 | 25.1 | 4.20599214038 | 7.17 |
| P051 | CG | 8.7 | 24.6 | 3.36459221125 | |
| P052 | CG | 0 | 18 | 10.3158621001 | 4.53 |
| P052 | CG | 3.4 | 26.6 | 9.29307099317 | 4.79 |
| P052 | CG | 6.2 | 21.4 | 8.338269314 | 4.84 |
| P053 | CG | 0 | 31.4 | 3.61040956584 | 7.18 |
| P053 | CG | 2.2 | 24.6 | 3.44646771579 | 7.9 |
| P053 | CG | 7.8 | 31.9 | 3.18793608586 | 7.67 |
| P054 | CG | 0 | 38.8 | 4.08205469483 | 7.57 |
| P054 | CG | 1.5 | 30 | 3.64089032878 | 8.19 |
| P054 | CG | 3.7 | 28 | 4.4494484588 | 7.86 |
| P055 | CG | 0 | 24.8 | 4.12860581686 | 6.19 |
| P055 | CG | 5 | 25.4 | 4.40234546234 | 6.46 |
| P055 | CG | 9.7 | 27.1 | 4.26818490278 | 6.54 |
| P056 | CG | 0 | 20.7 | 6.30057286045 | 6.08 |
| P056 | CG | 2.5 | 24.9 | 5.59963492621 | |
| P057 | CG | 0 | 28 | 4.69179171754 | 7.48 |
| P057 | CG | 3.9 | 20.9 | 5.28280507967 | 8.49 |
| P057 | CG | 9 | 38.2 | 4.13897475279 | 7.59 |
| P058 | CG | 0 | 16.9 | 8.92722001384 | |

| | | | | | |
|---|---|---|---|---|---|
| P058 | CG | 1.5 | 19.8 | 6.211799 | |
| P058 | CG | 5.1 | 23.3 | 5.57244401174 | 6.66 |
| P059 | CG | 0 | 17.6 | 4.84105154212 | 7.59 |
| P059 | CG | 1.5 | 19.8 | 4.65891612451 | 7.9 |
| P059 | CG | 3.3 | 19.6 | 4.79703373901 | 3.14 |
| P060 | CG | 0 | 36 | 3.47512213169 | 7.43 |
| P060 | CG | 1.7 | 33.2 | 4.06839332099 | 7.66 |
| P060 | CG | 5.5 | 34.6 | 3.63661202783 | 8.67 |
| P061 | CG | 0 | 11.1 | 7.70808416535 | 6.73 |
| P061 | CG | 4.4 | 18.4 | 5.77039198438 | 6.56 |
| P061 | CG | 7.7 | 30.5 | 4.22683734776 | 7.12 |
| P062 | CG | 0 | 30.8 | 3.49253043386 | 6.82 |
| P062 | CG | 5.2 | 31.3 | 3.55402718694 | 6.41 |
| P062 | CG | 9.6 | 32.8 | 3.46250873901 | 6.67 |
| P063 | CG | 0 | 7.3 | 23.7459703272 | 2.19 |
| P063 | CG | 1.9 | 8.7 | 18.2812211137 | 2.7 |
| P063 | CG | 3.5 | 7.7 | 18.4657241308 | 2.27 |
| P064 | CG | 0 | 24.4 | 9.82350167383 | 4.11 |
| P064 | CG | 4.7 | 34.3 | 5.18235074589 | 4.49 |
| P064 | CG | 9.7 | 15 | 5.93630859402 | 4.74 |
| P065 | CG | 0 | 26.5 | 3.49330749895 | 7.15 |
| P065 | CG | 2.7 | 20.5 | 4.17946982274 | 6.16 |
| P065 | CG | 7 | 22.6 | 4.17432638761 | 5.76 |
| P066 | CG | 0 | 30.3 | 3.25988088381 | 6.05 |
| P066 | CG | 3.5 | 28.9 | 3.63902326741 | 5.96 |
| P066 | CG | 5.9 | 27.2 | 3.16227123002 | 5.91 |
| P067 | CG | 0 | 13.7 | 10.7352236385 | 4.48 |
| P067 | CG | 2.4 | 19.9 | 7.69980308376 | 5 |
| P067 | CG | 4.5 | 22.4 | 7.78873217421 | 4.74 |
| P068 | CG | 0 | 25.1 | 7.72269895466 | 5.87 |
| P068 | CG | 2.8 | 18.7 | 8.01035036651 | 5.73 |
| P068 | CG | 4.5 | 25.9 | 7.09920681754 | 6.24 |
| P069 | CG | 0 | 29.6 | 4.31410392665 | 6.44 |
| P069 | CG | 5.8 | 18.8 | 5.48966112708 | 6.84 |
| P069 | CG | 9.2 | 18 | 6.58228743786 | 6.33 |
| P070 | CG | 0 | 26.3 | 7.14305538566 | 5.45 |
| P070 | CG | 4 | 24.1 | 8.31545205587 | 5.01 |
| P070 | CG | 8.2 | 22.4 | 8.04262571662 | 4.79 |
| P071 | CG | 0 | 38 | 3.91994244146 | 6.11 |
| P071 | CG | 4.1 | 40.8 | 2.94375730354 | 6.02 |
| P071 | CG | 7.3 | 25.1 | 3.76648472198 | 6.54 |
| P072 | CG | 0 | 18.3 | 5.32790678612 | 6.58 |
| P072 | CG | 1.6 | 18.3 | 4.10964591008 | 7.08 |
| P072 | CG | 5.1 | 11 | 4.72828137398 | |
| P073 | CG | 0 | 13.9 | 9.29899982265 | 3.97 |
| P073 | CG | 3 | 14.2 | 10.2881265581 | 4.43 |
| P073 | CG | 4.5 | 23.1 | 6.76558906547 | 4.96 |
| P074 | CG | 0 | 11.2 | 6.60225088228 | 6.22 |
| P074 | CG | 5.9 | 7.7 | 9.08871378552 | 6.11 |
| P074 | CG | 8.5 | 15.5 | 5.1808994501 | |
| P075 | CG | 0 | 16.8 | 11.3553655092 | |
| P075 | CG | 3.8 | 27.2 | 6.26269369987 | |
| P075 | CG | 8.6 | 24.7 | 5.53137008036 | 4.62 |
| P076 | CG | 0 | 35.1 | 6.67761960747 | 5.94 |
| P076 | CG | 2.6 | 30 | 5.91185110311 | 6.76 |
| P076 | CG | 7.2 | 24.9 | 4.96210938685 | |
| P077 | CG | 0 | 42.3 | 4.71555556899 | 5.26 |
| P077 | CG | 3.1 | 32 | 4.85527265624 | |

| | | | | | |
|---|---|---|---|---|---|
| P077 | CG | 8.1 | 31.4 | 4.95022254089 | |
| P078 | CG | 0 | 34.3 | 3.0578951498 | 6.1 |
| P078 | CG | 2.3 | 17.9 | 4.41507465467 | 6.87 |
| P078 | CG | 4.8 | 26.1 | 4.15925829792 | 6.03 |
| P079 | CG | 0 | 16.5 | 7.47540880387 | |
| P079 | CG | 5.6 | 16.3 | 6.83590469591 | |
| P079 | CG | 7.1 | 20.2 | 5.99768746459 | |
| P080 | CG | 0 | 28.6 | 6.84953664904 | 6.46 |
| P080 | CG | 1.6 | 31.5 | 6.31176682492 | 6.29 |
| P080 | CG | 5.5 | 17.1 | 6.91330256001 | 5.66 |

#### Table S3C. Mineral metabolism and parathyroid hormone

De-identified individual participant data are shown in source-workbook row order. Patient_ID, Group and Time_Month are repeated across Table S3A–F for row-level cross-reference. Blank cells indicate missing values; no imputation was applied. Ca×P is the source-workbook product of Ca and P. Field definitions, units and availability are given in Table S4.

| Patient_ID | Group | Time_Month | Ca | P | Ca×P | PTH |
|---|---|---|---|---|---|---|
| P001 | EG | 0 | 1.74 | 2.66 | 4.6284 | 594.1 |
| P001 | EG | 1.7 | 1.78 | 2.24 | 3.9872 | 791.6 |
| P001 | EG | 2.9 | 1.73 | 1.09 | 1.8857 | 337 |
| P002 | EG | 0 | 2.23 | 2.11 | 4.7053 | 171 |
| P002 | EG | 3.6 | 2.37 | 1.87 | 4.4319 | 188 |
| P002 | EG | 6.7 | 2.4 | 1.5 | 3.6 | 131 |
| P003 | EG | 0 | 3.02 | 1.05 | 3.171 | 286.9 |
| P003 | EG | 5.2 | 2.44 | 1.35 | 3.294 | 383 |
| P003 | EG | 8.8 | 2.47 | 1.21 | 2.9887 | 314 |
| P004 | EG | 0 | 2.33 | 1.2 | 2.796 | 88.2 |
| P004 | EG | 2.2 | 2.11 | 1.7 | 3.587 | 219 |
| P004 | EG | 8.2 | 2.4 | 2.27 | 5.448 | 311 |
| P005 | EG | 0 | 2.16 | 1.41 | 3.0456 | 261 |
| P005 | EG | 3.8 | 2.34 | 0.76 | 1.7784 | 181 |
| P005 | EG | 6.1 | 2.21 | 1.31 | 2.8951 | 299 |
| P006 | EG | 0 | 2.18 | 1.96 | 4.2728 | 530 |
| P006 | EG | 3 | 2.22 | 1.77 | 3.9294 | 485 |
| P006 | EG | 6.8 | 2.07 | 1.55 | 3.2085 | 208 |
| P007 | EG | 0 | 1.96 | 2.26 | 4.4296 | 471 |
| P007 | EG | 2.6 | 2.45 | 0.93 | 2.2785 | 363 |
| P007 | EG | 4.8 | 2.41 | 2.45 | 5.9045 | 710 |
| P008 | EG | 0 | 2.11 | 3.64 | 7.6804 | 1530.5 |
| P008 | EG | 7.9 | 2.09 | 3.76 | 7.8584 | 1860 |
| P008 | EG | 10.9 | 2.26 | 3.42 | 7.7292 | 1421 |
| P009 | EG | 0 | 2.22 | 1.06 | 2.3532 | 136 |
| P009 | EG | 3.8 | 2.93 | 2.01 | 5.8893 | 29.5 |
| P009 | EG | 7.1 | 2.52 | 1.84 | 4.6368 | 93.6 |
| P010 | EG | 0 | 2 | 2.75 | 5.5 | 522 |
| P010 | EG | 1.4 | 2.19 | 1.96 | 4.2924 | 502 |
| P010 | EG | 6.8 | 2.02 | 2.16 | 4.3632 | 289 |
| P011 | EG | 0 | 2.6 | 2.45 | 6.37 | 374.2 |
| P011 | EG | 5.2 | 2.48 | 2.77 | 6.8696 | 232 |
| P011 | EG | 8 | 2.79 | 2.77 | 7.7283 | 108 |
| P012 | EG | 0 | 2.05 | 1.65 | 3.3825 | 256.6 |
| P012 | EG | 6.5 | 1.83 | 1.6 | 2.928 | 120 |
| P012 | EG | 9.8 | 2.04 | 1.7 | 3.468 | 276 |
| P013 | EG | 0 | 2.64 | 1.78 | 4.6992 | 146 |
| P013 | EG | 3.4 | 2.69 | 2.12 | 5.7028 | 48.3 |
| P013 | EG | 5.8 | 2.51 | 2.35 | 5.8985 | 201 |
| P014 | EG | 0 | 2.56 | 2.43 | 6.2208 | 505 |
| P014 | EG | 3.2 | 2.47 | 2.64 | 6.5208 | 916 |
| P014 | EG | 5.2 | 2.67 | 3.12 | 8.3304 | 798 |
| P015 | EG | 0 | 2 | 3.37 | 6.74 | 83.5 |
| P015 | EG | 1.5 | 2.09 | 2.12 | 4.4308 | 58.5 |
| P015 | EG | 3.9 | 2.14 | 2.79 | 5.9706 | 114 |
| P016 | EG | 0 | 1.56 | 2.72 | 4.2432 | 471 |
| P016 | EG | 3.1 | 2.04 | 1.93 | 3.9372 | 528 |
| P016 | EG | 5.4 | 1.88 | 2.61 | 4.9068 | 643 |
| P017 | EG | 0 | 2.35 | 1.1 | 2.585 | 258 |
| P017 | EG | 2.8 | 2.4 | 1.65 | 3.96 | 255 |
| P017 | EG | 6.1 | 2.53 | 1.8 | 4.554 | 403 |
| P018 | EG | 0 | 2.03 | 1.19 | 2.4157 | 320 |

| | | | | | | |
|---|---|---|---|---|---|---|
| P018 | EG | 1.6 | 1.93 | 0.96 | 1.8528 | 308 |
| P018 | EG | 6.1 | 2.18 | 1.27 | 2.7686 | 234 |
| P019 | EG | 0 | 2.29 | 1.59 | 3.6411 | 237 |
| P019 | EG | 4.7 | 2.33 | 1.44 | 3.3552 | 259 |
| P019 | EG | 9.6 | 2.42 | 1.35 | 3.267 | 176 |
| P020 | EG | 0 | 2.25 | 3.36 | 7.56 | |
| P020 | EG | 2.9 | 2.4 | 3.02 | 7.248 | 1462 |
| P020 | EG | 5.7 | 2.32 | 2.87 | 6.6584 | 1229 |
| P021 | EG | 0 | 2.22 | 0.85 | 1.887 | 200 |
| P021 | EG | 2.7 | 2.3 | 0.87 | 2.001 | 174 |
| P021 | EG | 6.6 | 2.22 | 1.28 | 2.8416 | 272 |
| P022 | EG | 0 | 2.14 | 2.68 | 5.7352 | 788 |
| P022 | EG | 2 | 2.11 | 1.96 | 4.1356 | 847 |
| P022 | EG | 6.7 | 2.39 | 2.57 | 6.1423 | 737 |
| P023 | EG | 0 | 2.42 | 1.57 | 3.7994 | |
| P023 | EG | 2.5 | 2.2 | 0.77 | 1.694 | 321 |
| P023 | EG | 6.4 | 2.12 | 1.63 | 3.4556 | 369 |
| P024 | EG | 0 | 2.26 | 1.85 | 4.181 | 272 |
| P024 | EG | 4.1 | 2.36 | 2.23 | 5.2628 | 304 |
| P024 | EG | 6.1 | 2.17 | 3.03 | 6.5751 | 459 |
| P025 | EG | 0 | 2.15 | 2 | 4.3 | 294 |
| P025 | EG | 2.8 | 2.41 | 1.67 | 4.0247 | 377 |
| P025 | EG | 6.1 | 2.64 | 1.97 | 5.2008 | 249 |
| P026 | EG | 0 | 1.95 | 1.91 | 3.7245 | 382 |
| P026 | EG | 4.6 | 2.01 | 2.18 | 4.3818 | 574 |
| P026 | EG | 8.1 | 2.02 | 1.58 | 3.1916 | 386 |
| P027 | EG | 0 | 2.42 | 1.76 | 4.2592 | 539 |
| P027 | EG | 3.2 | 2.41 | 2.5 | 6.025 | 773 |
| P027 | EG | 6.3 | 2.46 | 2.4 | 5.904 | 988 |
| P028 | EG | 0 | 2.3 | 2.35 | 5.405 | 592 |
| P028 | EG | 2 | 2.63 | 2.94 | 7.7322 | 420 |
| P028 | EG | 5.3 | 2.78 | 2.43 | 6.7554 | 326 |
| P029 | EG | 0 | 2.12 | 2.38 | 5.0456 | 740 |
| P029 | EG | 1.5 | 1.81 | 3.42 | 6.1902 | 975 |
| P029 | EG | 6.2 | 1.97 | 3.22 | 6.3434 | 1210 |
| P030 | EG | 0 | 2.45 | 2.06 | 5.047 | 56.8 |
| P030 | EG | 3 | 2.38 | 2.2 | 5.236 | 72 |
| P030 | EG | 8.1 | 2.43 | 2.3 | 5.589 | 149 |
| P031 | EG | 0 | 1.81 | 2.35 | 4.2535 | 393.8 |
| P032 | EG | 0 | 1.97 | 0.34 | 0.6698 | 99.9 |
| P032 | EG | 2.6 | 2.38 | 0.71 | 1.6898 | 79.2 |
| P032 | EG | 5.6 | 2.16 | 1 | 2.16 | 135 |
| P033 | EG | 0 | 2 | 2.31 | 4.62 | 302 |
| P033 | EG | 2.8 | 2.09 | 2.92 | 6.1028 | 326 |
| P033 | EG | 6.6 | 1.99 | 2.4 | 4.776 | 372 |
| P034 | EG | 0 | 2.2 | 3.38 | 7.436 | 1004 |
| P034 | EG | 1.2 | 2.41 | 3.2 | 7.712 | |
| P035 | EG | 0 | 2.43 | 1.93 | 4.6899 | 347 |
| P035 | EG | 4 | 2.41 | 1.83 | 4.4103 | 404 |
| P035 | EG | 7.3 | 2.61 | 1.39 | 3.6279 | 244 |
| P036 | EG | 0 | 1.84 | 2.08 | 3.8272 | 165.6 |
| P036 | EG | 5.1 | 1.84 | 1.72 | 3.1648 | 162 |
| P036 | EG | 9 | 1.88 | 2.16 | 4.0608 | 180 |
| P037 | EG | 0 | 2.21 | 3.17 | 7.0057 | 909 |
| P037 | EG | 2.2 | 2.36 | 2.09 | 4.9324 | 375 |
| P037 | EG | 6.5 | 2.14 | 2.39 | 5.1146 | 1014 |
| P038 | EG | 0 | 2.06 | 1.65 | 3.399 | 560 |
| P038 | EG | 5.4 | 2.23 | 1.78 | 3.9694 | 356 |

| | | | | | | |
|---|---|---|---|---|---|---|
| P038 | EG | 7 | 2.11 | 1.23 | 2.5953 | 447 |
| P039 | EG | 0 | 1.82 | 1.89 | 3.4398 | 336 |
| P039 | EG | 1.5 | 1.59 | 2.88 | 4.5792 | 266 |
| P039 | EG | 6.2 | 1.85 | 1.71 | 3.1635 | 221 |
| P040 | EG | 0 | 1.86 | 2.73 | 5.0778 | 585 |
| P040 | EG | 1.8 | 2.08 | 1.92 | 3.9936 | 557 |
| P040 | EG | 3.9 | 2.15 | 1.57 | 3.3755 | 503 |
| P041 | CG | 0 | 2.45 | 1.3 | 3.185 | 170 |
| P041 | CG | 4.8 | 2.25 | 1.88 | 4.23 | 222 |
| P041 | CG | 9.3 | 2.24 | 1.36 | 3.0464 | 166 |
| P042 | CG | 0 | 2.05 | 1.7 | 3.485 | |
| P042 | CG | 4.2 | 2.35 | 1.67 | 3.9245 | |
| P042 | CG | 5.9 | 2.02 | 1.1 | 2.222 | 13.5 |
| P043 | CG | 0 | 1.85 | 3.18 | 5.883 | 873 |
| P043 | CG | 3.6 | 2.13 | 2.56 | 5.4528 | 1036 |
| P043 | CG | 5.1 | 2.15 | 2.57 | 5.5255 | |
| P044 | CG | 0 | 2.08 | 2.02 | 4.2016 | 153 |
| P044 | CG | 5.1 | 2.19 | 1.63 | 3.5697 | 163 |
| P044 | CG | 8.8 | 2.26 | 2.38 | 5.3788 | 282 |
| P045 | CG | 0 | 2.18 | 2.65 | 5.777 | 197 |
| P045 | CG | 2.1 | 2.02 | 1.78 | 3.5956 | 208 |
| P045 | CG | 6.1 | 2.41 | 1.74 | 4.1934 | 243 |
| P046 | CG | 0 | 1.88 | 1.52 | 2.8576 | |
| P046 | CG | 1.8 | 1.9 | 1.58 | 3.002 | |
| P046 | CG | 7 | 1.65 | 1.53 | 2.5245 | |
| P047 | CG | 0 | 2.61 | 2.21 | 5.7681 | 513 |
| P047 | CG | 5.3 | 2.61 | 2.21 | 5.7681 | 1115 |
| P047 | CG | 10.1 | 2.38 | 1.86 | 4.4268 | 475 |
| P048 | CG | 0 | 1.97 | 2.4 | 4.728 | 332 |
| P048 | CG | 4.5 | 1.9 | 2.46 | 4.674 | 348 |
| P048 | CG | 7.2 | 2.09 | 1.71 | 3.5739 | 466 |
| P049 | CG | 0 | 1.85 | 2.26 | 4.181 | 168 |
| P049 | CG | 5.5 | 1.69 | 1.88 | 3.1772 | 176 |
| P049 | CG | 7.8 | 1.68 | 1.74 | 2.9232 | 190 |
| P050 | CG | 0 | 1.79 | 2.14 | 3.8306 | 255 |
| P050 | CG | 4.6 | 1.96 | 2.91 | 5.7036 | |
| P050 | CG | 6.4 | 2 | 3.03 | 6.06 | |
| P051 | CG | 0 | 2.38 | 1.07 | 2.5466 | 155 |
| P051 | CG | 3.3 | 2.4 | 1.05 | 2.52 | 158 |
| P051 | CG | 8.7 | 2.21 | 1.53 | 3.3813 | 231 |
| P052 | CG | 0 | 2.16 | 1.7 | 3.672 | 504.2 |
| P052 | CG | 3.4 | 2.19 | 1.83 | 4.0077 | 421 |
| P052 | CG | 6.2 | 2.15 | 1.46 | 3.139 | 350 |
| P053 | CG | 0 | 2.07 | 1.83 | 3.7881 | 157 |
| P053 | CG | 2.2 | 2.23 | 1.74 | 3.8802 | 125 |
| P053 | CG | 7.8 | 2.43 | 1.46 | 3.5478 | 38 |
| P054 | CG | 0 | 2.5 | 2.11 | 5.275 | 17.3 |
| P054 | CG | 1.5 | 2.52 | 1.92 | 4.8384 | 13.6 |
| P054 | CG | 3.7 | 2.5 | 2.21 | 5.525 | 15.2 |
| P055 | CG | 0 | 2 | 2.67 | 5.34 | 18.1 |
| P055 | CG | 5 | 1.58 | 2.67 | 4.2186 | |
| P055 | CG | 9.7 | 2.06 | 2.79 | 5.7474 | 364 |
| P056 | CG | 0 | 1.65 | 2.24 | 3.696 | 367 |
| P056 | CG | 2.5 | 2.15 | 2.54 | 5.461 | 518 |
| P057 | CG | 0 | 2.38 | 2.08 | 4.9504 | |
| P057 | CG | 3.9 | 2.24 | 1.63 | 3.6512 | 434 |
| P057 | CG | 9 | 2.37 | 2.33 | 5.5221 | 757 |
| P058 | CG | 0 | 1.85 | 0.99 | 1.8315 | 570 |

| | | | | | | |
|---|---|---|---|---|---|---|
| P058 | CG | 1.5 | 1.79 | 1.72 | 3.0788 | 341 |
| P058 | CG | 5.1 | 1.87 | 2.05 | 3.8335 | 579 |
| P059 | CG | 0 | 2.37 | 1.75 | 4.1475 | 222 |
| P059 | CG | 1.5 | 2.35 | 1.4 | 3.29 | 195 |
| P059 | CG | 3.3 | 2.18 | 2.06 | 4.4908 | 253 |
| P060 | CG | 0 | 2.43 | 2.94 | 7.1442 | 173 |
| P060 | CG | 1.7 | 2.24 | 1.58 | 3.5392 | 129 |
| P060 | CG | 5.5 | 2.38 | 2.34 | 5.5692 | 162 |
| P061 | CG | 0 | 2.18 | 2.49 | 5.4282 | 138 |
| P061 | CG | 4.4 | 2.3 | 1.32 | 3.036 | 84.4 |
| P061 | CG | 7.7 | 2.23 | 2.08 | 4.6384 | 101 |
| P062 | CG | 0 | 2.47 | 2.8 | 6.916 | 433 |
| P062 | CG | 5.2 | 2.67 | 2.03 | 5.4201 | 400 |
| P062 | CG | 9.6 | 2.5 | 2.37 | 5.925 | 447 |
| P063 | CG | 0 | 2.18 | 1.12 | 2.4416 | |
| P063 | CG | 1.9 | 2.27 | 1.07 | 2.4289 | 58.5 |
| P063 | CG | 3.5 | 2.31 | 1.31 | 3.0261 | |
| P064 | CG | 0 | 2.09 | 1.63 | 3.4067 | |
| P064 | CG | 4.7 | 1.95 | 1.97 | 3.8415 | 219 |
| P064 | CG | 9.7 | 2 | 1.64 | 3.28 | 249 |
| P065 | CG | 0 | 1.74 | 2.56 | 4.4544 | 305 |
| P065 | CG | 2.7 | 2.09 | 2.86 | 5.9774 | 391 |
| P065 | CG | 7 | 2.01 | 3.17 | 6.3717 | 433 |
| P066 | CG | 0 | 2.22 | 2.26 | 5.0172 | 487.8 |
| P066 | CG | 3.5 | 2.02 | 1.14 | 2.3028 | 364 |
| P066 | CG | 5.9 | 1.81 | 1.99 | 3.6019 | 329 |
| P067 | CG | 0 | 2.4 | 1.02 | 2.448 | 32.5 |
| P067 | CG | 2.4 | 2.26 | 0.8 | 1.808 | 90.1 |
| P067 | CG | 4.5 | 2.21 | 1.06 | 2.3426 | 172 |
| P068 | CG | 0 | 2.12 | 0.83 | 1.7596 | 98.3 |
| P068 | CG | 2.8 | 2.16 | 0.85 | 1.836 | 82 |
| P068 | CG | 4.5 | 2.38 | 1.01 | 2.4038 | 108 |
| P069 | CG | 0 | 2.05 | 2.65 | 5.4325 | 108 |
| P069 | CG | 5.8 | 2.12 | 0.93 | 1.9716 | 91.6 |
| P069 | CG | 9.2 | 2.18 | 1.5 | 3.27 | 99.1 |
| P070 | CG | 0 | 2.17 | 2.11 | 4.5787 | 1219 |
| P070 | CG | 4 | 2.28 | 1.64 | 3.7392 | 1084 |
| P070 | CG | 8.2 | 2.01 | 1.62 | 3.2562 | 599 |
| P071 | CG | 0 | 2.45 | 0.98 | 2.401 | 94.7 |
| P071 | CG | 4.1 | 2.64 | 2.02 | 5.3328 | 71.3 |
| P071 | CG | 7.3 | 3.18 | 1.06 | 3.3708 | 25.7 |
| P072 | CG | 0 | 2.31 | 1.31 | 3.0261 | 55.4 |
| P072 | CG | 1.6 | 2.38 | 1.15 | 2.737 | 90.8 |
| P072 | CG | 5.1 | 2.52 | 0.47 | 1.1844 | 21.2 |
| P073 | CG | 0 | 1.96 | 1.7 | 3.332 | 148 |
| P073 | CG | 3 | 1.96 | 1.57 | 3.0772 | 124 |
| P073 | CG | 4.5 | 1.93 | 2.1 | 4.053 | 50 |
| P074 | CG | 0 | 2.43 | 1.99 | 4.8357 | 182 |
| P074 | CG | 5.9 | 2.62 | 1.12 | 2.9344 | 228 |
| P074 | CG | 8.5 | 2.42 | 1.35 | 3.267 | 176 |
| P075 | CG | 0 | 2.08 | 1.37 | 2.8496 | |
| P075 | CG | 3.8 | 1.92 | 1.88 | 3.6096 | 416 |
| P075 | CG | 8.6 | 1.73 | 1.84 | 3.1832 | 213 |
| P076 | CG | 0 | 2.22 | 2.76 | 6.1272 | 2027 |
| P076 | CG | 2.6 | 2.33 | 2.46 | 5.7318 | 2166 |
| P076 | CG | 7.2 | 1.99 | 2.43 | 4.8357 | |
| P077 | CG | 0 | 1.84 | 2.48 | 4.5632 | 360 |
| P077 | CG | 3.1 | 1.95 | 2.38 | 4.641 | 354 |

| | | | | | | |
|---|---|---|---|---|---|---|
| P077 | CG | 8.1 | 2.04 | 1.87 | 3.8148 | 298 |
| P078 | CG | 0 | 2.1 | 2.9 | 6.09 | 561 |
| P078 | CG | 2.3 | 2.17 | 2.16 | 4.6872 | 526 |
| P078 | CG | 4.8 | 2.12 | 2.15 | 4.558 | 426 |
| P079 | CG | 0 | 1.93 | 2.01 | 3.8793 | 361 |
| P079 | CG | 5.6 | 1.86 | 1.78 | 3.3108 | 250 |
| P079 | CG | 7.1 | 1.76 | 2.19 | 3.8544 | 351 |
| P080 | CG | 0 | 2.01 | 1.88 | 3.7788 | 312 |
| P080 | CG | 1.6 | 1.82 | 2.26 | 4.1132 | 303 |
| P080 | CG | 5.5 | 2.08 | 0.78 | 1.6224 | 156 |

#### Table S3D. Haematological fields

De-identified individual participant data are shown in source-workbook row order. Patient_ID, Group and Time_Month are repeated across Table S3A–F for row-level cross-reference. Blank cells indicate missing values; no imputation was applied. Ca×P is the source-workbook product of Ca and P. Field definitions, units and availability are given in Table S4.

| Patient_ID | Group | Time_Month | NEUT | LYMPH | WBC | Hb |
|---|---|---|---|---|---|---|
| P001 | EG | 0 | 4.53 | 1.23 | 6.37 | 116 |
| P001 | EG | 1.7 | 2.95 | 1.15 | 4.64 | 119 |
| P001 | EG | 2.9 | 3.81 | 1.4 | 5.78 | 130 |
| P002 | EG | 0 | 6.45 | 1.09 | 8.35 | 118 |
| P002 | EG | 3.6 | 6.96 | 1.27 | 9.04 | 113 |
| P002 | EG | 6.7 | 5.34 | 0.9 | 6.97 | 107 |
| P003 | EG | 0 | 4.79 | 0.84 | 6.59 | 106 |
| P003 | EG | 5.2 | 4.8 | 0.82 | 6.58 | 103 |
| P003 | EG | 8.8 | 5.77 | 0.86 | 7.76 | 110 |
| P004 | EG | 0 | 3.41 | 1.1 | 5.07 | 102 |
| P004 | EG | 2.2 | 3.33 | 0.79 | 4.68 | 120 |
| P004 | EG | 8.2 | 2.97 | 1.16 | 4.77 | 106 |
| P005 | EG | 0 | 2.99 | 0.88 | 4.52 | 132 |
| P005 | EG | 3.8 | 3.89 | 1.24 | 5.85 | 110 |
| P005 | EG | 6.1 | 3.86 | 1 | 5.39 | 106 |
| P006 | EG | 0 | 4.43 | 0.95 | 5.77 | 67 |
| P006 | EG | 3 | 3.48 | 0.93 | 4.78 | 81 |
| P006 | EG | 6.8 | 4.78 | 0.75 | 5.85 | 91 |
| P007 | EG | 0 | 3.53 | 0.58 | 4.49 | 122 |
| P007 | EG | 2.6 | 2.25 | 0.59 | 3.21 | 96 |
| P007 | EG | 4.8 | 3.68 | 0.56 | 4.54 | 119 |
| P008 | EG | 0 | 3.95 | 1.08 | 5.76 | 92 |
| P008 | EG | 7.9 | 5.64 | 0.94 | 7.25 | 92 |
| P008 | EG | 10.9 | 4.9 | 1.45 | 7.13 | 102 |
| P009 | EG | 0 | 3 | 0.89 | 4.82 | 107 |
| P009 | EG | 3.8 | 3.43 | 0.62 | 5.67 | 99 |
| P009 | EG | 7.1 | 3.03 | 0.77 | 4.85 | 102 |
| P010 | EG | 0 | 6.82 | 1.27 | 8.62 | 138 |
| P010 | EG | 1.4 | 4.04 | 1.4 | 5.79 | 125 |
| P010 | EG | 6.8 | 4.54 | 0.98 | 5.94 | 117 |
| P011 | EG | 0 | 3.27 | 1.25 | 4.87 | 121 |
| P011 | EG | 5.2 | 3.32 | 0.76 | 4.46 | 99 |
| P011 | EG | 8 | 2.9 | 0.71 | 3.9 | 131 |
| P012 | EG | 0 | 5.09 | 1.69 | 7.66 | 110 |
| P012 | EG | 6.5 | 3.54 | 1.27 | 5.75 | 104 |
| P012 | EG | 9.8 | 3.52 | 1.6 | 5.89 | 111 |
| P013 | EG | 0 | 2.71 | 1.01 | 4.53 | 143 |
| P013 | EG | 3.4 | 3.03 | 0.96 | 4.3 | 141 |
| P013 | EG | 5.8 | 4.2 | 0.77 | 5.3 | 118 |
| P014 | EG | 0 | 4.73 | 1.39 | 6.76 | 91 |
| P014 | EG | 3.2 | 4.44 | 1.17 | 6.07 | 121 |
| P014 | EG | 5.2 | 4.79 | 1.29 | 6.6 | 108 |
| P015 | EG | 0 | 2.54 | 1.33 | 4.48 | 139 |
| P015 | EG | 1.5 | 2.96 | 1.82 | 5.69 | 121 |
| P015 | EG | 3.9 | 4.41 | 1.57 | 6.83 | 124 |
| P016 | EG | 0 | 4.72 | 0.85 | 6 | 115 |
| P016 | EG | 3.1 | 3.51 | 0.83 | 4.7 | 131 |
| P016 | EG | 5.4 | 4.69 | 0.85 | 5.99 | 114 |
| P017 | EG | 0 | 3.85 | 1.03 | 5.93 | 103 |
| P017 | EG | 2.8 | 6.81 | 1.27 | 9.89 | 129 |
| P017 | EG | 6.1 | 5.89 | 1.02 | 7.99 | 105 |
| P018 | EG | 0 | 2.94 | 0.85 | 4.16 | 122 |

| | | | | | | |
|---|---|---|---|---|---|---|
| P018 | EG | 1.6 | 3.7 | 0.8 | 4.93 | 117 |
| P018 | EG | 6.1 | 3.23 | 0.79 | 4.43 | 124 |
| P019 | EG | 0 | 3.07 | 2.29 | 6 | 116 |
| P019 | EG | 4.7 | 4.06 | 1.92 | 6.62 | 103 |
| P019 | EG | 9.6 | 3.01 | 40.2 | 6.24 | 109 |
| P020 | EG | 0 | 6.48 | 0.86 | 8.09 | 127 |
| P020 | EG | 2.9 | 4.17 | 0.32 | 4.73 | 98 |
| P020 | EG | 5.7 | 2.95 | 0.46 | 3.89 | 73 |
| P021 | EG | 0 | 3.31 | 1.5 | 5.15 | 121 |
| P021 | EG | 2.7 | 3.72 | 1.39 | 5.53 | 132 |
| P021 | EG | 6.6 | 4.15 | 1.52 | 6.13 | 115 |
| P022 | EG | 0 | 4.72 | 1.23 | 6.71 | 111 |
| P022 | EG | 2 | 2.99 | 0.99 | 4.76 | 115 |
| P022 | EG | 6.7 | 3.65 | 1.11 | 5.47 | 106 |
| P023 | EG | 0 | 5.36 | 1.73 | 7.72 | 132 |
| P023 | EG | 2.5 | 4.58 | 2.37 | 7.67 | 121 |
| P023 | EG | 6.4 | 3.63 | 1.5 | 5.81 | 107 |
| P024 | EG | 0 | 6.17 | 0.77 | 7.48 | 113 |
| P024 | EG | 4.1 | 7.5 | 0.97 | 9.07 | 125 |
| P024 | EG | 6.1 | 5.41 | 0.57 | 6.57 | 95 |
| P025 | EG | 0 | 3.92 | 1 | 5.38 | 114 |
| P025 | EG | 2.8 | 6.24 | 0.92 | 7.68 | 116 |
| P025 | EG | 6.1 | 4.12 | 1.05 | 5.65 | 107 |
| P026 | EG | 0 | 3.07 | 1.35 | 4.76 | 159 |
| P026 | EG | 4.6 | 3.18 | 1.04 | 4.45 | 116 |
| P026 | EG | 8.1 | 2.83 | 1.14 | 4.27 | 123 |
| P027 | EG | 0 | 4.76 | 1.4 | 6.77 | 106 |
| P027 | EG | 3.2 | 4.1 | 1.53 | 6.28 | 122 |
| P027 | EG | 6.3 | 3.8 | 1.5 | 5.95 | 121 |
| P028 | EG | 0 | 2.29 | 1.05 | 3.67 | 82 |
| P028 | EG | 2 | 3.07 | 1.35 | 5.16 | 93 |
| P028 | EG | 5.3 | 2.98 | 0.97 | 4.63 | 101 |
| P029 | EG | 0 | 6.28 | 1.36 | 8.22 | 96 |
| P029 | EG | 1.5 | 8.96 | 1.49 | 11.12 | 85 |
| P029 | EG | 6.2 | 8.38 | 1.31 | 10.38 | 99 |
| P030 | EG | 0 | 10.94 | 1.65 | 13.5 | 104 |
| P030 | EG | 3 | 11.92 | 1.54 | 14.4 | 136 |
| P030 | EG | 8.1 | 9.37 | 1.85 | 12.09 | 83 |
| P031 | EG | 0 | 2.53 | 0.99 | 3.95 | 82 |
| P032 | EG | 0 | 4.37 | 0.8 | 5.56 | 111 |
| P032 | EG | 2.6 | 5.84 | 1.1 | 7.42 | 122 |
| P032 | EG | 5.6 | 5.4 | 1.07 | 6.95 | 125 |
| P033 | EG | 0 | 3.4 | 1.05 | 5.2 | 102 |
| P033 | EG | 2.8 | 3.03 | 0.91 | 4.6 | 102 |
| P033 | EG | 6.6 | 2.87 | 0.99 | 4.64 | 102 |
| P034 | EG | 0 | 4.6 | 0.52 | 5.59 | 70 |
| P034 | EG | 1.2 | 8.63 | 0.71 | 10.27 | 90 |
| P035 | EG | 0 | 3.73 | 0.76 | 4.87 | 127 |
| P035 | EG | 4 | 3.24 | 0.56 | 4.03 | 106 |
| P035 | EG | 7.3 | 2.51 | 0.5 | 3.28 | 87 |
| P036 | EG | 0 | 8.65 | 1.33 | 11.3 | 70 |
| P036 | EG | 5.1 | 6.61 | 1.42 | 8.89 | 88 |
| P036 | EG | 9 | 6.2 | 1.35 | 8.68 | 95 |
| P037 | EG | 0 | 8 | 1.05 | 9.81 | 113 |
| P037 | EG | 2.2 | 7.8 | 1.35 | 10.1 | 107 |
| P037 | EG | 6.5 | 8.48 | 0.86 | 10.01 | 95 |
| P038 | EG | 0 | 4.83 | 0.67 | 6.87 | 66 |
| P038 | EG | 5.4 | 4.12 | 0.94 | 6.42 | 94 |

| | | | | | | |
|---|---|---|---|---|---|---|
| P038 | EG | 7 | 4.53 | 0.83 | 6.72 | 92 |
| P039 | EG | 0 | 2.61 | 1.32 | 5.02 | 4.7 |
| P039 | EG | 1.5 | 5.82 | 0.69 | 7.09 | 117 |
| P039 | EG | 6.2 | 3.49 | 0.75 | 4.9 | 92 |
| P040 | EG | 0 | 2.87 | 0.72 | 4.19 | 119 |
| P040 | EG | 1.8 | 3.83 | 1.01 | 5.22 | 131 |
| P040 | EG | 3.9 | 3.13 | 0.85 | 4.4 | 97 |
| P041 | CG | 0 | 3.49 | 1.38 | 5.33 | 101 |
| P041 | CG | 4.8 | 2.16 | 1.16 | 3.79 | 91 |
| P041 | CG | 9.3 | 2.43 | 0.95 | 3.77 | 100 |
| P042 | CG | 0 | 3.21 | 1.73 | 5.96 | 102 |
| P042 | CG | 4.2 | 8.39 | 1.8 | 12.04 | 92 |
| P042 | CG | 5.9 | 3.86 | 1.91 | 6.34 | 77 |
| P043 | CG | 0 | 6.96 | 1.31 | 9.13 | 81 |
| P043 | CG | 3.6 | 4.1 | 0.33 | 5.46 | 83 |
| P043 | CG | 5.1 | 5.71 | 1.16 | 7.73 | 112 |
| P044 | CG | 0 | 4.17 | 1.31 | 6.21 | 126 |
| P044 | CG | 5.1 | 3.17 | 1.24 | 5.11 | 109 |
| P044 | CG | 8.8 | 3.23 | 1.01 | 4.72 | 128 |
| P045 | CG | 0 | 6.36 | 1.02 | 8.05 | 88 |
| P045 | CG | 2.1 | 3.13 | 1.27 | 4.87 | 112 |
| P045 | CG | 6.1 | 4.14 | 1.18 | 5.92 | 121 |
| P046 | CG | 0 | 5.73 | 2.35 | 9.89 | 107 |
| P046 | CG | 1.8 | 6.12 | 2.7 | 10.23 | 101 |
| P046 | CG | 7 | 6.17 | 1.88 | 9.15 | 103 |
| P047 | CG | 0 | 2.13 | 1.18 | 3.57 | 114 |
| P047 | CG | 5.3 | 2.13 | 1.18 | 3.75 | 116 |
| P047 | CG | 10.1 | 2.1 | 1.17 | 3.48 | 105 |
| P048 | CG | 0 | 2.27 | 1.01 | 6.07 | 118 |
| P048 | CG | 4.5 | 2.89 | 0.93 | 6.9 | 117 |
| P048 | CG | 7.2 | 3.11 | 0.86 | 6.4 | 123 |
| P049 | CG | 0 | 5.41 | 1.8 | 7.85 | 121 |
| P049 | CG | 5.5 | 3.68 | 1.58 | 5.81 | 114 |
| P049 | CG | 7.8 | 6.09 | 1.46 | 8.27 | 114 |
| P050 | CG | 0 | 3.52 | 0.88 | 4.63 | 103 |
| P050 | CG | 4.6 | 3.88 | 1.07 | 5.53 | 78 |
| P050 | CG | 6.4 | 3.66 | 0.96 | 4.93 | 98 |
| P051 | CG | 0 | 2.46 | 0.87 | 3.69 | 103 |
| P051 | CG | 3.3 | 2.89 | 1.14 | 4.47 | 104 |
| P051 | CG | 8.7 | 3.24 | 0.74 | 4.37 | 105 |
| P052 | CG | 0 | 5.49 | 1.49 | 7.52 | 128 |
| P052 | CG | 3.4 | 4.89 | 1.63 | 7.01 | 127 |
| P052 | CG | 6.2 | 5.89 | 1.69 | 8.2 | 126 |
| P053 | CG | 0 | 6.53 | 167 | 9.1 | 100 |
| P053 | CG | 2.2 | 6.07 | 1.48 | 8.28 | 97 |
| P053 | CG | 7.8 | 5.66 | 1.47 | 7.78 | 122 |
| P054 | CG | 0 | 6.04 | 2.26 | 9.15 | 127 |
| P054 | CG | 1.5 | 6.13 | 1.87 | 8.97 | 121 |
| P054 | CG | 3.7 | 5.23 | 2.04 | 8.14 | 113 |
| P055 | CG | 0 | 2.44 | 0.54 | 3.25 | 118 |
| P055 | CG | 5 | 2.26 | 0.55 | 3.14 | 117 |
| P055 | CG | 9.7 | 1.82 | 0.48 | 2.52 | 112 |
| P056 | CG | 0 | 3.58 | 1.35 | 5.85 | 106 |
| P056 | CG | 2.5 | 3.54 | 1.17 | 5.31 | 128 |
| P057 | CG | 0 | 3.56 | 1.4 | 5.59 | 90 |
| P057 | CG | 3.9 | 2.21 | 0.56 | 3.17 | 106 |
| P057 | CG | 9 | 3.68 | 1.54 | 5.94 | 109 |
| P058 | CG | 0 | 4.64 | 1.11 | 6.3 | 141 |

| | | | | | | |
|---|---|---|---|---|---|---|
| P058 | CG | 1.5 | 4.11 | 1.34 | 5.98 | 122 |
| P058 | CG | 5.1 | 5.83 | 1.4 | 7.96 | 97 |
| P059 | CG | 0 | 3.58 | 1.22 | 5.37 | 112 |
| P059 | CG | 1.5 | 4.82 | 1.63 | 7.19 | 139 |
| P059 | CG | 3.3 | 3.88 | 1.27 | 6.08 | 140 |
| P060 | CG | 0 | 6.89 | 1.85 | 9.46 | 123 |
| P060 | CG | 1.7 | 6.48 | 1.45 | 8.7 | 112 |
| P060 | CG | 5.5 | 5.15 | 1.47 | 7.4 | 108 |
| P061 | CG | 0 | 4.69 | 1.07 | 6.65 | 145 |
| P061 | CG | 4.4 | 4.59 | 1.31 | 6.88 | 133 |
| P061 | CG | 7.7 | 6.68 | 1.22 | 8.73 | 151 |
| P062 | CG | 0 | 6.58 | 1.45 | 8.4 | 126 |
| P062 | CG | 5.2 | 5.26 | 2.03 | 7.78 | 115 |
| P062 | CG | 9.6 | 5.33 | 1.74 | 7.46 | 109 |
| P063 | CG | 0 | 7.01 | 2.08 | 9.85 | 112 |
| P063 | CG | 1.9 | 5.89 | 2.21 | 8.72 | 106 |
| P063 | CG | 3.5 | 6.23 | 1.98 | 8.81 | 103 |
| P064 | CG | 0 | 4.57 | 1.79 | 6.98 | 89 |
| P064 | CG | 4.7 | 3.44 | 1.12 | 5.07 | 72 |
| P064 | CG | 9.7 | 3.09 | 1.34 | 5.06 | 88 |
| P065 | CG | 0 | 5.45 | 0.37 | 5.9 | 130 |
| P065 | CG | 2.7 | 3.58 | 1.21 | 5.23 | 118 |
| P065 | CG | 7 | 4.07 | 0.83 | 5.27 | 88 |
| P066 | CG | 0 | 3.74 | 1.36 | 5.61 | 125 |
| P066 | CG | 3.5 | 2.77 | 1.5 | 4.81 | 121 |
| P066 | CG | 5.9 | 3.58 | 1.36 | 5.54 | 120 |
| P067 | CG | 0 | 3.58 | 1.45 | 5.76 | 153 |
| P067 | CG | 2.4 | 3.57 | 2.52 | 6.59 | 138 |
| P067 | CG | 4.5 | 4.27 | 2.25 | 7.32 | 93 |
| P068 | CG | 0 | 4.3 | 1.01 | 6.08 | 124 |
| P068 | CG | 2.8 | 4.96 | 0.88 | 6.6 | 114 |
| P068 | CG | 4.5 | 5.59 | 1.11 | 7.56 | 127 |
| P069 | CG | 0 | 3.29 | 1.69 | 5.59 | 146 |
| P069 | CG | 5.8 | 3.3 | 1.4 | 5.19 | 132 |
| P069 | CG | 9.2 | 3.04 | 1.58 | 5.07 | 99 |
| P070 | CG | 0 | 4.32 | 1.33 | 6.34 | 84 |
| P070 | CG | 4 | 4.94 | 1.26 | 6.94 | 92 |
| P070 | CG | 8.2 | 5 | 1.26 | 6.89 | 116 |
| P071 | CG | 0 | 5.18 | 1 | 6.68 | 127 |
| P071 | CG | 4.1 | 4.8 | 0.86 | 6.07 | 121 |
| P071 | CG | 7.3 | 4 | 1.05 | 5.54 | 129 |
| P072 | CG | 0 | 5.15 | 1.14 | 7.06 | 88 |
| P072 | CG | 1.6 | 3.37 | 1.2 | 5.23 | 112 |
| P072 | CG | 5.1 | 3.37 | 1.15 | 5.06 | 115 |
| P073 | CG | 0 | 3.28 | 3.37 | 7.52 | 87 |
| P073 | CG | 3 | 3.96 | 2.41 | 7.04 | 96 |
| P073 | CG | 4.5 | 5.28 | 1.42 | 7.42 | 90 |
| P074 | CG | 0 | 3.84 | 1.97 | 6.28 | 133 |
| P074 | CG | 5.9 | 3.85 | 1.99 | 6.37 | 114 |
| P074 | CG | 8.5 | 3.01 | 2.51 | 6.24 | 109 |
| P075 | CG | 0 | 2.85 | 1.86 | 5.37 | 100 |
| P075 | CG | 3.8 | 3.25 | 1.49 | 5.6 | 85 |
| P075 | CG | 8.6 | 2.47 | 0.66 | 3.75 | 99 |
| P076 | CG | 0 | 4.71 | 0.42 | 5.72 | 62 |
| P076 | CG | 2.6 | 5.23 | 0.39 | 6.18 | 105 |
| P076 | CG | 7.2 | 4 | 0.7 | 5.29 | 94 |
| P077 | CG | 0 | 4.05 | 1.14 | 5.63 | 78 |
| P077 | CG | 3.1 | 2.61 | 1.18 | 4.19 | 102 |

| | | | | | | |
|---|---|---|---|---|---|---|
| P077 | CG | 8.1 | 4.93 | 1.11 | 6.44 | 110 |
| P078 | CG | 0 | 3.62 | 0.69 | 4.6 | 97 |
| P078 | CG | 2.3 | 1.46 | 0.61 | 2.35 | 97 |
| P078 | CG | 4.8 | 3.23 | 0.63 | 4.26 | 78 |
| P079 | CG | 0 | 5.38 | 1.88 | 14.22 | 112 |
| P079 | CG | 5.6 | 6.06 | 2.06 | 12.56 | 110 |
| P079 | CG | 7.1 | 5.39 | 1.7 | 10.68 | 104 |
| P080 | CG | 0 | 2.77 | 1.09 | 4.56 | 89 |
| P080 | CG | 1.6 | 2.33 | 0.66 | 3.51 | 87 |
| P080 | CG | 5.5 | 1.91 | 0.73 | 3.07 | 89 |

#### Table S3E. Hepatic laboratory fields

De-identified individual participant data are shown in source-workbook row order. Patient_ID, Group and Time_Month are repeated across Table S3A–F for row-level cross-reference. Blank cells indicate missing values; no imputation was applied. Ca×P is the source-workbook product of Ca and P. Field definitions, units and availability are given in Table S4.

| Patient_ID | Group | Time_Month | AST | ALT | CHE | ALP |
|---|---|---|---|---|---|---|
| P001 | EG | 0 | 6 | 10 | 7040 | 113 |
| P001 | EG | 1.7 | 11 | 24 | 5963 | 120 |
| P001 | EG | 2.9 | 9 | 14 | 6731 | 111 |
| P002 | EG | 0 | 7 | 8 | 7077 | 42 |
| P002 | EG | 3.6 | 12 | 9 | 6993 | 51 |
| P002 | EG | 6.7 | 8 | 10 | 6988 | 49 |
| P003 | EG | 0 | 14 | 13 | 4592 | 326 |
| P003 | EG | 5.2 | 21 | 18 | 4172 | 341 |
| P003 | EG | 8.8 | 16 | 5 | 3766 | 346 |
| P004 | EG | 0 | 8 | 10 | 7989 | 39 |
| P004 | EG | 2.2 | 9 | 16 | 6860 | 43 |
| P004 | EG | 8.2 | 10 | 14 | 6008 | 47 |
| P005 | EG | 0 | 7 | 11 | 5595 | 51 |
| P005 | EG | 3.8 | 3 | 7 | 5614 | 61 |
| P005 | EG | 6.1 | 2 | 5 | 5007 | 55 |
| P006 | EG | 0 | 4 | 5 | 4542 | 46 |
| P006 | EG | 3 | 7 | 7 | 5019 | 50 |
| P006 | EG | 6.8 | 5 | 6 | 4827 | 47 |
| P007 | EG | 0 | 14 | 11 | 6432 | 55 |
| P007 | EG | 2.6 | 12 | 12 | 7709 | 60 |
| P007 | EG | 4.8 | 3 | 8 | 8055 | 65 |
| P008 | EG | 0 | 2 | 6 | 2259 | 82 |
| P008 | EG | 7.9 | 4 | 4 | 1959 | 96 |
| P008 | EG | 10.9 | 5 | 5 | 2107 | 129 |
| P009 | EG | 0 | 10 | 13 | 3984 | 73 |
| P009 | EG | 3.8 | 8 | 8 | 3550 | 117 |
| P009 | EG | 7.1 | 14 | 19 | 4790 | 167 |
| P010 | EG | 0 | 6 | 4 | 5893 | 103 |
| P010 | EG | 1.4 | 5 | 3 | 6208 | 116 |
| P010 | EG | 6.8 | 4 | 4 | 5537 | 122 |
| P011 | EG | 0 | 10 | 6 | 6793 | 33 |
| P011 | EG | 5.2 | 9 | 7 | 5486 | 28 |
| P011 | EG | 8 | 9 | 10 | 6945 | 48 |
| P012 | EG | 0 | 12 | 18 | 5151 | 39 |
| P012 | EG | 6.5 | 13 | 30 | 4660 | 51 |
| P012 | EG | 9.8 | 12 | 20 | 5022 | 61 |
| P013 | EG | 0 | 9 | 14 | 5274 | 53 |
| P013 | EG | 3.4 | 10 | 11 | 5898 | 65 |
| P013 | EG | 5.8 | 6 | 8 | 5631 | 78 |
| P014 | EG | 0 | 13 | 7 | 5883 | 70 |
| P014 | EG | 3.2 | 11 | 7 | 5777 | 85 |
| P014 | EG | 5.2 | 11 | 10 | 5956 | 92 |
| P015 | EG | 0 | 16 | 12 | 5250 | 49 |
| P015 | EG | 1.5 | 17 | 10 | 5278 | 45 |
| P015 | EG | 3.9 | 15 | 9 | 5900 | 58 |
| P016 | EG | 0 | 9 | 6 | 6099 | 63 |
| P016 | EG | 3.1 | 8 | 9 | 6104 | 48 |
| P016 | EG | 5.4 | 9 | 6 | 6311 | 66 |
| P017 | EG | 0 | 16 | 11 | 5586 | 82 |
| P017 | EG | 2.8 | 12 | 14 | 5704 | 72 |
| P017 | EG | 6.1 | 12 | 11 | 6010 | 98 |
| P018 | EG | 0 | 10 | 6 | 6430 | 91 |

| | | | | | | |
|---|---|---|---|---|---|---|
| P018 | EG | 1.6 | 10 | 5 | 6121 | 92 |
| P018 | EG | 6.1 | 16 | 11 | 6645 | 88 |
| P019 | EG | 0 | 13 | 14 | 7072 | 56 |
| P019 | EG | 4.7 | 16 | 25 | 7324 | 56 |
| P019 | EG | 9.6 | 8 | 11 | 7553 | 88 |
| P020 | EG | 0 | 15 | 10 | 5662 | 196 |
| P020 | EG | 2.9 | 11 | 6 | 5097 | 264 |
| P020 | EG | 5.7 | 13 | 12 | 3081 | 157 |
| P021 | EG | 0 | 5 | 8 | 6290 | 52 |
| P021 | EG | 2.7 | 11 | 9 | 6218 | 56 |
| P021 | EG | 6.6 | 10 | 12 | 6595 | 69 |
| P022 | EG | 0 | 10 | 10 | 5858 | 86 |
| P022 | EG | 2 | 9 | 8 | 7283 | 94 |
| P022 | EG | 6.7 | 7 | 14 | 7709 | 101 |
| P023 | EG | 0 | 13 | 7 | 8137 | 37 |
| P023 | EG | 2.5 | 19 | 16 | 7467 | 39 |
| P023 | EG | 6.4 | 15 | 10 | 7044 | 41 |
| P024 | EG | 0 | 2 | 7 | 9282 | 47 |
| P024 | EG | 4.1 | 3 | 9 | 10140 | 64 |
| P024 | EG | 6.1 | 1 | 4 | 8780 | 56 |
| P025 | EG | 0 | 18 | 21 | 8240 | 64 |
| P025 | EG | 2.8 | 15 | 16 | 8265 | 90 |
| P025 | EG | 6.1 | 18 | 18 | 8876 | 82 |
| P026 | EG | 0 | 8 | 7 | 4731 | 77 |
| P026 | EG | 4.6 | 14 | 10 | 4615 | 91 |
| P026 | EG | 8.1 | 14 | 13 | 4793 | 85 |
| P027 | EG | 0 | 7 | 6 | 4932 | 60 |
| P027 | EG | 3.2 | 9 | 3 | 4845 | 79 |
| P027 | EG | 6.3 | 7 | 5 | 5092 | 72 |
| P028 | EG | 0 | 9 | 15 | 2858 | 26 |
| P028 | EG | 2 | 5 | 10 | 2639 | 26 |
| P028 | EG | 5.3 | 7 | 8 | 3073 | 40 |
| P029 | EG | 0 | 7 | 9 | 7925 | 80 |
| P029 | EG | 1.5 | 9 | 9 | 7940 | 85 |
| P029 | EG | 6.2 | 11 | 9 | 7827 | 109 |
| P030 | EG | 0 | 9 | 10 | 6369 | 104 |
| P030 | EG | 3 | | | | |
| P030 | EG | 8.1 | 9 | 5 | 5819 | 76 |
| P031 | EG | 0 | 13 | 9 | 6031 | 78 |
| P032 | EG | 0 | 13 | 7 | 5757 | 40 |
| P032 | EG | 2.6 | 10 | 7 | 7307 | 70 |
| P032 | EG | 5.6 | 14 | 6 | 6903 | 90 |
| P033 | EG | 0 | 2 | 5 | 5752 | 58 |
| P033 | EG | 2.8 | 3 | 2 | 5883 | 55 |
| P033 | EG | 6.6 | 1 | 7 | 5854 | 59 |
| P034 | EG | 0 | 89 | 98 | 5352 | 114 |
| P034 | EG | 1.2 | | 10 | | |
| P035 | EG | 0 | 11 | 24 | 6266 | 99 |
| P035 | EG | 4 | 16 | 10 | 5560 | 78 |
| P035 | EG | 7.3 | 10 | 6 | 5347 | 80 |
| P036 | EG | 0 | 9 | 7 | 5828 | 63 |
| P036 | EG | 5.1 | 8 | 9 | 5331 | 89 |
| P036 | EG | 9 | 8 | 9 | 5878 | 101 |
| P037 | EG | 0 | 5 | 7 | 8084 | 54 |
| P037 | EG | 2.2 | 4 | 9 | 7845 | 52 |
| P037 | EG | 6.5 | 2 | | 7040 | 73 |
| P038 | EG | 0 | 6 | 9 | 4644 | 58 |
| P038 | EG | 5.4 | 10 | 10 | 4898 | 79 |

| | | | | | | |
|---|---|---|---|---|---|---|
| P038 | EG | 7 | 14 | 8 | 4415 | 79 |
| P039 | EG | 0 | 15 | 15 | 6948 | 57 |
| P039 | EG | 1.5 | 10 | 27 | 5815 | 86 |
| P039 | EG | 6.2 | 15 | 31 | 3847 | 99 |
| P040 | EG | 0 | 9 | 16 | 5427 | 54 |
| P040 | EG | 1.8 | 17 | 7 | 6972 | 78 |
| P040 | EG | 3.9 | 8 | 9 | 5414 | 84 |
| P041 | CG | 0 | 15 | 14 | 7752 | 79 |
| P041 | CG | 4.8 | 12 | 10 | 6919 | 59 |
| P041 | CG | 9.3 | 21 | 11 | 6702 | 53 |
| P042 | CG | 0 | 20 | 12 | 7558 | 55 |
| P042 | CG | 4.2 | 13 | 14 | 4946 | 35 |
| P042 | CG | 5.9 | 16 | 12 | 7392 | 31 |
| P043 | CG | 0 | 10 | 6 | 6139 | 75 |
| P043 | CG | 3.6 | 13 | 11 | 5130 | 94 |
| P043 | CG | 5.1 | 14 | 12 | 5000 | 84 |
| P044 | CG | 0 | 9 | 8 | 6962 | 48 |
| P044 | CG | 5.1 | 7 | 7 | 7424 | 46 |
| P044 | CG | 8.8 | | | | |
| P045 | CG | 0 | 11 | 9 | 6379 | 54 |
| P045 | CG | 2.1 | 13 | 12 | 7090 | 60 |
| P045 | CG | 6.1 | 12 | 10 | 6989 | 62 |
| P046 | CG | 0 | 13 | 10 | 10471 | 216 |
| P046 | CG | 1.8 | 12 | 9 | 8780 | 162 |
| P046 | CG | 7 | 13 | 13 | 9486 | 170 |
| P047 | CG | 0 | | | | |
| P047 | CG | 5.3 | | | | |
| P047 | CG | 10.1 | | | | |
| P048 | CG | 0 | 7 | 11 | 5819 | 52 |
| P048 | CG | 4.5 | 10 | 13 | 5757 | 49 |
| P048 | CG | 7.2 | 9 | 12 | 6465 | 65 |
| P049 | CG | 0 | 11 | 10 | 6308 | 58 |
| P049 | CG | 5.5 | 15 | 17 | 5805 | 44 |
| P049 | CG | 7.8 | 15 | 25 | 5703 | 87 |
| P050 | CG | 0 | 16 | 18 | 6094 | 77 |
| P050 | CG | 4.6 | 25 | 22 | 5187 | 69 |
| P050 | CG | 6.4 | 13 | 12 | 5098 | 45 |
| P051 | CG | 0 | 8 | 8 | 4799 | 58 |
| P051 | CG | 3.3 | 14 | 15 | 5105 | 56 |
| P051 | CG | 8.7 | 12 | 8 | 5513 | 52 |
| P052 | CG | 0 | 16 | 18 | 7458 | 76 |
| P052 | CG | 3.4 | 11 | 19 | 8302 | 87 |
| P052 | CG | 6.2 | 15 | 18 | 7651 | 89 |
| P053 | CG | 0 | 7 | 7 | 7778 | 66 |
| P053 | CG | 2.2 | 6 | 9 | 7129 | 49 |
| P053 | CG | 7.8 | 8 | 7 | 7757 | 47 |
| P054 | CG | 0 | 5 | 7 | 8922 | 69 |
| P054 | CG | 1.5 | 4 | 8 | 8393 | 55 |
| P054 | CG | 3.7 | 14 | 12 | 7862 | 44 |
| P055 | CG | 0 | 7 | 12 | 4271 | 90 |
| P055 | CG | 5 | 5 | 9 | 3970 | 68 |
| P055 | CG | 9.7 | 7 | 11 | 4005 | 71 |
| P056 | CG | 0 | 10 | 7 | 7247 | 54 |
| P056 | CG | 2.5 | 6 | 5 | 8061 | 87 |
| P057 | CG | 0 | 4 | 7 | 9606 | 84 |
| P057 | CG | 3.9 | 6 | 8 | 8573 | 97 |
| P057 | CG | 9 | 3 | 9 | 9793 | 91 |
| P058 | CG | 0 | 18 | 23 | 6167 | 130 |

| | | | | | | |
|---|---|---|---|---|---|---|
| P058 | CG | 1.5 | 11 | 16 | 7338 | 111 |
| P058 | CG | 5.1 | 15 | 19 | 7235 | 123 |
| P059 | CG | 0 | 5 | 6 | 4626 | 60 |
| P059 | CG | 1.5 | 3 | 7 | 4785 | 68 |
| P059 | CG | 3.3 | 1 | 4 | 3782 | 68 |
| P060 | CG | 0 | 7 | 6 | 8482 | 59 |
| P060 | CG | 1.7 | 5 | 8 | 8011 | 86 |
| P060 | CG | 5.5 | 4 | 7 | 7919 | 82 |
| P061 | CG | 0 | 10 | 10 | 7836 | 62 |
| P061 | CG | 4.4 | 7 | 6 | 7134 | 71 |
| P061 | CG | 7.7 | 13 | 12 | 7373 | 66 |
| P062 | CG | 0 | 7 | 5 | 5938 | 42 |
| P062 | CG | 5.2 | 6 | 4 | 6207 | 75 |
| P062 | CG | 9.6 | | | | |
| P063 | CG | 0 | | | | |
| P063 | CG | 1.9 | | | | |
| P063 | CG | 3.5 | | | | |
| P064 | CG | 0 | 19 | 10 | 6582 | 76 |
| P064 | CG | 4.7 | 14 | 9 | 4065 | 54 |
| P064 | CG | 9.7 | 18 | 10 | 4720 | 55 |
| P065 | CG | 0 | 7 | 7 | 5501 | 72 |
| P065 | CG | 2.7 | 7 | 11 | 7333 | 56 |
| P065 | CG | 7 | 8 | 8 | 6876 | 57 |
| P066 | CG | 0 | 12 | 7 | 7346 | 98 |
| P066 | CG | 3.5 | 12 | 9 | 7640 | 97 |
| P066 | CG | 5.9 | 12 | 11 | 7293 | 95 |
| P067 | CG | 0 | 14 | 3 | 5385 | 107 |
| P067 | CG | 2.4 | 13 | 4 | 6133 | 74 |
| P067 | CG | 4.5 | 14 | 5 | 5609 | 109 |
| P068 | CG | 0 | 11 | 8 | 5375 | 62 |
| P068 | CG | 2.8 | 11 | 9 | 5289 | 42 |
| P068 | CG | 4.5 | 12 | 10 | 5450 | 73 |
| P069 | CG | 0 | 31 | 18 | 6548 | 75 |
| P069 | CG | 5.8 | 21 | 14 | 8957 | 73 |
| P069 | CG | 9.2 | 18 | 10 | 8204 | 68 |
| P070 | CG | 0 | 17 | 20 | 4189 | 152 |
| P070 | CG | 4 | 15 | 19 | 4282 | 130 |
| P070 | CG | 8.2 | 3 | 9 | 3943 | 136 |
| P071 | CG | 0 | 18 | 10 | 5868 | 105 |
| P071 | CG | 4.1 | 23 | 20 | 5942 | 91 |
| P071 | CG | 7.3 | 23 | 19 | 6954 | 56 |
| P072 | CG | 0 | 8 | 7 | 8539 | 76 |
| P072 | CG | 1.6 | 10 | 5 | 7103 | 129 |
| P072 | CG | 5.1 | 4 | 3 | 6097 | 85 |
| P073 | CG | 0 | 13 | 14 | 8444 | 73 |
| P073 | CG | 3 | 11 | 11 | 7484 | 81 |
| P073 | CG | 4.5 | 9 | 8 | 5885 | 67 |
| P074 | CG | 0 | 7 | 10 | 7444 | 66 |
| P074 | CG | 5.9 | 15 | 14 | 7932 | 78 |
| P074 | CG | 8.5 | 8 | 11 | 7553 | 88 |
| P075 | CG | 0 | 13 | 10 | 7842 | 73 |
| P075 | CG | 3.8 | 49 | 36 | 7984 | 64 |
| P075 | CG | 8.6 | 4 | 6 | 5392 | 42 |
| P076 | CG | 0 | 7 | 15 | 3181 | 309 |
| P076 | CG | 2.6 | 10 | 8 | 3746 | 426 |
| P076 | CG | 7.2 | 8 | 7 | 4430 | 396 |
| P077 | CG | 0 | 15 | 7 | 5002 | 63 |
| P077 | CG | 3.1 | 23 | 19 | 5209 | 54 |

| | | | | | | |
|---|---|---|---|---|---|---|
| P077 | CG | 8.1 | 44 | 32 | 4746 | 80 |
| P078 | CG | 0 | 3 | 5 | 6485 | 47 |
| P078 | CG | 2.3 | 10 | 5 | 6434 | 52 |
| P078 | CG | 4.8 | 2 | 2 | 5717 | 54 |
| P079 | CG | 0 | 8 | 8 | 7277 | 77 |
| P079 | CG | 5.6 | | | | |
| P079 | CG | 7.1 | 10 | 13 | 7543 | 92 |
| P080 | CG | 0 | 10 | 15 | 7060 | 58 |
| P080 | CG | 1.6 | 8 | 9 | 6580 | 52 |
| P080 | CG | 5.5 | 10 | 10 | 6469 | 48 |

#### Table S3F. Albumin and iron-related laboratory fields

De-identified individual participant data are shown in source-workbook row order. Patient_ID, Group and Time_Month are repeated across Table S3A–F for row-level cross-reference. Blank cells indicate missing values; no imputation was applied. Ca×P is the source-workbook product of Ca and P. Field definitions, units and availability are given in Table S4.

| Patient_ID | Group | Time_Month | ALB | Fe | Ferritin | TIBC |
|---|---|---|---|---|---|---|
| P001 | EG | 0 | 45 | 6.07 | 21.4 | 60.27 |
| P001 | EG | 1.7 | 44 | 7.76 | 10.3 | 69 |
| P001 | EG | 2.9 | 47 | 11.79 | 106.2 | 48.26 |
| P002 | EG | 0 | 45 | 10.53 | 381.9 | 68.1 |
| P002 | EG | 3.6 | 44 | 11.56 | 343 | 47.6 |
| P002 | EG | 6.7 | 45 | 7.85 | 168.1 | 52.91 |
| P003 | EG | 0 | 44 | 15.68 | 142.1 | 56.28 |
| P003 | EG | 5.2 | 45 | 13.26 | 365 | 53.01 |
| P003 | EG | 8.8 | 46 | 19.29 | 291.8 | 53.47 |
| P004 | EG | 0 | 45 | 15.33 | 269.2 | 68.42 |
| P004 | EG | 2.2 | 44 | 27.46 | 217.3 | 63.44 |
| P004 | EG | 8.2 | 45 | 8.95 | 29.4 | 53.54 |
| P005 | EG | 0 | 43 | 11.8 | 56.2 | 42.38 |
| P005 | EG | 3.8 | 43 | 14.04 | 155 | 44.19 |
| P005 | EG | 6.1 | 43 | 14.04 | 44.5 | 38.42 |
| P006 | EG | 0 | 41 | 10.77 | 98.2 | 33.41 |
| P006 | EG | 3 | 44 | 9.8 | 149 | 46.67 |
| P006 | EG | 6.8 | 42 |  |  |  |
| P007 | EG | 0 | 44 | 15.9 | 162.9 | 73.33 |
| P007 | EG | 2.6 | 43 | 15.22 | 340 | 47.16 |
| P007 | EG | 4.8 | 48 | 12.79 | 57.8 | 58.86 |
| P008 | EG | 0 | 45 | 17.43 | 113.4 | 70.18 |
| P008 | EG | 7.9 | 40 | 15.38 | 5.8 | 43.59 |
| P008 | EG | 10.9 | 43 | 7.5 | 4.1 | 50.17 |
| P009 | EG | 0 | 40 | 18.93 | 219.3 | 65.66 |
| P009 | EG | 3.8 | 37 | 18.26 | 691 | 44.44 |
| P009 | EG | 7.1 | 46 | 15.31 | 350.9 | 58.78 |
| P010 | EG | 0 | 45 | 22.42 | 20.3 | 81.18 |
| P010 | EG | 1.4 | 47 | 31.7 | 345 | 46.59 |
| P010 | EG | 6.8 | 40 |  |  |  |
| P011 | EG | 0 | 53 | 20.92 | 15.3 | 56.52 |
| P011 | EG | 5.2 | 45 | 12.4 | 28.1 | 55.96 |
| P011 | EG | 8 | 48 | 20.36 | 76.1 | 53.76 |
| P012 | EG | 0 | 44 | 14.14 | 66.7 | 44.61 |
| P012 | EG | 6.5 | 40 | 16.42 | 64.2 | 42.72 |
| P012 | EG | 9.8 | 43 | 9.86 | 32.4 | 50.3 |
| P013 | EG | 0 | 43 | 19.5 | 1033 | 40.5 |
| P013 | EG | 3.4 | 46 | 22.11 | 1938 | 43.83 |
| P013 | EG | 5.8 | 42 | 14.21 | 1575 | 46.29 |
| P014 | EG | 0 | 46 | 17.03 | 25.8 | 93.54 |
| P014 | EG | 3.2 | 43 | 9.93 | 56.4 | 51.97 |
| P014 | EG | 5.2 | 44 | 8.58 | 32.2 | 69.04 |
| P015 | EG | 0 | 46 | 15.85 | 404.5 | 43.18 |
| P015 | EG | 1.5 | 46 | 18.27 | 355 | 45.81 |
| P015 | EG | 3.9 | 43 | 10.62 | 172.2 | 35.58 |
| P016 | EG | 0 | 50 | 7.11 | 22 | 50.27 |
| P016 | EG | 3.1 | 48 | 24.7 | 81.5 | 57.24 |
| P016 | EG | 5.4 | 52 | 11.4 | 80.9 | 57.6 |
| P017 | EG | 0 | 41 | 21.27 | 205.3 | 70.99 |
| P017 | EG | 2.8 | 41 | 14.57 | 194 | 46.45 |
| P017 | EG | 6.1 | 43 | 17.38 | 77.3 | 69.01 |
| P018 | EG | 0 | 40 | 15.39 | 153.4 | 40.57 |

| | | | | | | |
|---|---|---|---|---|---|---|
| P018 | EG | 1.6 | 40 | 14.2 | 306 | 47.65 |
| P018 | EG | 6.1 | 42 | 9.49 | 524.8 | 40.8 |
| P019 | EG | 0 | 40 | 6.92 | 117.9 | 40.45 |
| P019 | EG | 4.7 | 44 | 20.22 | 382 | 59.84 |
| P019 | EG | 9.6 | 43 | 13.97 | 35.7 | 64.01 |
| P020 | EG | 0 | 47 | 15.61 | 142.3 | 53.5 |
| P020 | EG | 2.9 | 44 | 6.46 | 24.1 | 67.59 |
| P020 | EG | 5.7 | 34 | 4.33 | 57.3 | 46.39 |
| P021 | EG | 0 | 42 | 19.28 | 270 | 35.97 |
| P021 | EG | 2.7 | 42 | 12.03 | 208 | 33.09 |
| P021 | EG | 6.6 | 42 | 9.66 | 216 | 42.89 |
| P022 | EG | 0 | 43 | 14.85 | 38.4 | 49.27 |
| P022 | EG | 2 | 43 | 7.61 | 32.5 | 63.08 |
| P022 | EG | 6.7 | 39 | 9.05 | 16.5 | 53.14 |
| P023 | EG | 0 | 46 | 19.64 | 285.8 | 33.88 |
| P023 | EG | 2.5 | 43 | 19.37 | 640 | 47.14 |
| P023 | EG | 6.4 | 43 | 16.41 | 458 | 36.32 |
| P024 | EG | 0 | 45 | 13.34 | 46.8 | 66.52 |
| P024 | EG | 4.1 | 48 | 18.69 | 171 | 49.05 |
| P024 | EG | 6.1 | 45 | 24.56 | 233.6 | 39.99 |
| P025 | EG | 0 | 42 | 15.56 | 34 | 78.05 |
| P025 | EG | 2.8 | 44 | 11.42 | 55.9 | 53.22 |
| P025 | EG | 6.1 | 44 | 9.41 | 11.3 | 53.87 |
| P026 | EG | 0 | 40 | 16.61 | 26.8 | 56.56 |
| P026 | EG | 4.6 | 37 | 10.73 | 31.9 | 52.68 |
| P026 | EG | 8.1 | 35 | 11.72 | 109.2 | 30.35 |
| P027 | EG | 0 | 39 | 15.45 | 49.3 | 32.3 |
| P027 | EG | 3.2 | 41 | 14.23 | 120 | 40.78 |
| P027 | EG | 6.3 | 40 | 26.23 | 71.8 | 54.8 |
| P028 | EG | 0 | 48 | 25.19 | 116.6 | 44.23 |
| P028 | EG | 2 | 47 | 6.96 | 24.2 | 62.49 |
| P028 | EG | 5.3 | 49 | 13.31 | 110.7 | 50.22 |
| P029 | EG | 0 | 39 | 16.08 | 499.4 | 40.74 |
| P029 | EG | 1.5 | 40 | 17.61 | 772 | 45.08 |
| P029 | EG | 6.2 | 42 | 11.57 | 1223.8 | 46.32 |
| P030 | EG | 0 | 43 | 11.94 | 100.8 | 55.3 |
| P030 | EG | 3 | | 13.72 | 220 | 53.17 |
| P030 | EG | 8.1 | 44 | 5.22 | 38.1 | 43.62 |
| P031 | EG | 0 | 41 | 15.88 | 160.8 | 41.77 |
| P032 | EG | 0 | 42 | 15.49 | 354.7 | 30.64 |
| P032 | EG | 2.6 | 44 | 13.42 | 705 | 40.96 |
| P032 | EG | 5.6 | 45 | 12.89 | 204.9 | 45.85 |
| P033 | EG | 0 | 41 | 9.57 | 109.2 | 63.26 |
| P033 | EG | 2.8 | 39 | 6.98 | 75.9 | 44 |
| P033 | EG | 6.6 | 40 | 10.06 | 100.4 | 44.13 |
| P034 | EG | 0 | 42 | 12.66 | 95.6 | 21.84 |
| P034 | EG | 1.2 | | | | |
| P035 | EG | 0 | 45 | 19.38 | 482.2 | 84.65 |
| P035 | EG | 4 | 41 | 11.62 | 895 | 39.04 |
| P035 | EG | 7.3 | 41 | 15.24 | 724.7 | 43.01 |
| P036 | EG | 0 | 40 | 8.1 | 22.8 | 40.5 |
| P036 | EG | 5.1 | 43 | 12.05 | 132 | 59.46 |
| P036 | EG | 9 | 44 | 1.43 | 13.1 | 50.67 |
| P037 | EG | 0 | 45 | 12.71 | 287.1 | 36.04 |
| P037 | EG | 2.2 | 44 | 17.73 | 699 | 48.58 |
| P037 | EG | 6.5 | 43 | 8.65 | 131.4 | 36.15 |
| P038 | EG | 0 | 42 | 8.38 | 5.4 | 44.22 |
| P038 | EG | 5.4 | 42 | 13.86 | 15.6 | 52.34 |

| | | | | | | |
|---|---|---|---|---|---|---|
| P038 | EG | 7 | 41 | 19.78 | 39.4 | 46.79 |
| P039 | EG | 0 | 38 | 26.72 | 46.8 | 46.18 |
| P039 | EG | 1.5 | 37 | 20.08 | 34.2 | 59.1 |
| P039 | EG | 6.2 | 61 | 5.82 | 22.6 | 70 |
| P040 | EG | 0 | 44 | 17.8 | 48.9 | 40.14 |
| P040 | EG | 1.8 | 51 | 17.37 | 155 | 47.42 |
| P040 | EG | 3.9 | 43 | 13.02 | 134 | 45.71 |
| P041 | CG | 0 | 48 | 13.3 | 743.4 | 51.27 |
| P041 | CG | 4.8 | 45 | 11.74 | 716.2 | 55.76 |
| P041 | CG | 9.3 | 43 | 10.45 | 513.3 | 37.07 |
| P042 | CG | 0 | 33 | | | |
| P042 | CG | 4.2 | 45 | | | |
| P042 | CG | 5.9 | 39 | | | |
| P043 | CG | 0 | 45 | 10.02 | 17.1 | 56.58 |
| P043 | CG | 3.6 | 48 | 5.21 | 12.5 | 62.57 |
| P043 | CG | 5.1 | 59 | 10.94 | 63.1 | 52.84 |
| P044 | CG | 0 | 42 | 19.3 | 20.4 | 110.3 |
| P044 | CG | 5.1 | 46 | 16.09 | 90.3 | 42.42 |
| P044 | CG | 8.8 | | 24.9 | 33.4 | 46.17 |
| P045 | CG | 0 | 44 | 17.48 | 1134 | 35.03 |
| P045 | CG | 2.1 | 47 | 10.08 | 849 | 32.8 |
| P045 | CG | 6.1 | 46 | 10.29 | 906 | 41.09 |
| P046 | CG | 0 | 34 | 11.15 | 85.2 | 50.11 |
| P046 | CG | 1.8 | 32 | | | |
| P046 | CG | 7 | 32 | | | |
| P047 | CG | 0 | | | | |
| P047 | CG | 5.3 | | | | |
| P047 | CG | 10.1 | | | | |
| P048 | CG | 0 | 45 | 11.25 | 64.4 | 47.34 |
| P048 | CG | 4.5 | 44 | 22.91 | 349 | 57.05 |
| P048 | CG | 7.2 | 43 | 15.21 | 136.5 | 40.55 |
| P049 | CG | 0 | 42 | 12.19 | 14.5 | 58.69 |
| P049 | CG | 5.5 | 40 | 12.49 | 17.3 | 40.39 |
| P049 | CG | 7.8 | 43 | 8.86 | 28 | 49.57 |
| P050 | CG | 0 | 32 | | | |
| P050 | CG | 4.6 | 35 | | | |
| P050 | CG | 6.4 | 35 | | 52.1 | |
| P051 | CG | 0 | 43 | 16.21 | 117.7 | 45.94 |
| P051 | CG | 3.3 | 44 | 17.42 | 189.8 | 47.51 |
| P051 | CG | 8.7 | 40 | 20.48 | | 45.5 |
| P052 | CG | 0 | 47 | 14.28 | 24.5 | 66.17 |
| P052 | CG | 3.4 | 48 | 21.69 | 29.4 | 70.44 |
| P052 | CG | 6.2 | 47 | 16.88 | 40.2 | 57.38 |
| P053 | CG | 0 | 42 | 13.16 | 171 | 44.47 |
| P053 | CG | 2.2 | 41 | 13.29 | 92 | 44.07 |
| P053 | CG | 7.8 | 44 | 15.47 | 277 | 42.95 |
| P054 | CG | 0 | 78 | 14.08 | 170.5 | 54.21 |
| P054 | CG | 1.5 | 44 | 16.86 | 113.1 | 95.28 |
| P054 | CG | 3.7 | 46 | 18.31 | 156.6 | 40.84 |
| P055 | CG | 0 | 45 | 9.66 | | 70.12 |
| P055 | CG | 5 | 41 | 9.46 | 38 | 39.49 |
| P055 | CG | 9.7 | 43 | 10.12 | 13.9 | 51.38 |
| P056 | CG | 0 | 42 | 20.64 | 63.4 | 65.95 |
| P056 | CG | 2.5 | 44 | 17.27 | 52.8 | 41.57 |
| P057 | CG | 0 | 48 | 13.35 | 269 | 64 |
| P057 | CG | 3.9 | 46 | 19.36 | 409.5 | 28.55 |
| P057 | CG | 9 | 48 | 14.62 | 488 | 67.54 |
| P058 | CG | 0 | 34 | 14.25 | 154 | 35 |

| | | | | | | |
|---|---|---|---|---|---|---|
| P058 | CG | 1.5 | 30 | 24.1 | 310 | 51.1 |
| P058 | CG | 5.1 | 38 | 19.4 | 348 | 67.6 |
| P059 | CG | 0 | 40 | 14.91 | 110 | 56.65 |
| P059 | CG | 1.5 | 43 | 15.09 | 25.9 | 56.3 |
| P059 | CG | 3.3 | 40 | 16.7 | 29.6 | 63.04 |
| P060 | CG | 0 | 47 | 15.96 | 380 | 50.87 |
| P060 | CG | 1.7 | 44 | 13.13 | 346 | 49.97 |
| P060 | CG | 5.5 | 43 | 11.7 | 317 | 48 |
| P061 | CG | 0 | 48 | 13.31 | 9.4 | 110.14 |
| P061 | CG | 4.4 | 45 | 12.72 | 11.1 | 47.61 |
| P061 | CG | 7.7 | 49 | 16.87 | 109 | 43.67 |
| P062 | CG | 0 | 46 | 17.65 | 134.4 | 37.45 |
| P062 | CG | 5.2 | 44 | 7.1 | 41.2 | 55.96 |
| P062 | CG | 9.6 | | 12.94 | 151.9 | 56.55 |
| P063 | CG | 0 | | 11.11 | | 57.05 |
| P063 | CG | 1.9 | | 9.43 | | 47.65 |
| P063 | CG | 3.5 | | | | |
| P064 | CG | 0 | 38 | 18.95 | | 113.44 |
| P064 | CG | 4.7 | 37 | | 315 | |
| P064 | CG | 9.7 | 35 | 9.12 | 38.7 | 44.47 |
| P065 | CG | 0 | 42 | 9.68 | 62.4 | 57.66 |
| P065 | CG | 2.7 | 45 | 11.15 | 25.3 | 57.29 |
| P065 | CG | 7 | 43 | 9.62 | 4.2 | 58.28 |
| P066 | CG | 0 | 45 | 11.71 | 17.7 | 62.45 |
| P066 | CG | 3.5 | 46 | 35.94 | 102.3 | 38.23 |
| P066 | CG | 5.9 | 43 | 16.9 | 58.3 | 51.24 |
| P067 | CG | 0 | 38 | 13.07 | 65.4 | 77.61 |
| P067 | CG | 2.4 | 41 | 23.48 | 204.2 | 40.65 |
| P067 | CG | 4.5 | 42 | 23.07 | 599 | 50.24 |
| P068 | CG | 0 | 43 | 12.82 | 144 | 63.41 |
| P068 | CG | 2.8 | 44 | 16.37 | 217.9 | 33.1 |
| P068 | CG | 4.5 | 44 | 9.75 | 230.6 | 35.71 |
| P069 | CG | 0 | 41 | 11.96 | 52.7 | 45.67 |
| P069 | CG | 5.8 | 41 | 23.85 | 411.9 | 34.75 |
| P069 | CG | 9.2 | 44 | 16.82 | 311.2 | 69.88 |
| P070 | CG | 0 | 41 | 8.1 | 35.9 | 78.28 |
| P070 | CG | 4 | 42 | 19.03 | 123.5 | 82.29 |
| P070 | CG | 8.2 | 40 | 24.12 | 175.9 | 56.08 |
| P071 | CG | 0 | 44 | 10.72 | 388.9 | 50.35 |
| P071 | CG | 4.1 | 40 | 10.01 | 256.8 | 57.98 |
| P071 | CG | 7.3 | 43 | 11.32 | 325.4 | 49.4 |
| P072 | CG | 0 | 43 | 10.67 | 164.8 | 36.63 |
| P072 | CG | 1.6 | 43 | 12.66 | 26.4 | 46.88 |
| P072 | CG | 5.1 | 37 | 9.37 | 127 | 28.07 |
| P073 | CG | 0 | 25 | 10.8 | 101.2 | 41.33 |
| P073 | CG | 3 | 30 | 14.43 | 74.6 | 38.01 |
| P073 | CG | 4.5 | 30 | 12.25 | 155 | 35.83 |
| P074 | CG | 0 | 44 | 12.29 | 59.4 | 59.72 |
| P074 | CG | 5.9 | 46 | 7.33 | 113 | 56.16 |
| P074 | CG | 8.5 | 43 | 13.97 | 35.7 | 64.01 |
| P075 | CG | 0 | 36 | 10.59 | 38.1 | 44.91 |
| P075 | CG | 3.8 | 30 | 10.23 | 142 | 39.33 |
| P075 | CG | 8.6 | 47 | 8.56 | 121 | 58.6 |
| P076 | CG | 0 | 40 | 15.44 | 21.3 | 101.23 |
| P076 | CG | 2.6 | 43 | 6.98 | 21.8 | 57.66 |
| P076 | CG | 7.2 | 42 | | | |
| P077 | CG | 0 | 37 | 7.06 | 123.1 | 37.48 |
| P077 | CG | 3.1 | 38 | 9.33 | 135.5 | 38.55 |

| | | | | | | |
|---|---|---|---|---|---|---|
| P077 | CG | 8.1 | 38 | 3.7 | 211.3 | 41.85 |
| P078 | CG | 0 | 44 | 10.67 | 27.2 | 61.42 |
| P078 | CG | 2.3 | 45 | 9.68 | 20.1 | 44.77 |
| P078 | CG | 4.8 | 44 | 5.9 | 16 | 38.11 |
| P079 | CG | 0 | 35 | 7.21 | 114.5 | 32.29 |
| P079 | CG | 5.6 | | 12.47 | 148 | 30.98 |
| P079 | CG | 7.1 | 39 | 12.02 | 150 | 38.09 |
| P080 | CG | 0 | 46 | 15.06 | 222 | 51.47 |
| P080 | CG | 1.6 | 44 | 9.76 | | 44.97 |
| P080 | CG | 5.5 | 41 | 10.12 | 481 | 25.53 |

#### Table S4. Clinical data dictionary and outcome-specific data availability

Counts were calculated directly from the 236-row, 80-participant source workbook. “Participants” denotes the number of unique Patient_ID values with at least one non-missing value for the field. Units marked “Not stated in source workbook” require author verification; no unit has been inferred solely from laboratory convention.

| Field | Definition | Unit | Non-missing records | Participants |
|---|---|---|---|---|
| Patient_ID | De-identified participant identifier | Not applicable | 236 | 80 |
| Group | Randomised group: CG (placebo) or EG (TEX) | Not applicable | 236 | 80 |
| Sex | Sex recorded in the source workbook | Not applicable | 236 | 80 |
| Age | Five-year age band | years | 236 | 80 |
| BMI | Body mass index | $kg/m^2$ | 236 | 80 |
| Time_Month | Elapsed time from baseline | months | 236 | 80 |
| CREA | Serum creatinine (Scr) | μmol/L | 236 | 80 |
| BUN | Blood urea nitrogen | mmol/L | 236 | 80 |
| eGFR | Estimated glomerular filtration rate | $mL/min/1.73\ m^2$ | 236 | 80 |
| Cys_c | Serum cystatin C | mg/L | 200 | 79 |
| Ca | Serum calcium | mmol/L | 236 | 80 |
| P | Serum phosphate | mmol/L | 236 | 80 |
| Ca×P | Calcium–phosphorus product, derived as Ca × P | $mmol^2/L^2$ (derived) | 236 | 80 |
| PTH | Parathyroid hormone | pg/mL | 218 | 79 |
| NEUT | Neutrophil count | Not stated in source workbook | 236 | 80 |
| LYMPH | Lymphocyte count | Not stated in source workbook | 236 | 80 |
| WBC | White blood cell count | Not stated in source workbook | 236 | 80 |
| ALB | Albumin | g/L | 225 | 78 |
| Hb | Haemoglobin | g/L | 236 | 80 |
| AST | Aspartate aminotransferase | U/L | 225 | 78 |
| ALT | Alanine aminotransferase | U/L | 225 | 78 |
| CHE | Cholinesterase | U/L | 225 | 78 |
| ALP | Alkaline phosphatase | Not stated in source workbook | 225 | 78 |
| Fe | Iron | Not stated in source workbook | 219 | 77 |
| Ferritin | Ferritin | Not stated in source workbook | 215 | 77 |
| TIBC | Total iron-binding capacity | Not stated in source workbook | 219 | 77 |
